# β_1_- and β_2_-adrenergic Receptor Haplotypes Regulate Therapeutic Responses to Placebo and the Biased Ligand β-blocker Bucindolol

**DOI:** 10.1101/2025.08.22.25333921

**Authors:** Ian A. Carroll, Dobromir Slavov, Eric R. Jonas, Wayne A. Minobe, Lori A. Walker, Amrut V. Ambardekar, Timothy A. McKinsey, Ammon D. Knaupp, Donghwa Kim, Stephen B. Liggett, Michael R. Bristow

## Abstract

**Background:** *ADRB1* and *ADRB2,* encoding cardiac myocyte β_1_- and β_2_-adrenergic receptors (ARs) that mediate pathologic myocardial remodeling in response to chronically increased signaling, contain N-terminus haplotype variants capable of influencing agonist- or biased ligand-induced receptor internalization that uncouples canonical signaling and initiates EGFR/ERK1/2 cardioprotection.

**Methods:** In two heart failure (HF) clinical trial genetic substudies we investigated effects of internalizing vs. internalization-resistant *αDRB1*/*ADRB2* haplotypes on clinical or biomarker responses to the biased ligand β-blocker bucindolol vs. placebo or vs. the nonbiased β_1_-antagonist metoprolol, and in haplotyped isolated human heart preparations we measured ERK1/2 activation in response to these same interventions.

**Results:** In subjects with ≥3 internalizing *ADRB1*+*ADRB2* haplotypes (6.7% subcohort) placebo treatment was associated with fewer clinical events compared to subjects with internalization-resistant haplotypes (Odds Ratio (OR) 0.28, 95% CI (0.10, 0.82)). In contrast, placebo treatment in subjects with ≥3 internalization-resistant haplotypes (70% subcohort) was associated with more clinical events in comparison to subjects with internalizing haplotype counterparts (OR 1.64 (1.46, 1.84)). Bucindolol treatment was equal to placebo in the ≥3 internalizing subcohort, but was superior to placebo in the internalization-resistant subcohort (bucindolol vs. placebo OR 0.49 (0.41, 0.58)). In subjects with all 4 haplotypes internalization-resistant (25% subcohort), bucindolol vs. placebo reduced time to first event rates by 62.3±17.5% (P <0.01, 1.68±0.34 fold > the all-haplotypes parent population and additive to 1.92±0.58 fold when the *ADRB1* haplotype contained Arg389 rather than Gly389). The same bucindolol vs. placebo pattern was observed for NT-proBNP or norepinephrine reduction vs. metoprolol. In these comparisons *ADRB2* and *ADRB1* haplotypes behaved similarly, and although the haplotypes differed in frequency between Black and non-Black subjects, within haplotypes there were no by-race differences in therapeutic effects. Bucindolol but not metoprolol activated ERK1/2 signaling in isolated ventricular preparations with ≥3 internalization-resistant haplotypes.

**Conclusions:** 1) Both β_1_- and β_2_-AR haplotypes regulate therapeutic responses in HF; internalizing species confer protection against clinical events in placebo-treated subjects, while in internalization-resistant haplotypes the biased ligand β-blocker bucindolol but not the non-biased ligand metoprolol is associated with favorable effects. 2) The biased ligand cardioprotective effect may be related to internalization-dependent or -independent ERK1/2 activation.

## BACKGROUND

Human cardiac myocytes contain 3 types of β-adrenergic receptors (β-ARs), with β_1_-ARs dominating but β_2_-ARs also potentially involved in the regulation of cardiac function and capable of mediating pathologic effects of chronic adrenergic stimulation in the failing heart [1, 2]. In heart failure (HF) increased norepinephrine (NE) signaling contributes to myocardial pathologic remodeling, the inhibition of which results in the beneficial effects of β-blockers in subjects with reduced ejection fraction (HFrEF, LVEF <0.40) [1, 3, 4]. Although chronically increased cardiac adrenergic drive has a net myopathic effect on the myocardium [1, 4] including predisposing to arrhythmias [5], certain cellular mechanisms can counter these adverse effects. The most obvious of these are β-AR desensitization and/or downregulation that diminish adrenergic signaling through canonical cAMP/PKA and CaMKII pathways [1]. In addition, for both β_1_-[6−8] and β_2_-ARs [9−11] desensitization also includes signaling through an alternative, EGFR/ERK1/2 cardioprotective pathway consisting of receptor phosphorylation, binding to β-arrestins, endocytic internalization and endosomal trafficking to fates that include lysosomal degradation [7, 8]. In this series of events receptor internalization is a critical step, as inhibitors of endocytic vesicle formation block EGFR transactivation and ERK1/2 phosphorylation [10, 12]. β-adrenergic agonists classically activate this pathway [6−8, 10−12], but with some differences [13] so can β-blocker “biased ligands”[14−17].

On short-term (≤1 hour) exposure to agonists some β-AR polymorphic variants differ from their counterparts in their abilities to desensitize, internalize, and/or traffic intracellularly. For *ADRB1* the more prominently desensitizing and internalizing β_1_-AR variant is Gly49 [18], which compared to Ser49 also exhibits differences in subsequent intracellular trafficking [19, 20] that are present in both the Arg389 and Gly389 haplotypes [20]. In addition, when in a laboratory-constructed Gly389 haplotype, Gly49 is associated with greater agonist-induced ERK1/2 phosphorylation than the Ser49Gly389 variant [21]. For β_2_-ARs, the Arg16Gln27 haplotype exhibits rapid agonist desensitization *in vivo* compared to other haplotypes [22], and rapidly and substantially desensitizes [23] and internalizes [23−25] on agonist exposure in cells transfected with the original human β_2_-AR cDNA containing the Arg16Gln27 haplotype [26]. Thus both *ADRB1* and *ADRB2* N-terminus nonsynonymous SNPs can affect internalization.

The *ADRB1* position 49 and the *ADRB2* position 16 or 27 polymorphisms do not influence signal transduction efficiency [18−20, 24]. However, the *ADRB1* Ser49Gly polymorphism is in strong linkage disequilibrium (LD) with the nonsynonymous C-terminus SNP Arg389Gly [27] whose encoded variants have major effects on signal transduction capacity and agonist affinity [28, 29]. The *ADRB2* position 16 and 27 variants are also in tight linkage disequilibrium [27]. *ADRB1* 49/389 haplotypes can therefore encode receptors with greater (Arg389) or lesser (Gly389) signaling capacity or agonist affinity that internalize to a greater (Gly49) or lesser (Ser49) extent in the presence of agonists or potentially to biased ligands, while the *ADRB2* Arg16Gln27 vs. other encoded haplotypes have the potential for more vs. less internalization.

In the MAP-MI study [27] we recently reported that the *ADRB1* Gly49Arg389 haplotype is associated with a reduction in incident ventricular fibrillation (VF) in acute myocardial infarction. There was also evidence of a VF protective signal in subjects with the *ADRB2* Arg16Gln27 haplotype [27].

These findings were interpreted as potentially due to EGFR/ERK1/2 cardioprotective pathway signaling [6, 12] that internalization of both β_1_-[8, 12, 30] and β_2_-ARs [10, 31, 32] are known to activate when occupied by an agonist, in this case NE. An earlier study had found that the *ADRB1* Gly49 polymorphism, in genotyping studies typically present only as the Gly49Arg389 haplotype, is associated with improved survival in dilated cardiomyopathy patients, also suggesting a myocardial protective mechanism [33]. Since HFrEF human ventricles contain 35-40% β_2_-ARs that are coupled to inotropic and pro-arrhythmic pathways and when overexpressed in model systems to a cardiomyopathy [1,2], there is also a rationale for β_2_-AR internalization affecting outcomes in HFrEF.

Here we test the hypothesis that internalization or internalization-related effects of *ADRB1* and *ADRB2* common haplotype variants influence clinical outcomes in HFrEF patients treated with placebo, the biased ligand β-blocker bucindolol, or the nonbiased β-blocker metoprolol succinate. The 2 clinical trial cohorts investigated [29, 34] were haplotyped for *ADRB1* and *ADRB2* variants, and one of them (BEST Trial Adrenergic Receptor Polymorphism Substudy [29, 35]) included populations with substantial racial heterogeneity that affects haplotype frequency and thus provides an opportunity for internal confirmation by investigating separate genetic subpopulations. The presented data support the utility of pharmacogenetic targeting of protein haplotypes [36], something that has long held promise [37, 38] but has been difficult to implement.

## METHODS

### Clinical Trials with Genetic Substudies

#### BEST Adrenergic Receptor Polymorphism Substudy

This 1040 patient pharmacogenomics Substudy [29] of the Beta-blocker Evaluation of Survival Trial (BEST, NCT00000560) [35], entitled “Pharmacogenomics of Beta-adrenergic Receptor Polymorphisms and the Response to Beta-blockers in Heart Failure,” had the primary aim of “delineating the relationship between β_1_-AR and β_2_-AR polymorphisms and the clinical response to β-blockers.” Secondary aims were to “determine the relationship between β_1_-AR and β_2_-AR polymorphisms and the natural history of moderate to severe heart failure in patients using the placebo arm of BEST,” and added after the trial was completed, to “see whether these polymorphisms can account for the racial differences in the response to bucindolol that have been observed in BEST.” Two polymorphisms in *ADRB1* (positions 49 and 389) and 3 in *ADRB2* (positions 16, 27, and 164), all originally discovered and characterized in the laboratory of the substudy’s Principal Investigator (Stephen Liggett), were prospectively identified for investigation, and the *ADRB2* Thr164Ile polymorphism was later removed from consideration because of a minor allele frequency (0.012) insufficient for analysis.

The BEST pharmacogenomics substudy patients had advanced HFrEF (NYHA Class III-IV HF, LVEFs ≤0.35) with elevated plasma NE levels (**Table S1, Supplemental Material**) who were randomized 1:1 to treatment with placebo or the nonselective β_1_-, β_2_-AR blocker/biased ligand bucindolol. Eligible subjects gave written informed consent for the parent trial [35] and as an option for peripheral blood DNA extraction and genotyping in the Substudy [29]. The primary endpoint of the pharmacogenomics substudy was time to all-cause mortality or cardiac transplantation.

#### GENETIC-AF

The GENETIC-AF trial (NCT01970501), a Phase 2 trial that randomized HFrEF and HFmrEF (HF with LVEF 0.40-0.49) patients to bucindolol or metoprolol succinate who were *ADRB1* Arg389Arg genotype (N=267) and at risk for developing recurrent AF [34], included a voluntary DNA Bank substudy for which 138 patients signed written informed consent. This substudy utilized banked DNA from extracted whole blood samples used for the qualifying *ADRB1* Arg389Arg testing, which was subsequently genotyped for *ADRB1* Ser49Gly and *ADRB2* position 27 and 16 polymorphisms. The Primary Aim of the Substudy was association of adrenergic receptor polymorphisms with NT-proBNP and NE biomarker measurements, with changes from baseline in these biomarkers being the primary endpoints. The primary endpoint of the parent trial was time to AF, atrial flutter or ACM (AF/AFL/ACM).

### Study Approvals

The BEST trial and its pharmacogenomics substudy were approved by ethical committees at the NHLBI, Department of Veterans affairs (Palo Alto VA, location of the statistical analysis center), the DNA Oversight Committee and each investigational site. The GENETIC-AF study and its DNA substudy were approved at each investigational site. Participants in each clinical trial signed written informed consent for both the parent trial and separately for voluntary DNA sampling that could be used for genetic or other analyses with de-identification of individual subjects.

### Race as Modifier of Polymorphism and Haplotype Frequencies

The race/ethnicity case report form classifications in both clinical trials were “White, Black, Hispanic, Asian/Pacific Islander, American Indian/Alaskan, Other” determined by self-identification. In the current as well as previous reports [35], Blacks were assigned one racial category, and all other races were classified as non-Blacks.

### Genotyping and Haplotyping

BEST Pharmacogenomics Substudy genotyping for *ADRB1* Arg389Gly, *ADRB1* Ser49Gly, *ADRB2* Gln27Glu, and *ADRB2* Gly16Arg was conducted by Restriction Fragment Length Polymorphism (RFLP) conducted in DNA extracted from whole blood as previously described [29, 39] with results subsequently confirmed by DNA sequencing. For GENETIC-AF subjects the *ADRB1* Arg389Arg genotype necessary for trial enrollment was determined by TaqMan^®^ SNP genotyping as previously described [34], while the *ADRB1* Ser49Gly genotype and *ADRB2* genotypes were measured by sequencing SNP flanking-primer PCR amplicons using DNA extracted for the *ADRB1* Arg389Gly genotyping. Haplotyping in both substudies was via an expectation-maximization (EM) algorithm for maximum likelihood estimates, using the “Haplo.stats” package available in R [27, 40]. Linkage disequilibrium was tested using the R based “genetics” package (https://CRAN.R-project.org/package=genetics). Haplotypes for pairwise SNPs were only ambiguous in the double heterozygote case; thus, for both *ADRB1* and *ADRB2* 20 double genotype heterozygote DNA samples were subjected to long-read sequencing [41] that confirmed the likelihood reads of missingness of 1 haplotype from each gene (Gly49Gly389 and Arg16Glu27, the double minor allele variants of each haplotype).

### Quantitation and Statistical Analysis

#### General Strategy for Clinical Trial Substudy Data

The overall aim of the statistical analysis was to compare clinical event and biomarker outcomes in HF subjects with more vs. fewer internalizing β-adrenergic receptor (β-AR) haplotypes, across 2 clinical trials where haplotyping was performed and which had extensive outcomes databases. We thus designed a statistical approach to be consistent with each trial’s original statistical analysis plan, but would also address the current primary hypothesis that β-AR haplotypes with different internalization potential affect HF outcomes. Because of variations in cohort sample sizes internalization subdivision definitions varied slightly between analyses (described in detail in the second paragraph of **Supplemental Material**, **Conventions and Presentation Formats** and summarized in **Table S2**) but always included a comparison that encompassed internalizing vs. internalization-resistant (*ADRB1*+*ADRB2*) haplotypes.

The greater sample size of the BEST Pharmacogenomics Substudy allowed more gradation in estimation of dose-response effects of haplotype internalization (i.e., sufficient sample size for analyses using three mutually-exclusive tiers and two separate treatment arms), whereas the GENETIC-AF DNA substudy, limited in sample size and restricted in *ADRB1* genotype to Arg389Arg, maintained the directionality of this classification by confining the analysis to two mutually-exclusive tiers that were also analyzed in the BEST cohort.

#### Genotype and Haplotype Analyses

Genotype-level analyses are inherently confounded by effects related to other loci in linkage disequilibrium with the polymorphism of interest. This can be reconciled by analysis at the haplotype level, where 3 haplotypes exist for both *ADRB1* (Gly49Arg389, Ser49Arg389, Ser49Gly389) and *ADRB2* (Arg16Gln27, Gly16Gln27, Gly16Glu27), which manifest independently at the genome (different chromosomes) or statistical (no detectable linkage disequilibrium and satisfying Hardy-Weinberg equilibrium) level. A standard Bonferroni adjustment for internalizing vs. internalization-resistant haplotypes for both *ADRB1* and *ADRB2* would therefore yield α = .025 for an endpoint analysis, which was considered the adjusted α-level.

#### Clinical Endpoints

The clinical endpoints prospectively chosen for analysis included the primary cardiovascular endpoints for both the BEST Pharmacogenomic and GENETIC-AF DNA substudies (ACM/Tx [29] and AF/AFL/ACM [34], respectively), the primary endpoint of the BEST parent trial [35] (ACM) that was a secondary endpoint in its substudy, 2 secondary endpoints from the BEST parent trial (CVM, HFH) [35], a composite endpoints from these secondaries (ACM/HFH), a previously reported primary endpoint used for BEST trial data (CVM/HFH) [42] that has become a favored clinical outcome assessment in HF trials, and 1 post hoc endpoint (CVH) requested by the FDA [43]. Clinical endpoints were analyzed by forest plots, structural equation modeling (SEM), and time to first event (Kaplan-Meier) curves. The methodology for these analyses is further described in **Supplemental Material**, **Methods**.

#### Isolated Human Heart Studies Tissue Bath Preparations

Explanted failing hearts from end stage heart failure patients with nonischemic or ischemic cardiomyopathies were obtained at the time of cardiac transplantation and immediately transported to the laboratory in iced Tyrode’s solution. Right ventricular trabeculae (N=10-14) of uniform size (1-2 x 6-8 mm) were mounted in tissue baths in Tyrode’s solution at 37 and field stimulated to contract at 60 bpm, as previously described [29]. Following a 1 hour equilibration period 1-2 trabeculae/group were incubated with vehicle, isoproterenol (1umol/L), bucindolol HCL(1umol/L) or metoprolol tartrate (10 umol/L) for either 5, 10 or 60 minutes. At the end of its incubation period each trabeculum was removed from the bath, flash frozen in liquid nitrogen and stored at −80°C.

#### pERK1/2 Immunoblotting

These details are in **Supplemental Materials**, **Supplemental Methods**. **Statistical Testing**

#### Statistical Testing

Details of the SEM analyses, including a listing of covariates used for adjustment, can be found in **Supplemental Methods, Structural Equation Modeling**. Time to first event Hazard Ratios (HzR) were generated by Cox proportional hazards models **(Supplemental Methods, Time to First Event Analyses**), with first clinical event Odds Ratios (ORs) obtained from logistic regression models.

Confidence intervals for linkage disequilibrium coefficients were calculated using 10,000 nonparametric bootstrapped estimates. Treatment effects between bucindolol vs. control (placebo or metoprolol succinate) were calculated from HzRs as: % treatment effect = ((1-HzR) x 100). Normality was determined by the Anderson Darling test. For normally distributed data assessing one clinical endpoint t-tests were used, and for >1 endpoint one-way ANOVA with a Holm-Sidak’s test was employed. Analysis of ERK1/2 phosphorylation was by a one-way ANOVA mixed model with repeat measures and a Holm-Sidak multiple comparison test. For non-normally distributed data Wilcoxon or Friedman’s/Kruskal-Wallis tests with a Dunn’s multiplicity adjustment were employed.

## RESULTS

### BEST Adrenergic Receptor Polymorphism Substudy

#### Allele, Genotype, and Haplotype Frequencies; Linkage Disequilibrium

A description of the formatting and terminology used for genotypes, haplotypes and diplotypes is in **Table S2** and the first paragraph of **Supplemental Material**. Allele frequencies of *ADRB1* and *ADRB2* polymorphisms by Black and non-Black self-identified races are in **Table S3**, and are similar to those previously reported for European-vs. African-Ancestry in the BEST Substudy [44]. Blacks have higher minor allele frequencies for *ADRB1* Gly389 and Gly49, and for *ADRB2* Arg16. *ADRB2* Glu27 is lower frequency in Blacks.

Genotypes and theoretical vs. identified diplotypes are in **Table S4.** For both *ADRB1* and *ADRB2*, of 10 theoretical diplotypes only 6 were identified; diplotypes with *ADRB1* Gly49Gly389 and *ADRB2* Arg16Glu27 haplotypes are missing. In each gene identified diplotypes consist of 3 homozygotes, 2 single heterozygotes, and 1 double heterozygote. There is strong LD between *ADRB1* positions 49 and 389 polymorphisms, as well as among *ADRB2* positions 16, 27 and 164 (**Table S5**); the latter polymorphism was very low frequency (minor allele 0.012, **Table S3**) and was not further investigated. The absence of *ADRB1* Gly49Gly389and *ADRB2* Arg16Glu27 haplotypes was confirmed by long-read DNA sequencing conducted in 26 *ADRB1* 49/389 and 50 *ADRB2* 16/27 double heterozygotes. **Table S6** also gives haplotypes by sex within race categories, where there is no difference between sexes within either race category.

Diplotype and haplotype frequencies by race classification are in **Tables 1** and **S6**, respectively. Gly49Arg389 and Ser49Gly389 haplotypes are more prevalent in Blacks, while the major allele haplotype Ser49Arg389 is more prevalent in non-Blacks (**Table S6**). For *ADRB2* Gly16Gln27 as well as Arg16Gln27 are more prevalent in Blacks. The internalizing haplotypes Gly49Arg389 (*ADRB1*) and Arg16Gln27 (*ADRB2*) are respectively 1.71 (P<0.0001) and 1.21 (P=0.004) fold more frequent in Blacks vs. non-Blacks. Internalization-resistant haplotypes are more prevalent in non-Blacks compared to Blacks, by 1.14 (P <0.0001) and 1.15 (P=0.004) fold in *ADRB1* and *ADRB2*, respectively (**Table S6**).

For *ADRB1* diplotypes (**Table 1**), any that contain at least 2 total glycines in positions 49 plus 389 are more prevalent in Blacks, while the Ser49Arg389 homozygous diplotype is more common in non-Blacks. The Gly49Arg389 homozygous diplotype is 2.34 fold more common in Blacks (P =0.012), as is the homozygous Ser49Gly389 diplotype (by 2.43 fold, P <0.0001).

**Table 1.**
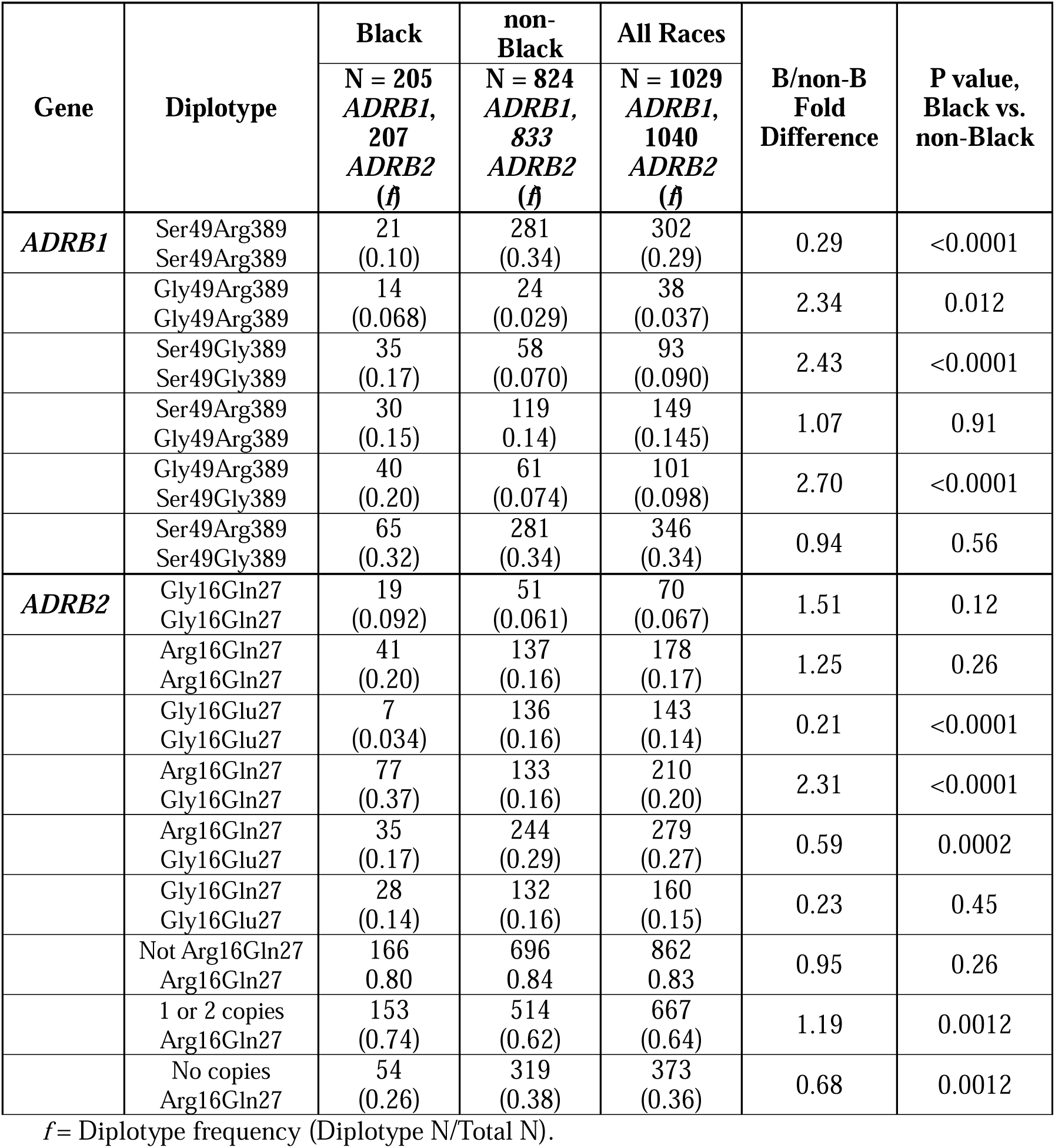
*ADRB1*, *ADRB2* diplotype frequencies (*f*) by race, BEST Adrenergic Receptor Polymorphism Substudy.

#### Baseline Characteristics

Baseline characteristics by diplotype, race and treatment group are in **Table S1**. For diplotypes the main differentiating characteristic is race (**Tables S1** and **S6)**. In comparison to non-Blacks, Blacks are 5 years younger and have higher resting heart rates and systolic blood pressures. There are no differences in baseline characteristics between treatment groups.

#### β-adrenergic Receptor Haplotype Internalization Potential Affects Clinical Outcomes Clinical Events in Internalizing vs. Internalization-resistant Haplotypes

As an initial test of the hypothesis that *ADRB1* and *ADRB2* haplotypes with different internalization potential affect clinical outcomes, in placebo- and bucindolol-treated subjects in the BEST substudy unadjusted odds ratios (ORs (95% C.I.s)) for first clinical event rates were compared between defined {*ADRB1*+*ADRB2*} haplotype internalizing vs. internalization-resistant groups. The index internalizing haplotypes were *ADRB1* Gly49Arg389 and *ADRB2* Arg16Gln27 as tabulated in {*ADRB1*+*ADRB2*} diplotype combinations, which were compared to all other diplotype combinations (see classification nomenclature, **Table S2)**. In the classification descriptors “number” is the sum of the *ADRB1*+*ADRB2* haplotypes in the diplotype combination, “D” (double) indicates that at least 1 defined haplotype is present in both genes, “I” is internalizing, and “IR” is internalization-resistant.

**Figures 1-3** (all-LVEF cohort) and **S1-S3** (cohort restricted to baseline LVEFs of ≥0.20) are forest plots of the between-haplotypes ORs for 7 clinical endpoints that include the primary endpoints for the BEST and GENETIC-AF substudies, respectively time to all-cause mortality or cardiac transplantation (ACM/Tx)^28^ and time to first atrial fibrillation or flutter (AF/AFL/ACM) [34]. To provide a robust statistical analysis as well as to correct for possible confounding variables, a covariate adjusted structural equation model (SEM) endpoint analysis is provided in **Table S7** where clinical events are modeled against increasing numbers of internalization-resistant haplotypes from 0 to 4. All analyses are constructed from the first clinical events in subjects who had complete *ADRB1* and *ADRB2* genotyping. In **Figures 1-3** and **S1-S3** {*ADRB1*+*ADRB2*} diplotype combinations are organized by 3 sections or tiers based on the total number of internalizing {*ADRB1*+*ADRB2*} index haplotypes. **Tier 1** is 3-4 copies of *ADRB1* AND *ADRB2* internalizing haplotypes (“Double-Internalizing”, ≥**3DI**); **Tier 2** has 2 copies of internalizing haplotypes (”Incompletely Internalizing”, **2I**); and **Tier 3** has 0-1 copies of internalizing (3-4 internalization-resistant) haplotypes (”Double Internalization-resistant”, ≥**3DIR**). The sequence of presentation is placebo treated patients (**Figures 1 and S1**), bucindolol patients (**Figures 2 and S2**), and bucindolol vs. placebo treatment effects (**Figures 3 and S3**). ORs in **Figures 1/S1** and **2/S2** are generated by comparing the haplotype pairs in Column 1 to all other haplotype pairs not defined in Column 1, and treatment effect ORs (**Figures 3/S3**) are based on bucindolol vs. placebo event ORs.

**Figure 1.**
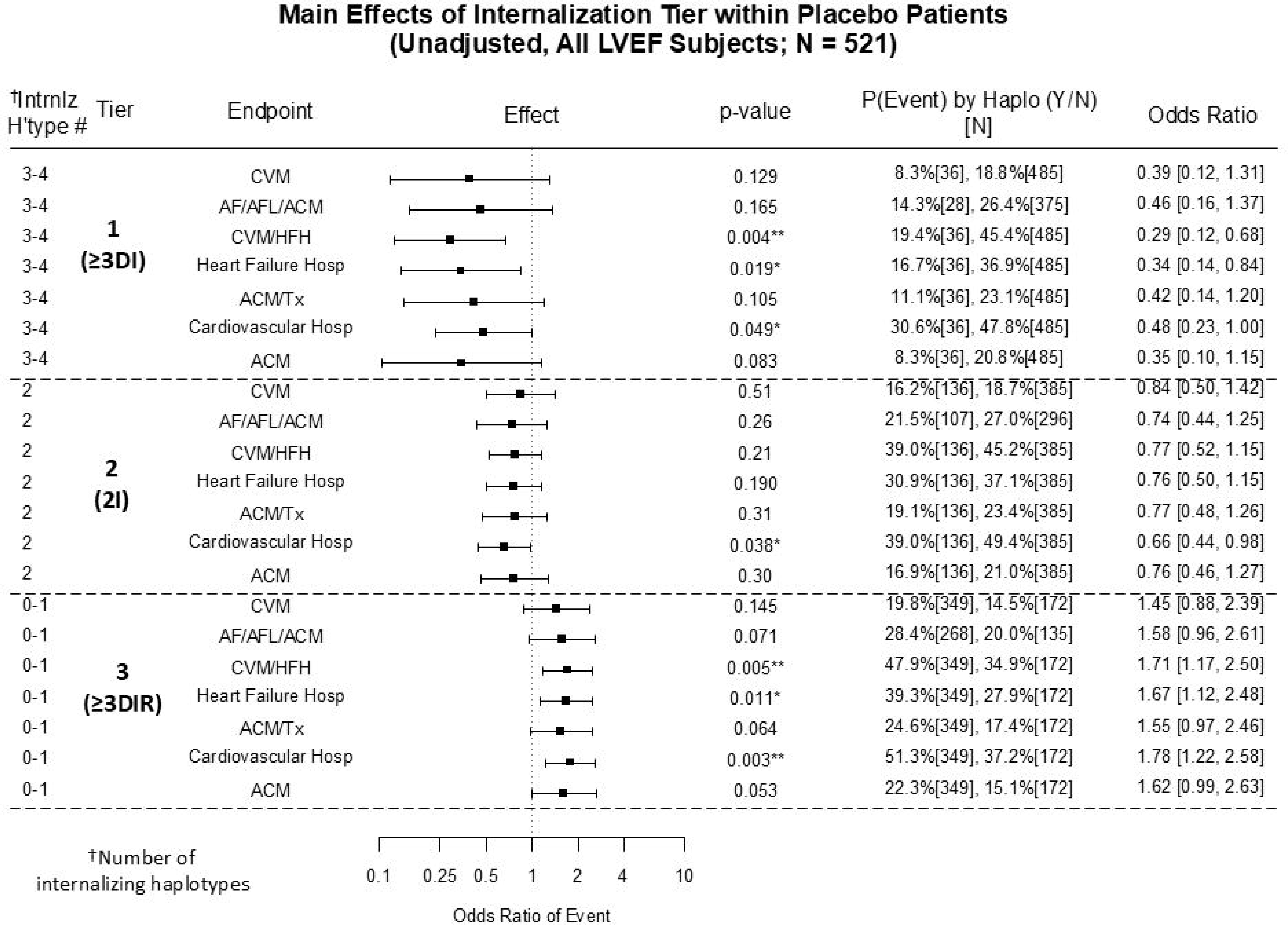
All LVEF cohort patients randomized to placebo in the BEST Adrenergic Receptor Polymorphism substudy, forest plot of *ADRB1+ADRB2* combination diplotypes arranged by the number of internalizing haplotypes (Column 1), displaying Odds Ratios (ORs, Columns 4 and 7) between event rates (Column 6) in the Column 1 diplotypes vs. all other combination diplotypes. The *ADRB1*, *ADRB2* diplotype combinations are displayed by “≥3 Double Internalizing” combinations in **Tier 1** (≥**3DI**, 1 or 2 copies of *ADRB1* Gly49Arg389 AND *ADRB2* Arg16Gln27, for a total of 3-4 copies of an internalizing haplotype); “Incompletely Internalizing” in **Tier 2** (**2I)**, 2 total copies of internalizing haplotypes, either 1 copy of *ADRB1* AND *ADRB2*, or 2 copies on one of them); and “≥3 Double Internalization-resistant” haplotype combinations in **Tier 3** (≥**3DIR,** 0-1 total copies of *ADRB1* Gly49Arg389 AND *ADRB2* Arg16Gln27, 3-4 copies of internalization-resistant haplotypes). The counterpart, comparison haplotypes used to generate ORs follow OR logic, i.e. if either the *ADRB1* or the *ADRB2* haplotype doesn’t match the cognate Column 1 haplotype then the pair becomes a member of the counterpart group. Column 6 lists the event rate and [N] in the paired haplotypes in column 1, followed by the event rate and [N] in the counterpart, comparator groups. CVM=Cardiovascular Mortality; AF/AFL/ACM=Atrial fibrillation, Atrial Flutter or All-Cause Mortality; CVM/HFH=CVM or Heart Failure hospitalization; HFH=Heart Failure Hospitalization; ACM/Tx=ACM or Cardiac Transplantation; CVH=Cardiovascular Hospitalization; ACM=All-Cause Mortality. For the AF/AFL/ACM endpoint, only non-AF patients at the time of randomization are analyzed, resulting in a smaller N than for the 6 HF endpoints. For all endpoints only first events are considered. Odds Ratios (ORs) are calculated by: [(Index event rate in %) / (100 – Index event rate)] / [(Comparator event rate) / (100 − Comparator event rate)], or in Row 1 (8.3/91.7) / (18.8/81.2) = 0.39.

**Figure 2.**
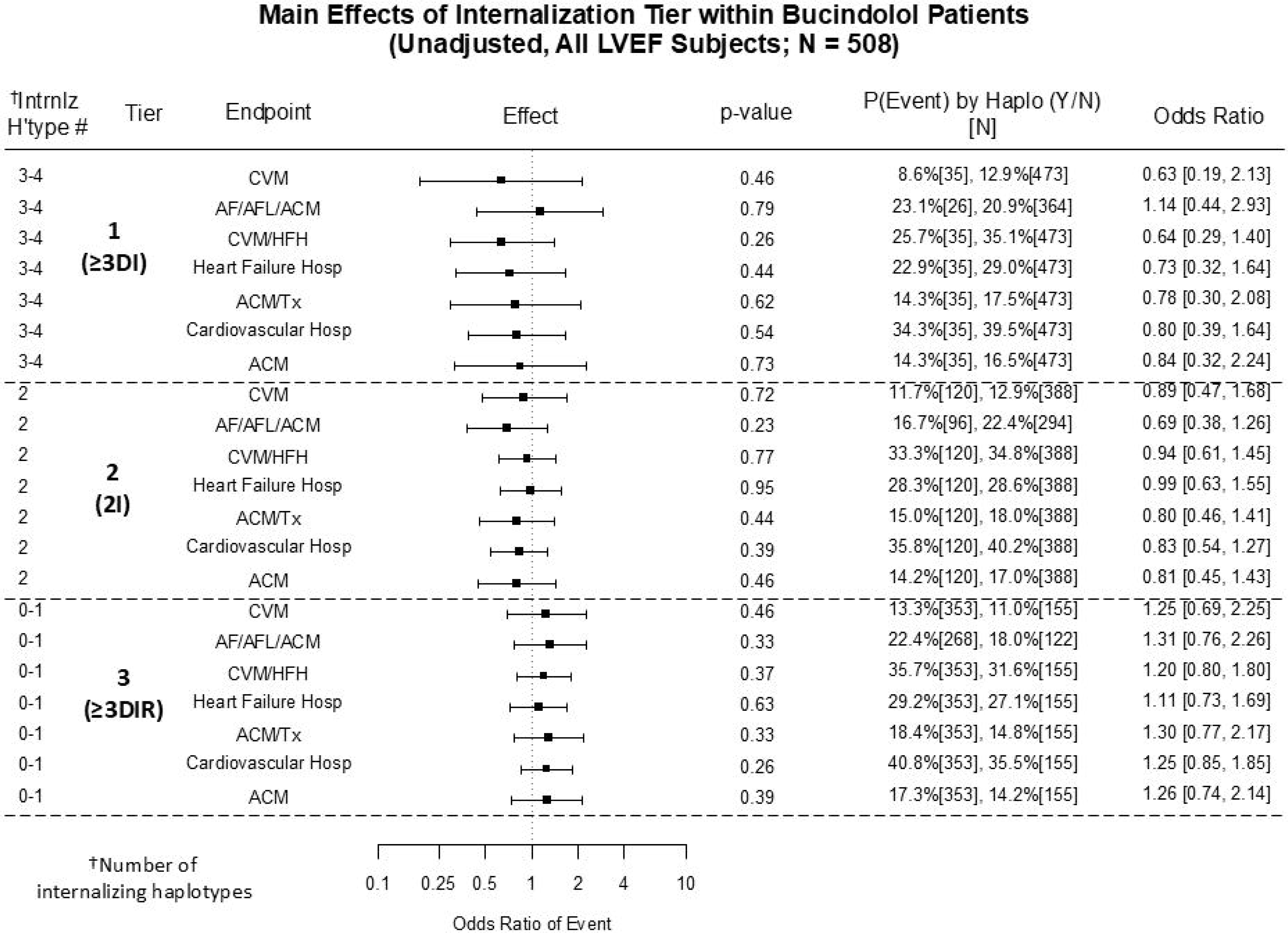
All LVEF cohort BEST substudy patients randomized to bucindolol in the BEST Adrenergic Receptor Polymorphism substudy. The forest plot is constructed as for **Figure 1**.

**Figure 3.**
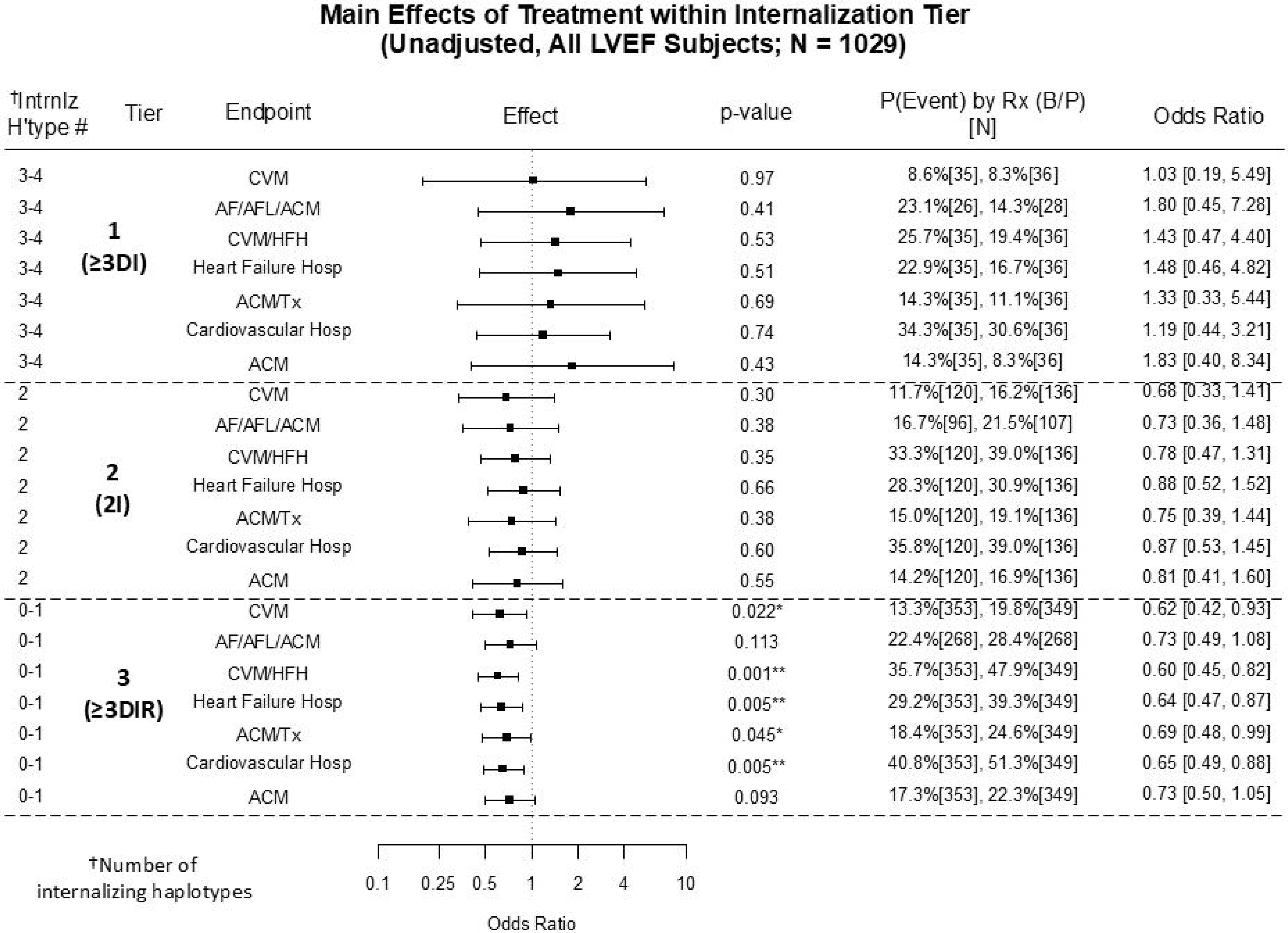
All LVEF cohort subjects, forest plot of bucindolol vs. placebo treatment effect (bucindolol event rate vs. placebo event rate) Odds Ratios (ORs) by internalizing haplotype number in the Column 1 *ADRB1+ADRB2* combination diplotypes. [Ns] are the number of subjects in the bucindolol and placebo groups respectively; otherwise set-up is as in Figures 1 and 2.

#### Placebo Treatment in Subjects with {ADRB1+ADRB2} Internalizing vs. Internalization-resistant Haplotypes is Associated with Lower Clinical Event Rates

In the All-LVEF cohort the endpoint analysis for placebo treatment (**Figure 1**) indicates that subjects within **Tier 1** (≥**3DI)** have OR point estimates to the left of the line of identity, ranging from 0.29 (0.12, 0.68) for cardiovascular mortality or heart failure hospitalization (CVM/HFH) to 0.48 (0.23, 1.00) for cardiovascular hospitalization (CVH). The mean for all 7 endpoints is 0.39 (0.27, 0.55). In contrast, for the **Tier 3** (≥**3DIR)** subjects ORs are to the right of the identity line, between 1.45 (0.88, 2.39) for CVM and 1.78 (1.22, 2.58) for CVH, mean 1.64 (1.46, 1.84). The **Tier 2** (**2I),** subjects position between **Tiers 1** and **3**, slightly to the left of identity, with a mean OR of 0.76 (0.66, 0.88). By an aggregate analysis of all 7 endpoints, **Tier 1** event rates and their ORs are significantly different from **Tier 3** (P=0.005). The less severe LV dysfunction cohort (LVEF ≥0.20) placebo forest plot (**Figure S1**) is very similar to the All-LVEF cohort, with a Tier 1 vs Tier 3 aggregate OR difference also P=0.005. The **Tier 1** mean OR for the 7 endpoints was 0.28 (0.10, 0.82), with a **Tier 3** of 2.16 (1.81, 2.58).

The SEM analysis in the All-LVEF cohort, adjusted for both dependencies between endpoints and direct effects of 13 covariates, confirms an increase in the placebo event rate slopes progressing from 0 to 4 internalization-resistant haplotypes in **Tier 1** through **Tier 3 (Table S7)**. All 7 endpoints in both cohorts have positive individual slopes, indicating increases in event likelihood as the number of internalization-resistant haplotypes is increased. Five of the 7 endpoints in the All-LVEF cohort satisfy P <0.05, with 4 of 7 in LVEF ≥0.20. By Likelihood Ratio Testing the slopes show no evidence of heterogeneity (χ^2^ P =0.58, 0.78 for All-LVEF, LVEF ≥0.20 respectively), indicating that the effect of internalization does not differ across endpoints. A follow-up SEM with equality constraints for this component of the structural model yields a significant positive effect of internalization tier on event likelihood across all 7 endpoints (P = 0.002 and 0.022 for the All-LVEF and ≥0.20 cohorts, respectively) (**Table S7**).

#### Bucindolol Treatment is Unaffected by Haplotype Internalizing Potential

In contrast to placebo treatment, the bucindolol forest plot OR point estimates for the All-LVEF (**Figure 2**) and LVEF ≥0.20 cohorts (**Figure S2)** exhibit minimal tendency for departure from the line of identity, with a nonsignificant SEM analysis (**Table S7**). In contrast to placebo, no estimated effect of increasing internalization-resistant haplotypes is statistically significant (**Table S7)**.

#### Bucindolol vs. Placebo Treatment Effects are Inversely Related to Haplotype Internalizing Potential

Figures 3 and **S3** contain between-treatment group ORs within internalization haplotype tiers, and demonstrate a pattern commensurate with the within-placebo and -bucindolol subjects in Figures 2 and **S2**. In both the All-LVEF (Figure 3) and LVEF ≥0.20 (**Figure S3**) cohorts the ≥**3DI Tier 1** ORs, although statistically nonsignificant, are to the right of the identity line, while in **Tier 3** the ≥**3DIR** ORs are <1.0 with the majority (5 of 7 in Figure 3, all 7 in **Figure S3**) statistically significant. The mean OR of the 7 endpoints for the All-LVEF cohort in **Tier 3** is 0.66 (0.58,0.76), while the LVEF ≥0.20 cohort is 0.49 (0.41, 0.58).

The SEM analysis (**Table S7**) difference between the placebo and bucindolol group slopes indicates consistent treatment effects across all endpoints in both LVEF cohorts. Bucindolol minus placebo slope values vary from −0.11 (HFH) to −0.27 (ACM/Tx) in the All LVEF Cohort, and −0.22 (AF/AFL/ACM) to −0.38 (ACM/Tx) in LVEF ≥0.20, indicting the bucindolol treatment effect is consistently against the presence of internalizing and in favor of internalization-resistant haplotypes. The All 7 Endpoints interaction P values are statistically significant in both cohorts, P=0.048 (All LVEF) and P=0.003 (LVEF ≥0.20), respectively indicating the slopes of the placebo and bucindolol groups are different (**Table S7**), and that bucindolol treatment negates the deleterious effects of increasing numbers of internalization-resistant haplotypes observed in placebo-treated subjects.

#### ADRB1 and ADRB2 Haplotypes Are Associated with Similar Internalization to Non-internalization Dose-Responses Placebo Treatment

**Figures S4-S6** contain All-LVEF cohort data for the internalizing *ADRB1* Gly49Arg389 haplotype held constant at 1 or 2 copies and paired with decreasing copies (from 2 to 0) of the *ADRB2* internalizing, Gln27/Arg16 haplotype. ORs are calculated and plotted as for Figures 1-3, representing an internalizing to internalization-resistant *ADRB2* haplotype dose-response for the 7 clinical endpoints. Within each clinical endpoint there is a progressive increase in ORs for all 7 endpoints as the copy number of *ADRB2* Arg16Gln27 decreases, from an OR range of 0.15 − 0.47 (mean 0.28±0.12) for 2 copy to 1.03 − 1.59 (mean 1.34±0.19) for 0 copies (P=0.016). For the 7 clinical endpoints, 8 of the 14 ORs in the Arg16Gln27 2 or 1-2 copy groups are statistically significantly reduced (upper bound <1.00), with none changed in the remaining 3 categories with fewer copy numbers. The converse, haplotype downtitration of *ADRB1* Gly49Arg389 in the presence of constant, 1-2 copies of *ADRB2* Arg16Gln27 (**Figure S7)** yields similar data.

#### Bucindolol Treatment

In contrast, the internalizing to internalization-resistant haplotype dose-response is not present in patients treated with bucindolol (**Figures S5, S8)**. In **Figure S5,** with downtitration of the internalizing *ADRB2* haplotype Arg16Gln27, ORs are on either side and close to the 1.0 identity line with none statistically significant. The downtitration of the *ADRB1* internalizing haplotype (**Figure S8**) reveals a slightly different pattern, with 2 copies of Gly49Arg389 demonstrating a lower OR (ranging from 0 to 0.73, mean 0.32±0.29) than all other copies in all 7 endpoints. Also, for mortality dominant endpoints (CVM, ACM/Tx, ACM), haplotypes with 0 copies of Gly49Arg389 have ORs consistently >1.0 (range 1.25-1.68, mean 1.52±0.16). These 2 deviations from Arg16Gln27 downtitration are likely due to the respective favorable and relatively unfavorable impacts of bucindolol in subjects with *ADRB1* Arg389Arg and Gly389 genotypes [29].

#### Bucindolol vs. Placebo Treatment Effects

Bucindolol vs. placebo treatment effects with downtitration of internalizing haplotypes (**Figures S6** and **S9**) generally indicate an overall progressive decrease in ORs in all endpoints, or the opposite pattern to that for placebo in **Figures S4** and **S7**. The one exception is consistently low ORs for 2 copy Gly49Arg389 in all 7 endpoints, again likely due to the favorable impact of the *ADRB1* Arg389Arg genotype on bucindolol treatment effects [30].

#### Hazard Ratios for Time to Clinical Events, Internalizing vs. Internalization-resistant Haplotypes Placebo or Bucindolol Treatment in Internalizing vs. Internalization-resistant Haplotypes

Clinical event hazard ratios (HzRs) differ from odds ratios by incorporating the element of time, which can increase the sensitivity for detection of treatment effect differences. For placebo treated patients in the BEST substudy All-LVEF cohort, **Figure S10** contains time to first event curves and HzRs comparing ≥2 Double Internalizing (≥**2DI**, 1-2 copies of *ADRB1* Arg389Gly16 AND 1-2 copies of *ADRB2* Arg16Gln27) to subjects with all other (≥2 Not Double Internalizing, ≥**2IR**) haplotypes, for 4 of the clinical endpoint ORs plotted in Figures 1-3 and **S1-S3**. In all 4 endpoints placebo-treated subjects with ≥**2DI** haplotypes exhibit progressive, favorable separation in the Kaplan-Meier (K-M) curves compared to ≥**2IR** haplotypes, with HzRs ranging from 0.62 (0.40, 0.97) to 0.69 (0.48, 1.00). **Figure S11** contains the same comparisons for bucindolol-treated patients in the All-LVEF cohort, where curve separation does not occur for ≥**2DI** vs. ≥**2IR**subjects; HzRs are near 1.0 for all 4 endpoints, and event rates are similar to the ≥**2DI** placebo curves in **Figure S10**.

#### Bucindolol vs. Placebo Treatment Effects in Maximum Internalization-resistant Haplotypes

As an extreme evaluation of the consequences of internalization-resistant *ADRB1* and *ADRB2* haplotypes on treatment effects, in the LVEF ≥0.20 cohort Figure 4 shows bucindolol vs. placebo time to first event curves for “Maximum Double Internalization-resistant” (**4DIR**) *ADRB1* and *ADRB2* combination diplotypes (2 copies of an *ADRB1* Ser49 containing haplotype AND 2 copies of an *ADRB2* non-Arg16Gln27 diplotype). HzRs (95% C.I.s) range from 0.17 (0.05, 0.56) for ACM/Tx (substudy primary endpoint) to 0.53 (0.33, 0.84) for CVH.

**Figure 4.**
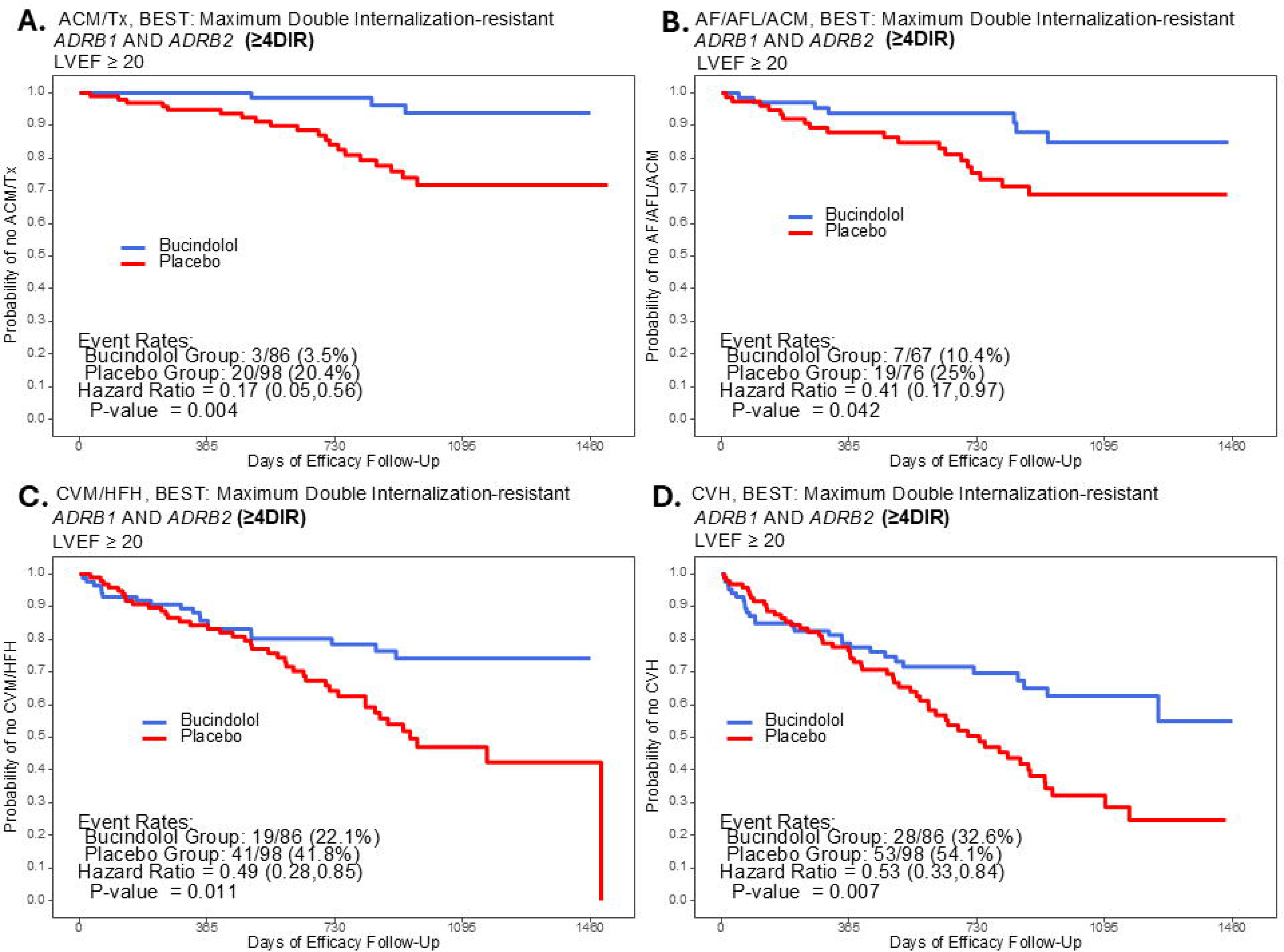
Time to first event curves in bucindolol vs. placebo treated patients with Maximum Double Internalization-resistant (**4DIR**), total of 4 internalization-resistant haplotypes), LVEF ≥0.20 cohort N=184. **A.** All-cause Mortality or Cardiac Transplantation (ACM/Tx); **B.** Heart Failure Hospitalization (HFH); **C.** Cardiovascular Mortality or Heart Failure Hospitalization (CVM/HFH); **D.** Cardiovascular Hospitalization (CVH).

#### ADRB1 and ADRB2 Haplotype Effects Are Equivalent in Black and Non-Black Race

As shown in **Tables 1** and **S1**, Black vs. other races exhibits differences in common genetic variation within β-ARs, as in other neurohumoral [44] and non-neurohumoral systems [45]. Figure 5 consists of bucindolol vs. placebo time to event curves in the All-LVEF cohort by Black or non-Black racial categories for the high prevalence composite endpoints of time to first CVM/HFH and CVH, in subjects with Double Internalizing (≥**2DI)**, and Not Double Internalizing (≥**2IR)** haplotypes. For ≥**2DI** subjects due to favorable effects in placebo-treated subjects there is no evidence of a bucindolol treatment effect in either Blacks or non-Blacks. In contrast, in ≥**2IR** subjects for both endpoints there is evidence of a treatment effect, with little or no difference in HzRs between Black and non-Black subjects (HzRs ranging from 0.64 (0.39, 1.06) for CVH in Blacks to 0.76 (0.61, 0.96) for CVH in non-Blacks, nearly identical HzRs in CVM/HFH). These data indicate that within the ≥**2IR** haplotype phenotype there are no differences between Blacks and non-Blacks for enhancement of bucindolol treatment effects.

**Fig 5.**
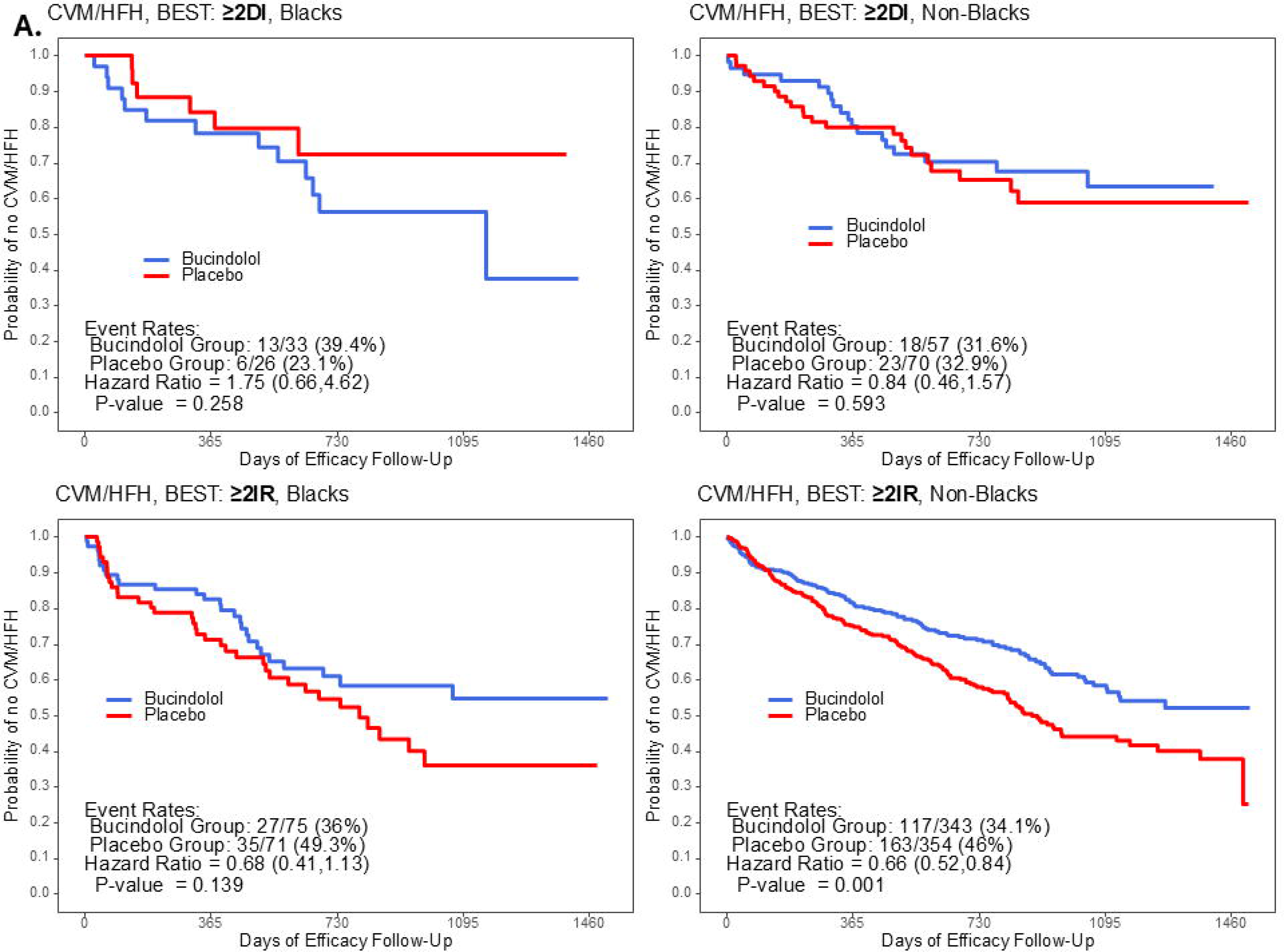
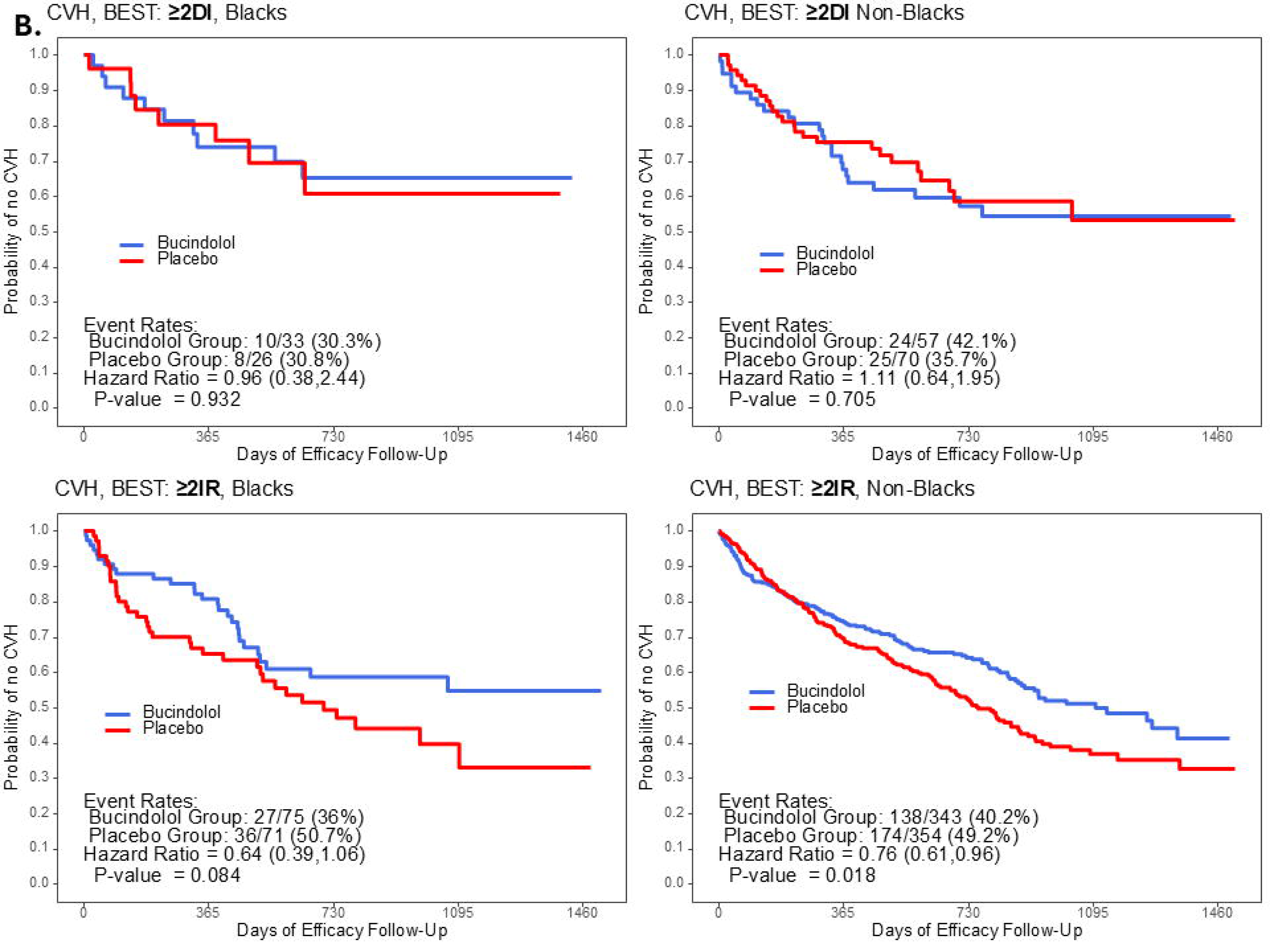
Time to first event curves curves in Blacks or non-Blacks in the All LVEF cohort for the CVM/HFH (**A**) and CVH (**B**) time to first event endpoints, with haplotype categories of: Double Internalizing (≥2**DI**) = ≥1 *ADRB1* AND *ADRB2* internalizing haplotypes (*ADRB1* Gly49Arg389, *ADRB2* Arg16Gln27); Double internalization-resistant (≥**2IR**) = all other haplotypes. The ≥2DI subpopulation is 18.4% of subjects treated ith placebo.

The haplotype dose response progressing from ≥2 Double Internalizing to ≥2 Internalization-resistant in placebo treated patients (Figures 1**, S1**) is observed in both Blacks and non-Blacks (**Figure S12**). For both groups the 7 endpoint ORs in **Tier 1**, ≥**3DI** subjects are consistently to the left of the line of identity, **Tier 3**, ≥**3DIR** point estimates are to the right of 1.0, and **2I** Incompletely Internalizing estimates are intermediate, positioned around 1.0.

### GENETIC-AF DNA Substudy

#### Internalizing/Internalization-resistant Haplotypes

GENETIC-AF Substudy subjects baseline characteristics by *ADRB1* or *ADRB2* diplotype are in **Table S8**. The small sample size precluded assessment of clinical events, but the trial included peripheral venous plasma NT-proBNP and NE biomarker measurements at baseline (pre-randomization to bucindolol or metoprolol succinate) and at 4, 12 and 24 weeks (trial end) of efficacy follow-up [34]. The genotypes, haplotypes and diplotypes are in **Table S9.** Subject diplotypes were divided by {*ADRB1+ADRB2*} diplotype combination into ≥**2DI** and ≥**2IR** groups as for the BEST substudy, and biomarker changes were assessed in the 2 treatment groups (Figure 6, **Tables S10-S13**). Biomarkers were also analyzed in **4DIR** haplotype subjects (**Tables S10-S13**). Neither biomarker was normally distributed, and both were analyzed by non-parametric tests.

**Fig 6.**
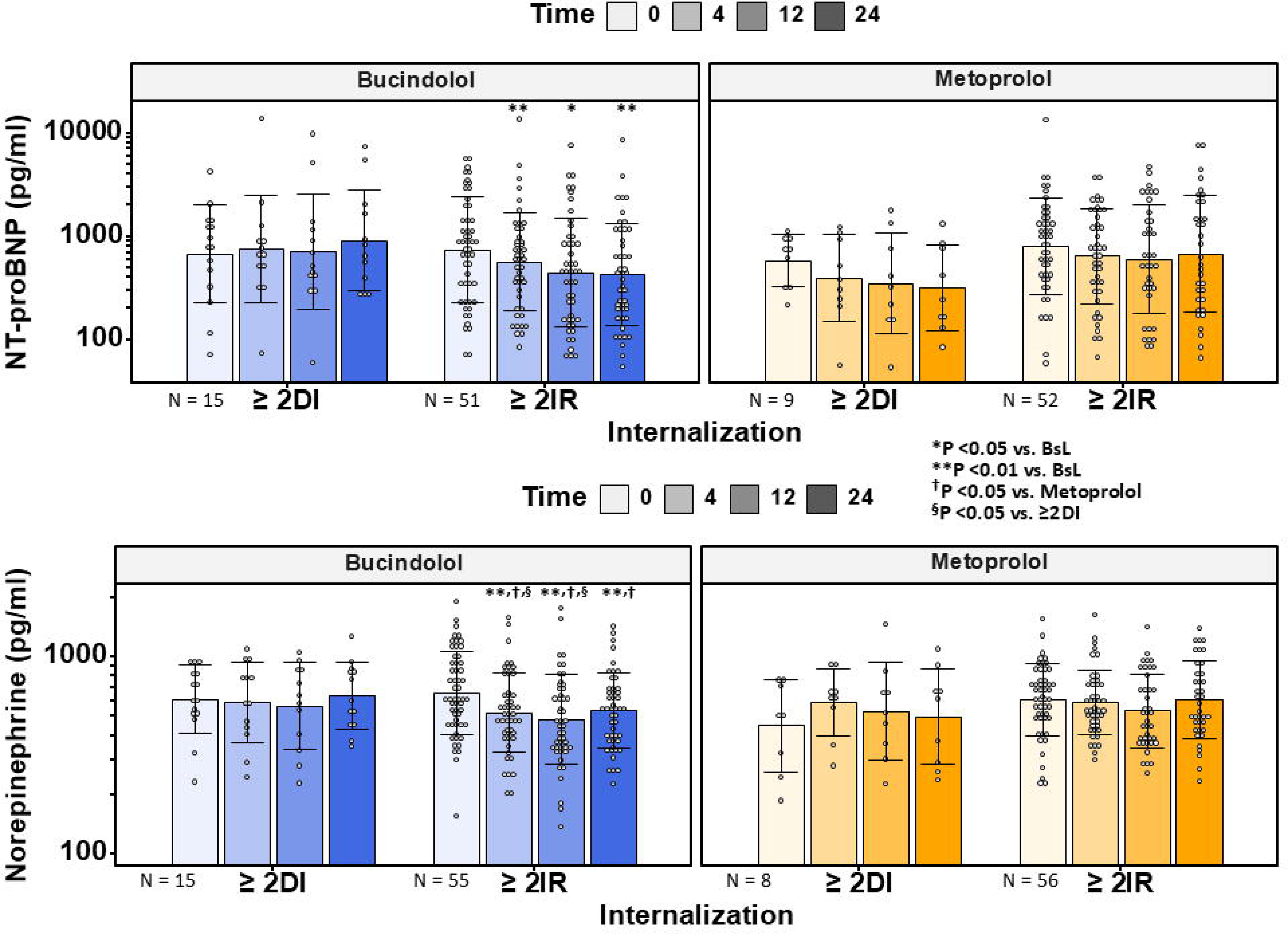
GENETIC-AF study, peripheral venous plasma levels of NT-proBNP (top panel) or norepinephrine (bottom panel) for subjects with ≥2 double internalizing *ADRB1* AND *ADRB2* haplotypes (≥**2DI**) or ≥2 Internalization-resistant haplotypes (≥**2IR**) in the Bucindolol or Metoprolol Groups. Central tendencies of data are in mean±SD, with non-parametric tests used for determining statistical significance.

Baseline characteristics within *ADRB1* or *ADRB2* diplotype cohorts (**Table S8**) or in the treatment arms do not differ meaningfully. NT-proBNP is a biomarker for both AF and HF [34], and thus is a suitable surrogate for GENETIC-AF’s primary endpoint of time to symptomatic AF/AFL/ACM in a HF population. In bucindolol-treated subjects with maximum internalization-resistant, **4DIR** haplotypes (**Table S10**), at 4, 12 and 24 weeks NT-proBNP is decreased by respective amounts of 737±312 pg/ml (mean±sem, P= 0.009), 493±329 pg/ml (P=0.15) and 778±266 pg/ml (P= 0.005). In contrast, metoprolol treated **4DIR** patients (**Table S11**) exhibit no statistically significant changes at any timepoint.For *ADRB1+ADRB2* ≥**2DI** haplotypes, neither bucindolol nor metoprolol treated subjects exhibit changes in NT-proBNP at any timepoint (Figure 6). In contrast, in ≥**2IR** haplotype subjects treated with bucindolol NT-proBNP is reduced from a baseline of 1364±1494 pg/ml by 252±242 pg/ml (mean±sem, P=0.004 vs. baseline), 361±158 pg/ml (P=0.043), and 380±143 pg/ml (P=0.003) at 4, 12 and 24 weeks respectively (Figure 6). Metoprolol-treated patients with ≥**2IR** haplotypes have no significant changes from baseline (Figure 6).

NE measurements are in Figure 6 and in **Tables S12** and **S13**. For bucindolol-treated subjects with **4DIR** haplotypes the pattern is similar to NT-proBNP; there are statistically significant reductions at 4, 12 and 24 weeks (**Table S12**). Also similar to NT-proBNP, in **4DIR** haplotypes there are no reductions in NE at any timepoint in the Metoprolol Arm (**Table S13**). Results for ≥**2DI**, Double Internalizing haplotypes are also similar to NT-proBNP, with no significant changes at any timepoint in either the bucindolol or metoprolol arms (Figure 6)). The ≥**2IR,** bucindolol group (Figure 6) results are also similar to those for NT-proBNP, with statistically significant reductions at 4, 12 and 24 weeks. In the metoprolol arm ≥**2IR** subjects there are no changes in NE at any timepoint. (Figure 6).

### Expansion of the Bucindolol Pharmacogenetic Target Population and Efficacy by the Inclusion of *ADRB1*, *ADRB2* Internalization-resistant Haplotypes

For bucindolol vs. placebo treatment effects, efficacy enhancement by internalization-resistant β-AR haplotypes in the BEST substudy creates the potential for enlarging bucindolol’s pharmacogenetically targeted population beyond *ADRB1* Arg389Arg [29, 34]. Also, within the expanded target population there is the potential for subgroups yielding treatment effects superior to *ADRB1* Arg389Arg [29, 34].

For the LVEF ≥0.20 cohort, treatment effects for all listed pharmacogenetic groups (**Table S15, columns 2-8**) exceed that for the All-Genotypes parent index cohort (Column 1), at a significance level of at least P <0.05. In addition, in 4 of the 6 internalization-resistant diplotype-containing subgroups (Columns 3-8) treatment effects statistically exceed those of the Column 2 *ADRB1* Arg389Arg genotype index pharmacogenetic enhancement subgroup, and all 6 are quantitatively greater. Moreover, the combined *ADRB1* AND *ADRB2* Maximum Double Internalization-resistant (**4DIR**) diplotype subgroup (Column 6) treatment effect exceeds the Column 2 *ADRB1* Arg389Arg group by 1.34 fold (P=0.0017) and the *ADRB1* Arg389Arg group by 1.68 fold (P <0.01) (**Table S15**, **Figure S13**). The combination of the **4DIR** subgroup with non-overlapping *ADRB1* Arg389Arg genotype subjects (Column 7) creates a subgroup that is 33% larger than the Column 2 index pharmacogenetic group (N=441 vs. 331), with a 1.03 fold marginally higher (P=0.064) treatment effect on the paired endpoints analysis. Finally, the requirement that both the *ADRB1* Ser49Arg389 homozygous diplotype AND internalization-resistant *ADRB2* diplotypes be present (Column 8, 10% of the total cohort, the *ADRB1* Arg389 Ser49 homozygous diplotype subset of 4DIR) creates an increase in treatment effect of 1.52 fold >Column 2 (P=0.004), and 1.92 fold >Column1 (P=0.01) (**Figure S13**). Analogous data for the All LVEF cohort are similar (**Table S16**).

### ERK1/2 Phosphorylation in Human Isolated Right Ventricular Trabeculae

In order to confirm that bucindolol acts as a biased ligand in the human heart and is capable of activating a putative cardioprotective pathway, ERK1/2 phosphorylation (pERK1/2) was assessed in preparations of right ventricular (RV) trabeculae isolated from 6 human explanted failing hearts, 5 of which had ≥3 copies of {*ADRB1+ADRB2*} (**Table S17**). After a 5-(N=1) or 10-minute (N=5) tissue bath incubation, bucindolol (P=0.0027 vs. Vehicle) but not metoprolol (P=0.38) is associated with an increase in pERK1/2 compared to vehicle (Figure 7**, Table S17**). Isoproterenol’s pERK1/2/total ERK is similar to bucindolol’s (respectively 1.36±0.36 vs. 1.40±0.27, P=0.73) (**Table S17**) but due to greater variability (Figure 7B) it is not statistically significant (P=0.104 vs. Vehicle). At 60 minutes bucindolol uniquely is decreased compared to values at 5/10 minutes, 0.87±0.55 vs. 1.40±0.27, P=0.037 for interaction with time.

**Figure 7.**
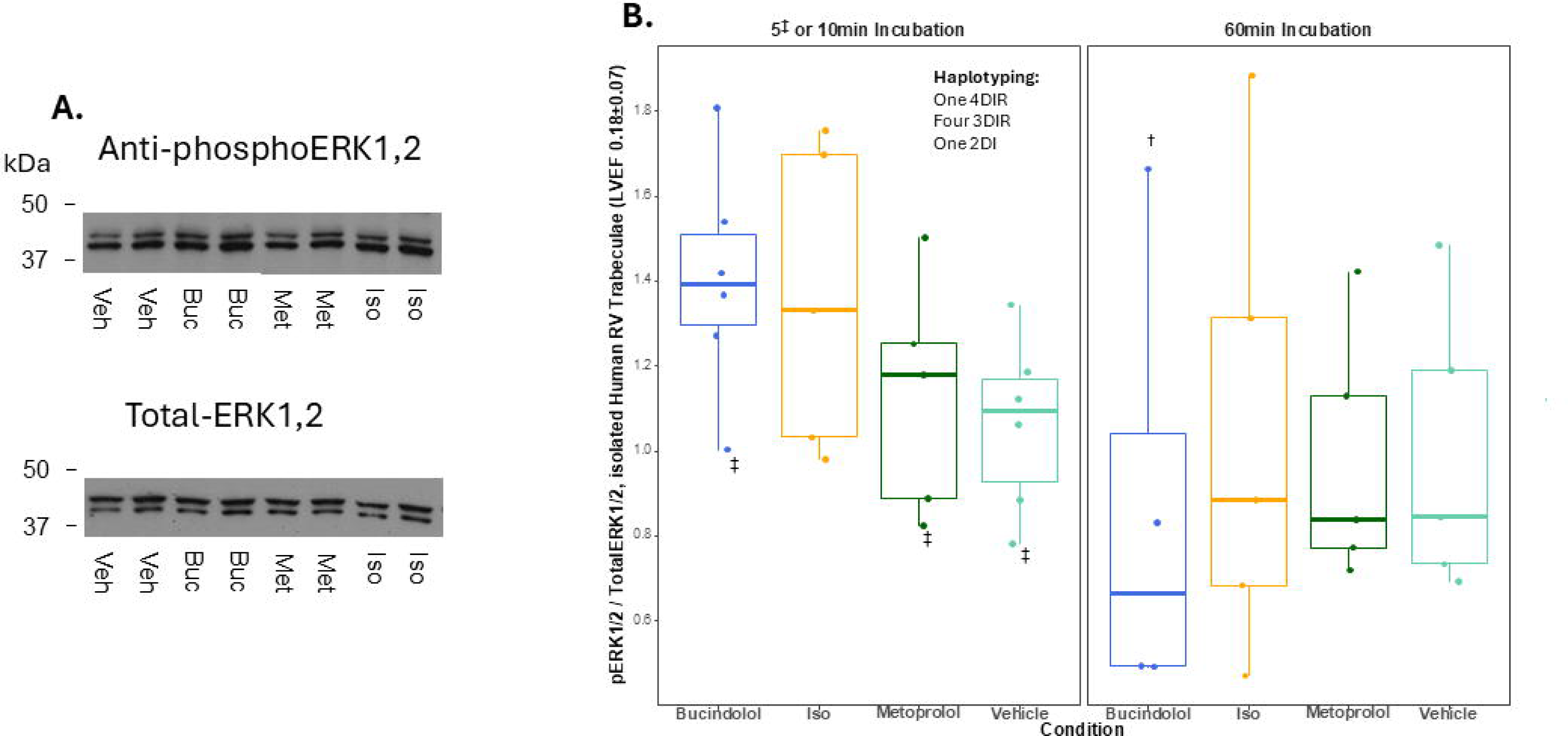
A. ERK1/2 phosphorylation (pERK) and total ERK in preparations of right ventricular trabeculae incubated for 10 minutes with vehicle (Veh); bucindolol (Buc, 1e-6M); metoprolol (Met, 1e-5M); or isoproterenol (Iso, 1e-6M) (patient 5 in Table S17). **B**. Box plots, median and 25, 75 percentile plus individual values of pERK1/2/total ERK1/2. L-panel, Data at 5 (N=1) and 10 minutes incubation (N=5); R-panel, 60 minutes incubation. Drug concentrations are bucindolol 1e-6M, metoprolol 1e-5M and isoproterenol 1e-6M. Vehicle is H_2_0. Data are normally distributed by the Anderson-Darling test; statistical analysis (Table S17) is by ANOVA/mixed model with repeat measures and a Holm-Sidak’s multiple comparison test compared to Vehicle control. *P=0.0027; ^†^P=0.037 for lower value compared to 5/10 minutes (min) and interaction with time; ^‡^5 min incubation, all other points are 10 min.

## DISCUSSION

For same-allele nonsynonymous SNPs it is the encoded protein haplotype encompassing each polymorphic locus that has the potential to modify biologic effects [36−38, 46]. This has been previously shown for *ADRB2* haplotypes expressed in COS-7 cells, where mRNA, protein and pharmacologic phenotype are haplotype encoded [47]. The *ADRB1* 49/389 haplotype would be expected to encode a higher function β_1_-AR that easily internalizes on agonist exposure (Gly49Arg389), a higher function receptor relatively resistant to internalization (Ser49Arg389), and a lower function receptor resistant to internalization (Ser49Gly389) [27]. The 4th possibility, Gly49Gly389, would theoretically have lower function and would easily internalize, but was not found in this and most other genotyping studies [27, 48], presumably due to strong linkage disequilibrium. The β_2_-AR has 2 loci that in tandem regulate internalization, with the Arg16Gln27 haplotype exhibiting a high level of agonist mediated internalization [9, 23−25] and/or rapid desensitization [22, 23]. Similar to *ADRB1*, only 3 *ADRB2* haplotypes are usually found in genetic studies including in the current report, with Arg16Glu27 absent. The absence of 1 of 4 possible haplotypes in each β-AR gene allows near certain maximum likelihood haplotyping from genotype data, and long-read DNA sequencing confirmed that for both *ADRB1* and *ADRB2* double heterozygote genotypes only 1 expected haplotype was present (respectively Gly49Arg389/Ser49Gly389 and Arg16Gln27/Gly16Glu27).

### *ADRB1*, *ADRB2* Haplotypes in the BEST Adrenergic Receptor Polymorphism Substudy

The data from the BEST Adrenergic Receptor Polymorphism substudy indicated that placebo treatment results in reduced clinical event rates in patients who have *ADRB1* and *ADRB2* haplotypes encoding internalizing receptor variants. In a HF patient treated with placebo internalization would presumably occur on exposure to increased NE levels, present in both clinical trial populations. In the BEST substudy, across 7 clinical endpoints placebo-treated subjects with ≥3DI internalizing haplotypes had lower clinical event rates with ORs <1.0 compared to counterpart haplotypes, whereas patients with internalization-resistant (≥3DIR) haplotypes exhibited higher event rates with ORs >1.0. These results are consistent with data from the MAP-MI study, with those previous observations extended to the combination of {*ADRB1+ADRB2*} internalizing haplotypes.

In contrast, subjects in the bucindolol group exhibited more uniform effects across internalization tiers, with event rates similar to placebo in internalizing haplotypes and no statistically significant event rate differences in ORs between haplotype tiers. As a result, on unadjusted analyses bucindolol vs. placebo treatment effect event rate ORs were lower in subjects with internalization-resistant (≥3DIR) haplotypes and trended lower in Incompletely Internalizing (2I) subjects. Structural equation modeling of 7 clinical endpoint events progressing from 0 to 4 internalization-resistant haplotypes confirmed the effects suggested by the unadjusted analyses, as placebo-treated subjects exhibited an increase in event rate slopes and bucindolol treatment was associated with no changes. These differences led to bucindolol vs. placebo treatment effects consistently favoring internalization-resistant haplotypes. The event rate OR data were further corroborated by time to clinical event analyses, where placebo treatment resulted in a reduction in HzRs in subjects with internalizing (≥2DI) compared to internalization-resistant (≥2IR) haplotypes. In contrast, for bucindolol treatment there was no difference between subjects with ≥2DI vs. ≥2IR haplotypes, with HzRs similar to placebo in the ≥2DI haplotype group. That is, when there is at least 1 internalizing haplotype for *ADRB1* AND *ADRB2*, placebo in the presence of a β-agonist (NE) has a favorable effect approximately equivalent to bucindolol. With bucindolol treatment the effects of NE on receptor internalization in subjects with ≥2DI, internalizing haplotypes would presumably be blocked, with biased ligand effects leading to internalization/and or blockade of any adverse effects mediated through canonical cell surface signaling. In contrast, in subjects with ≥2IR, internalization-resistant haplotypes, bucindolol exhibited efficacy in excess of placebo with statistically significant treatment effects ranging from 24% to 36% for time to effect analyses regardless of race. The therapeutic preference of bucindolol for internalization-resistant haplotypes was underscored by results in 4DIR subjects, who exhibited treatment effects from 51% to 83%. Therefore, compared to placebo, the biased ligand β-blocker bucindolol was associated with a more favorable treatment effect in subjects with internalization-resistant haplotypes, which constituted approximately 70% of the BEST substudy All LVEF cohort.

### *ADRB1*, *ADRB2* Haplotypes in the GENETIC-AF Trial

The limited sample size of the GENETIC-AF DNA substudy was nevertheless sufficient to detect β-AR haplotype effects on NT-proBNP and NE. Both NT-proBNP and NE plasma levels were reduced by bucindolol compared to metoprolol, similar to results in the entire cohort [34, 49]. In the substudy the reductions in both biomarkers occurred only in the Bucindolol Arm in subjects with 4DIR or ≥2IR internalization-resistant haplotypes. In contrast, metoprolol succinate was not associated with a reduction in NT-proBNP or NE in subjects with internalization-resistant haplotypes. Thus relative to internalization category haplotypes the effect of the nonbiased antagonist metoprolol was different from bucindolol, and from placebo in the BEST substudy where clinical events were reduced in ≥2DI subjects. This suggests that in subjects with ≥2DI, internalizing haplotypes metoprolol blocked the beneficial, receptor internalizing effects of NE, but did not produce any favorable effects on its own.

This could be because metoprolol is a nonbiased ligand, or because as β_1_-AR selective antagonist β_2_-AR adverse canonical signaling was unaffected.

### Haplotype Data Indicate that Both *ADRB1* and *ADRB2* Internalizing Variants Affect Heart Failure Clinical Events and Bucindolol Treatment Effects

Human cardiac myocytes contain β_2_-ARs that are coupled to positive inotropic effects via the canonical cAMP/PKA pathway [1] and to ERK1/2 signaling following internalization [10]. When overexpressed in mouse hearts human β_2_-ARs mediate histopathological effects [2, 51], and in failing human ventricles the proportion of β_2_-ARs/Total β-ARs is increased, from 20-25% to 35-40% [1].

However, the numerically dominant β_1_-AR signaling pathway is more biologically adverse, and in clinical trials mortality reduction outcomes are similar for β_1_-AR selective blocking agents and the nonselective, biased ligand β-blocker carvedilol [1]. Despite these observations, in the current study with placebo treatment *ADRB2* internalizing haplotypes were associated with improved clinical outcomes, and internalization-resistant haplotypes conferred improved bucindolol treatment effects. Furthermore, haplotype copy number dose-response indicated that *ADRB2* effects were equivalent to those for *ADRB1* haplotypes. These data support that in the human heart myocardial β_2_-ARs are operational in the natural history of adrenergically-mediated pathologic ventricular remodeling, and that their pharmacologic manipulation can have therapeutic consequences. Moreover, for clinical endpoints the combination of *ADRB1* and *ADRB2* internalization-resistant haplotypes was additive, as might be expected based on the known differences in their encoded receptor signaling pathways [1].

### Effects of Race

As it does for the *ADRB1* Arg389Gly polymorphism [44], race also affected β-AR internalization variants, and was the only baseline characteristic in the BEST adrenergic receptor polymorphism substudy that consistently varied by diplotype. Blacks compared to non-Blacks had higher frequencies of internalizing and lower frequencies of internalization-resistant haplotypes, which suggests that in the absence of other confounding variables placebo treatment (or presumably no treatment) would favor Blacks based on the beneficial impact of a greater frequency of internalizing *ADRB1* and *ADRB2* haplotypes. In contrast, some of bucindolol’s efficacy enhancement is dependent on the presence of internalization-resistant haplotype encoded receptors, for which non-Blacks had a slight advantage.

However, as shown in the current and a previous genotype study [44], within the same haplotypes Blacks have therapeutic responses to bucindolol that are equivalent to those in non-Blacks. These observations underscore the value of pharmacogenetic targeting in predicting drug response, which can override any contribution by baseline characteristics such as race or other potentially confounding variables.

### Biased Ligand Properties of Bucindolol

A biased β-adrenergic receptor ligand can be defined as a compound that can signal through both G-protein dependent and independent pathways [52, 53], favoring one or the other in certain contexts [47]. Cardiac myocyte surface membrane β_1_-or β_2_-ARs typically signal through the G-protein-cAMP/PKA or CaMKII pathway [1], and internalized receptors [8, 10, 12, 30−32] through EGFR/ERK1/2. As it applies to bucindolol or the structurally similar compound carvedilol [53], in systems that are hypersensitive for the detection of G protein mediated adenylate cyclase stimulation/cAMP generation both compounds exhibit weak β-agonist activity, but also can activate MAP kinase ERK1/2 independent of G-proteins [13] qualifying them as biased ligands. For both compounds the β-agonist activity is insufficient for activation of the β_1_-or β_2_-AR pathway in the human heart either in vitro [29, 54, 55] or in vivo [56, 57]; data from the current study indicate that in failing human hearts bucindolol is capable of activating ERK1/2 signaling, further qualifying it as a biased ligand.

A comparison of factors influencing the biased ligand properties of bucindolol and carvedilol includes: similar ligand binding characteristics that differ from other β-AR antagonists when co-crystallized with the avian β_1_-AR [53]; higher potency of bucindolol for ERK1/2 activation [14]; similar guanine nucleotide modulatable binding (a characteristic of agonists) in human β_1_- and β_2_-ARs [54, 55]; and for bucindolol but not carvedilol, following 24 hours of incubation downregulation of β_1_- and β_2_-ARs with no reduction in cognate mRNA [58] (evidence of internalization leading to receptor degradation from trafficking to lysosomes [59, 60]. These latter experiments were performed in both cultured avian cardiac myocytes (primarily β_1_-ARs) and hamster β_2_-AR containing smooth muscle cells that express internalization-resistant β-AR haplotypes [61, 62]. In addition, bucindolol’s or carvedilol’s ERK1/2 activation can be blocked by endocytosis/internalization inhibitors [63].

In the current study the increase in ERK1/2 phosphorylation in response to bucindolol occurred rapidly (after a 10 minute incubation) as noted by others for isoproterenol [12] and nebivolol [17], and converted to dephosphorylation by 60 minutes. The degree of early activation was quantitatively similar to the statistically nonsignificant change for isoproterenol, and the nonbiased β-blocker metoprolol did not activate ERK1/2 signaling. Five of the 6 isolated RV trabeculae preparations were from failing hearts with a majority of internalization-resistant (*ADRB1+ADRB2*) haplotypes (four ≥3DIR and one 4DIR).

These findings support the possibility that the favorable effects of bucindolol in internalization-resistant receptors could have been due to “cardioprotective” signaling mediated either by biased ligand-induced receptor internalization [15−17] or ERK1/2 signaling independent of internalization [64, 65]. That a biased ligand β-blocker can internalize relatively resistant β-AR haplotypes and activate ERK1/2 signaling has been previously established with carvedilol [16], in β_1_-ARs expressed from the original human β_1_-AR cDNA that is Ser49Gly389 [66].

Carvedilol’s interaction with β-AR haplotypes in clinical HF settings has not been investigated. Despite their similarities bucindolol and carvedilol have somewhat different properties, including moderate sympatholysis with bucindolol but not by carvedilol [1, 50] that likely contributes to the former’s enhanced effectiveness in HF patients homozygous for *ADRB1* Arg389 [1, 29, 67, 68]. Unlike bucindolol, carvedilol has not exhibited selective enhancement of transplant-free survival in subjects with *ADRB1* Arg389Arg [69, 70] genotypes. Bucindolol is also a more potent β_2_-AR antagonist than carvedilol, by approximately 5-8 fold [55].

### Haplotype Targeting Expands the Pharmacogenetic Heart Failure or Atrial Fibrillation Prevention Potential of Bucindolol

In BEST substudy subjects with LVEFs ≥0.20 the enhanced treatment effects of bucindolol in subjects with both *ADRB1* and *ADRB2* Maximum Double Internalization-resistant (4DIR) diplotypes (a 25% subgroup) resulted in a 1.34 fold increase in treatment effects compared to the *ADRB1* Arg389Arg index population, and 1.68 fold increase compared to the All-Genotypes cohort. If *ADRB1* Ser49Arg389 homozygous diplotypes AND 4DIR diplotypes were considered (a 10% subgroup) the treatment effect enhancement compared to the index *ADRB1* Arg389Arg group was by 1.52 fold in the LVEF ≥0.20 cohort and by 1.40 fold in the All-LVEFs cohort, with respective increases vs. the All-Genotypes cohort of 1.92 and 2.06 fold. Thus, efficacy enhancement through targeting internalization-resistant *ADRB1* and *ADRB2* haplotypes exceeds that for *ADRB1* Arg389Arg, and when combined with this genotype using AND logic yields an approximate doubling in effectiveness.

When the *ADRB1* AND *ADRB2* ≥4DIR subgroup was combined with *ADRB1* Arg389Arg subjects by OR logic the result was a 60-62% subgroup that was essentially identical to *ADRB1* Arg389Arg for treatment effects, in both LVEF cohorts. As a result, in the BEST pharmacogenomics substudy the addition of internalization-resistant *ADRB1* and *ADRB2* haplotypes (“haplotype targeting”) to the current *ADRB1* Arg389Arg targeting resulted in a larger population eligible for benefit, increasing from 45-47% to 60-62% of the all-genotype parent population. Since the BEST pharmacogenetic substudy was enriched in Blacks, who have a lower frequency of both *ADRB1* Arg389 and internalization-resistant *ADRB1* and *ADRB2* haplotypes, the race-adjusted eligibility for therapeutic enhancement in the entire U.S. population would increase from 49% to 65%. Thus, by using the *ADRB1* Arg389Arg OR (*ADRB1* AND *ADRB2* Internalization-resistant) joint criterion, the population eligible for treatment enhancement can be increased by 33%.

### Heart Failure Natural History Implications

The effects in the placebo-treated group could have implications for HF natural history, which suggest that patients with ≥2 (*ADRB1*+*ADRB2*) internalizing haplotypes have a more favorable prognosis, may not respond any better than placebo to a biased ligand β-blocker and could have worse outcomes when treated with a non-biased β-blocker. Equivalently, in comprising a likely substantial majority of HF patients (approximately 70%), patients with 3 or more internalization-resistant haplotypes constitute a major at-risk population.

### Potential Limitations

These data were generated in 2 previously conducted clinical trials, one of which [35] was from an era when HFrEF therapy did not include two (sacubitril-valsartan and SGLT2 inhibitors) of the current “Four Pillars” [71] of HF drug treatment, and the other [34] was prior to the regulatory approval and therapeutic implementation of SGLT2is. Although the tested hypothesis was generated from observations made in a more recent study [27], further clinical outcomes testing of the β-AR haplotype internalization hypothesis is warranted in contemporary HF populations.

In addition to the biased ligand/receptor internalization/cardioprotective signaling hypothesis there are other possible explanations for the differential effects of apparent receptor internalization treatment associated clinical events. These include non-EGFR/ERK1/2 subcellular signal trafficking [72] of encoded internalized haplotypes, and internalization-based exclusion of cell surface β-ARs from adverse canonical signaling. In the latter paradigm the benefit of internalized, desensitized β-ARs in the presence of placebo is simply to attenuate the adverse effects of cell surface canonical signaling [16, 73], while bucindolol as a high affinity β_1_-, β_2_-AR antagonist [1, 54, 55] exerts a greater therapeutic effect when receptors are non-internalized. These possible mechanistic explanations will need to be further investigated in cardiac myocyte model systems expressing human β-AR haplotypes, using a non-radioligand receptor internalization assay to accommodate the slow β_1_- and β_2_-AR offset kinetics of bucindolol.

### Conclusions

In HFrEF and HFmrEF patients, polymorphic loci in the intronless *ADRB1* and *ADRB2* genes encoding receptor haplotypes that regulate internalization affect HF and AF clinical events as well as biomarker behavior, in both placebo- and bucindolol-treated subjects. Including *ADRB1* and *ADRB2* internalization-resistant haplotypes in bucindolol’s pharmacogenetic targeting results in efficacy enhancement similar to that produced by the *ADRB1* Arg389Arg genotype, and restricting the *ADRB1*contribution to Arg389 in a internalization-resistant diplotype combination produces an additive effect as would be expected by polymorphisms affecting different mechanisms. Haplotype pharmacogenetic targeting is feasible in HF, and can identify populations at differential risk for serious outcomes as well as candidates for increased therapeutic effectiveness.

## Data Availability

Lead contact, MRB materials available, this study did not generate new unique reagents; outputs and code for structural equation modeling and haplotyping data are available on GitHub, Group level data not included in the manuscript are available on request, De-identified subject level data for use in a research context will be available on request

## Acknowledgements

Funding sources: This work was supported by National Heart, Lung, and Blood Institute (NHLBI) Grants HL052318, HL48013, HL071609, and HL077101. Additional support is from Genvara Biopharma, Westminster, CO. DNA samples from the BEST trial were provided by the BEST DNA Bank, co-sponsored by the NHLBI and the Department of Veterans Affairs Cooperative Studies. DNA Samples for the GENETIC-AF trial were provided by ARCA Biopharma (Westminster, CO), the trial’s sponsor. We thank Rachel Rosenberg for manuscript and submission handling.

## Author contributions

Conceptualization, M.R.B., I.A.C., S.B.L.; Methodology, I.A.C., M.R.B., D.S., W.A.M., E.R.J., L.A.W., A.D.K., A.V.A., S.B.L., D.K.; Software, I.A.C.; Validation, M.R.B., I.A.C., W.A.M., D.S.; Formal Analysis, I.A.C., M.R.B., D.S., W.A.M., L.A.W.; A.D.K. Investigation, S.B.L., M.R.B., I.A.C., D.S., E.R.J., W.A.M., L.A.W., D.K., A.D.K.; Resources, M.R.B., S.B.L., I.A.C., A.V.A., T.A.M; Data Curation, I.A.C, M.R.B., D.S.; Writing− Original Draft, M.R.B., I.A.C., D.S., W.A.M.; Writing – Review & Editing, M.R.B., I.A.C., S.B.L., T.A.M.,L.A.W.; Visualization, I.A.C. E.R.J., M.R.B., W.A.M; Supervision, M.R.B., S.B.L., T.A.M, D.S, A.V.A.; Project Administration, M.R.B., S.B.L.; Funding Acquisition, M.R.B., S.B.L.

## Availability of Data and Materials

Outputs and code for structural equation modeling and haplotyping data are available on GitHub (https://github.com/CUAMC-SoM-Cardiology/M_Bristow-Lab). Group level data not included in the manuscript are available on request. De-identified subject level data for use in a research context will be available on request.

