## Supplemental File for "β_1_- and β_2_-adrenergic Receptor Haplotypes Regulate Therapeutic Responses to Placebo and the Biased Ligand β-blocker Bucindolol"

#### SUPPLEMENTAL MATERIAL

##### Conventions and Presentation Formats

Single gene, two loci haplotypes are presented in an allelic orientation as: encoded amino acid-position1-amino acid-position 2, e.g. *ADRB1* Gly49Arg389 in a 5′ to 3′ order). Diplotypes are presented as haplotype 1/haplotype 2, with the amino acid preceding the polymorphism position in either a stacked $\frac{\text{Gly49Arg389}}{\text{Ser49Arg49}}$. or linear configuration, e.g. *ADRB1* or Arg389Gly49/Gly389Ser49. Genotypes are designated by the encoded amino acids flanking their position number, e.g. Arg389Gly, Arg389Arg. Diplotypes are described as homozygous (two identical haplotypes), double heterozygous (different encoded amino acids in each polymorphic locus), or single homozygous (1 position with the same encoded amino acid on both haplotypes, the other position with different amino acids). Polymorphisms are presented with the major allele preceding and the minor allele following the amino acid position, and annotated as a "polymorphism" to differentiate from a heterozygote genotype designation, e.g. "Arg389Gly polymorphism" vs. "Arg389Gly genotype". *ADRB1* and *ADRB2* receptor haplotypes are further classified as either "Internalizing" and "Internalization-resistant" to categorize them as having previously been shown to have respective greater or lesser degrees of internalization or internalization-related pharmacologic or pharmacologic biologic behavior (acute desensitization, effects on receptor trafficking including a decrease in receptor density with no change in mRNA abundance in response to agonist exposure of ≤24 hours).

Clinical events vs. haplotype gene dose-response analyses were assessed by comparing subjects with Internalizing vs. Internalization-resistant *ADRB1* and *ADRB2* variants within diplotypes or *ADRB1* + *ADRB2* diplotype combinations. Because of smaller sample sizes in some subgroups (Internalizing haplotypes, Black race) or genotype restrictions (GENETIC-AF cohort) the definition of internalizing groups varied by the type of analysis, but always consisted of more vs. fewer internalizing haplotypes. As an aid to comparing these analyses, each defined diplotype was assigned a unique descriptor as described in **Table S2.**

#### SUPPLEMENTAL METHODS

##### Forest Plots

Forest plots of clinical event odds ratios (ORs) were used to perform a group-level analysis of events within endpoints in patients with internalizing vs. internalization-resistant haplotypes (Figures 1-3, S1-S3). For graphical efficiency, the ACM/HFH endpoint was not included. Combined {*ADRB1* and *ADRB2*} haplotypes were rank-ordered into 3 mutually exclusive tiers by haplotype internalization potential, progressing from internalizing to internalization-resistant potential. Only first events were assessed, in an unadjusted analysis in each tier's internalizing vs. all other haplotypes. The analyses were performed by treatment group and on the treatment effects of bucindolol vs. placebo, using intention to treat data. For the *ADRB1*/*ADRB2* haplotype titration forest plots (**Figures S4-S9**), constructed for pattern recognition and consistency between clinical endpoints, only the individual comparison OR 95% C.I.s and P values were calculated.

##### Structural Equation Modeling

The second evaluation of clinical events in patients with internalizing vs. internalization-resistant haplotypes was a subject-level analysis with covariate adjustment. These analyses were performed by treatment group, and on the treatment effects in the bucindolol vs. placebo group, using intention to treat data.

The structural equation model ("SEM") for binary outcomes (representing events from each endpoint) used the WLSMV estimator (diagonally-weighted least squares with a full mean- and variance-adjusted weight matrix for robust estimation of standard errors and chi-square based statistics). The readout is event likelihood for each of the seven candidate endpoints simultaneously, regressing binary outcomes onto internalization (0-4 copies of internalization-resistant haplotypes), treatment arm, the interaction of treatment arm and internalization (treatment effect), and the covariates. Inputted covariates are LVEF, Coronary Artery Disease, Sex, Race, Volume Overload, NYHA Class, AF at

Baseline, DNA Lag (time between randomization and blood draw for DNA extraction), duration since HF diagnosis, duration of event free survival, age, baseline systolic blood pressure, and baseline heart rate. All these covariates have been used for stratification in the 2 clinical trials or have been shown to relate to the probability of occurrence of the primary endpoints [34, 35].

Full model readouts are archived on the GitHub site (https://github.com/CUAMC-SoM-Cardiology/M_Bristow-Lab).

**Time to First Event Analyses**

For time to event (Kaplan-Meier) curves all analyses used unadjusted methodologies, as adjustments, particularly in small sample sizes, may yield misleading results. However, for sensitivity purposes we also performed analyses adjusted for randomization stratifiers and other covariates, which were not different from the unadjusted versions and are archived in the GitHub site (https://github.com/CUAMC-SoM-Cardiology/M_Bristow-Lab).

**pERK1/2 Immunoblotting**

Frozen trabeculae (25-35 mg) were homogenized in 25 volumes of a homogenization buffer cocktail (8M Urea, 2.5M thio-Urea, 4% CHAPS, 2mM EDTA, 0.01M dithiothreitol, 1% Protease Inhibitor Cocktail (Sigma P-8340) and 1% ReadyPrep TBP Reducing Agent (BioRad 1632101)) in glass mortar and pestle tissue grinders on ice. Supernatant fractions were saved after a 13,600xg centrifugation, aliquoted and frozen at -80°C. Total protein content of the supernatants was determined by a Bradford assay (Pierce Detergent Compatible 1863028).

Immunoblots were performed on 5 micrograms of protein per sample by SDS-PAGE separation in 10% polyacrylamide gels (BioRad, Mini PROTEAN TGX 4561036), then transferred to 0.2 µm PVDF membranes (BioRad, ImmunBlot 16201777). Primary antibodies for ERK (Cell Signaling 9102), pERK (Cell Signaling 9101) and GAPDH (Santa Cruz 32233) were applied sequentially to the blots with the subsequent appropriate HRP-linked secondary antibody following (Sigma A2304 and A9169). Chemiluminescence was initiated (ThermoScientific SuperSignal West Pico-plus 34580) and captured by exposure to film that was then scanned and imaged by software package (ImageJ, NIH, imageJ.org). Chemical stripping of antibodies from the blot was performed between each primary antibody detection, before application of the next set of antibodies. pERK1/2 density was normalized to its respective unphosphorylated band, and compared to vehicle control at each incubation timepoint.

#### SUPPLEMENTAL TABLES

**Table S1.** Subject characteristics by diplotype, treatment, or Race, (Mean±SD), BEST Adrenergic Receptor Polymorphisms Substudy (N= 1029 *ADRB1*, 1040 *ADRB2*).

| Diplotype, Treatment | N | Age (Yrs) | M/F (%) | †Race | HF Duration (Mos) | BMI (Kg/Ht$^2$) | Hx DM (%) | Hx Htn (%) | NYHA III/IV (%) | LVEF (%) | HR (bpm) | SBP (mmHg) | PNE, pg/ml | AF (%) | ‡VOL OL (%) |
|---|---|---|---|---|---|---|---|---|---|---|---|---|---|---|---|
| ***ADRB1*** | | | | | | | | | | | | | | | |
| Ser49Arg389<br>Ser49Arg389 | 302 | 61<br>±12 | 81/<br>19 | 93/7*** | 48<br>±52 | 28<br>±6 | 33 | 47 | 94/<br>6 | 23<br>±7 | 82<br>±14 | 117<br>±17 | 479<br>±257 | 24 | 36 |
| Gly49Arg389<br>Gly49Arg389 | 38 | 56<br>±14* | 71/<br>29 | 63/37* | 48<br>±45 | 30<br>±6* | 50 | 71 | 95/<br>5 | 23<br>±6 | 82<br>±14 | 121<br>±19 | 382<br>±197** | 24 | 34 |
| Ser49Gly389<br>Ser49Gly389 | 93 | 58<br>±14 | 82/<br>18 | 62/38** | 47<br>±45 | 30<br>±6* | 30 | 63 | 89/<br>11 | 23<br>±7 | 80<br>±15 | 121<br>±16* | 510<br>±349 | 16 | 43 |
| Ser49Arg389<br>Gly49Arg389 | 149 | 60<br>±11 | 79/<br>21 | 80/20 | 47<br>±51 | 28<br>±5 | 38 | 56 | 95/<br>5 | 24<br>±7 | 80<br>±14 | 119<br>±20 | 493<br>±354 | 23 | 40 |
| Gly49Arg389<br>Ser49Gly389 | 101 | 60<br>±12 | 73/<br>27 | 60/40*** | 43<br>±52 | 28<br>±6 | 38 | 62 | 93/<br>7 | 25<br>±7* | 82<br>±12 | 119<br>±20 | 449<br>±226 | 19 | 32 |
| Ser49Arg389<br>Ser49Gly389 | 346 | 61<br>±12 | 80/<br>20 | 81/19 | 43<br>±41 | 28<br>±6 | 34 | 58 | 90/<br>10* | 24<br>±7 | 82<br>±13 | 118<br>±17 | 506<br>±268* | 25 | 32 |
| ***ADRB2*** | | | | | | | | | | | | | | | |
| Gly16Gln27<br>Gly16Gln27 | 70 | 61<br>±10 | 70/<br>30 | 73/27 | 47<br>±50 | 27<br>±5 | 34 | 49 | 94/<br>6 | 25<br>±7* | 80<br>±15 | 119<br>±18 | 452<br>±218 | 17 | 33 |
| Arg16Gln27<br>Arg16Gln27 | 178 | 61<br>±12 | 79/<br>21 | 77/23 | 44<br>±48 | 28<br>±6 | 37 | 54 | 91/<br>9 | 23<br>±7 | 82<br>±13 | 118<br>±18 | 491<br>±273 | 21 | 31 |
| Gly16Glu27<br>Gly16Glu27 | 143 | 61<br>±12 | 78/<br>22 | 95/5*** | 44<br>±43 | 29<br>±6 | 42 | 50 | 93/<br>7 | 23<br>±7 | 80<br>±15 | 118<br>±17 | 490<br>±336 | 27 | 40 |
| Arg16Gln27<br>Gly16Gln27 | 210 | 59<br>±12* | 79/<br>21 | 63/37*** | 49<br>±48 | 29<br>±7 | 34 | 68 | 92/<br>8 | 23<br>±8 | 82<br>±12 | 118<br>±18 | 494<br>±272 | 23 | 37 |
| Arg16Gln27<br>Gly16Glu27 | 279 | 61<br>±12 | 84/<br>16* | 87/13*** | 44<br>±47 | 28<br>±6 | 34 | 52 | 92/<br>8 | 23<br>±7 | 81<br>±13 | 117<br>±18 | 499<br>±305 | 26 | 35 |
| Gly16Gln27<br>Gly16Glu27 | 160 | 60<br>±13 | 79/<br>21 | 82/18 | 45<br>±49 | 29<br>±6 | 28 | 57 | 92/<br>8 | 24<br>±7 | 82<br>±13 | 120<br>±18 | 467<br>±236 | 20 | 36 |
| Black | 207 | 56<br>±13*** | 74/<br>26 | 0/<br>100*** | 46<br>±45 | 29<br>±8 | 39 | 83 | 94/<br>6 | 24<br>±7 | 85<br>±13*** | 121<br>±19** | 502<br>±336 | 14 | 41 |
| Non-Black | 833 | 61<br>±12*** | 81/<br>19 | 100/<br>0*** | 45<br>±48 | 28<br>±5 | 34 | 49 | 92/<br>8 | 24<br>±7 | 80<br>±13*** | 117<br>±18** | 484<br>±270 | 25 | 34 |
| Placebo | 525 | 60<br>±12 | 78/<br>22 | 81/19 | 46<br>±48 | 28<br>±6 | 33 | 54 | 93/<br>7 | 24<br>±7 | 81<br>±13 | 118<br>±18 | 475<br>±276 | 23 | 36 |

| | | | | | | | | | | | | | | |
|---|---|---|---|---|---|---|---|---|---|---|---|---|---|---|
| Bucindolol | 515 | 60 ±12 | 80/ 20 | 79/21 | 45 ±46 | 28 ±6 | 37 | 57 | 92/ 8 | 23 ±7 | 82 ±14 | 118 ±18 | 500 ±289 | 23 | 35 |

*P<0.05, **P<0.01, ***P<0.001 vs. all other members of the same genotype group. †non-Black/Black; ‡VOL OL (Volume overloaded). M/F refers to male or female self-identified gender. HF=heart failure, HxDM = history of diabetes, Hx Htn=history of hypertension, HR=heart rate, SBP=systolic blood pressure, PNE=plasma norepinephrine, AF= atrial fibrillation.

**Table S2.** Combination diplotype terminology based on internalization potential of *ADRB1* and *ADRB2* haplotypes.

| **Cohort, Internalization Term based on number of haplotypes** | **Acronym** | **Rationale** | **Figures, Tables** |
|---|---|---|---|
| **BEST** | | | |
| ≥3 Double* internalizing | ≥3DI | 3-4 internalizing haplotypes; 3DI = 3 internalizing | Figs. 1-3, S1-S3 |
| Incompletely internalizing | 2I | All Non-overlapping intermediates between ≥3DI and ≥3NDI | Figs. 1-3, S1-S3 |
| ≥3 Double* Internalization-resistant | ≥3DIR | 3-4 internalization-resistant haplotypes; 3DIR = 3 internalization-resistant | Figs. 1-3, S1-S3 |
| Maximum Internalization-resistant | 4DIR | All diplotype haplotypes are internalization-resistant | Fig. 4 |
| ≥2 Double* internalizing | ≥2DI | 2-4 internalizing haplotypes; 2DI = at least 1 on each gene | Figs. 5, S10, S11 |
| ≥2 Internalization-resistant ("Not Double Internalizing") | ≥2IR | Haplotypes other than ≥2DI; 2IR = 2 internalization-resistant haplotypes on same gene, 2 internalizing on the other (2 internalization-resistant haplotypes on at least 1 gene) | Fig. 5, S10, S11 |
| **GENETIC-AF** | | | |
| Maximum Internalization-resistant | 4DIR | All diplotype haplotypes are internalization-resistant | Tables S10-S13 |
| ≥2 Double* internalizing | ≥2DI | 2-4 internalizing haplotypes; 2DI = at least 1 on each gene | Fig. 6 |
| ≥2 Internalization-resistant ("Not Double Internalizing") | ≥2IR | Haplotypes other than ≥2DI; 2IR = 2 internalization-resistant haplotypes on same gene, 2 internalizing on the other (2 internalization-resistant haplotypes on at least 1 gene) | Fig. 6 |

*"Double" means present on both *ADRB1* and *ADRB2* genes. †Internalizing haplotypes = *ADRB1* Gly49Arg389, *ADRB2* Arg16Gln27; All other haplotypes are Internalization-resistant.

**Table S3** Minor allele frequencies (MAF) of *ADRB1* and *ADRB2* polymorphisms by Black (B), non-Black (non-B) race; BEST Adrenergic Receptor Polymorphisms Substudy (from genotype data).

| **Gene, amino acid position, SNP reference (rs) number** | **Black (B)** | **Non-Black (non-B)** | **All Races** | **B/non-B Fold Difference** | **†P-value B vs. non-B** | **Subject Ns** | |
|---|---|---|---|---|---|---|---|
| | | | | | | **B** | **Non-B** |
| *ADRB1* Arg389Gly (rs1801253) | 0.43 Gly | 0.28 Gly | 0.31 | 1.54 | <0.0001 | 207 | 833 |
| *ADRB1* Ser49Gly (rs1801252) | 0.24 Gly | 0.14 Gly | 0.16 | 1.71 | <0.0001 | 205 | 824 |
| *ADRB2* Gln27Glu (rs1042714) | 0.19 Glu | 0.39 Glu | 0.35 | 0.49 | <0.0001 | 207 | 833 |
| *ADRB2* Gly16Arg (rs1042713) | 0.47 Arg | 0.39 Arg | 0.41 | 1.21 | 0.004 | 207 | 833 |
| *ADRB2* Thr164Ile (rs1800888) | 0.002 Ile | 0.014 Ile | 0.012 | 0.14 | 0.067 | 207 | 833 |

†Chi-square 2-sided test (Fisher exact test substituted for low cell counts) performed on the number of alleles.

**Table S4.** Diplotypes, Genotypes BEST Adrenergic Receptor Polymorphisms Substudy.

| ***ADRB1*** **Diplotype Category** | ***ADRB1*** **Diplotype N (%) (Total = 1029)** | ***ADRB1*** **Diplotypes (Type)** | ***ADRB1*** **Genotypes** | ***ADRB2*** **Diplotype N (%) (Total = 1040)** | ***ADRB2*** **Diplotypes (Type)** | ***ADRB2*** **Genotypes** |
|---|---|---|---|---|---|---|
| 1 | 302 (29) | Ser49Arg389 Ser49Arg389 (HMZG) | Arg389Arg, Ser49Ser | 70 (7) | Gly16Gln27 Gly16Gln27 (HMZG) | Gln27Gln, Gly16Gly |
| 2 | 38 (4) | Gly49Arg389 Gly49Arg389 (HMZG) | Arg389Arg, Gly49Gly | 178 (17) | Arg16Gln27 Arg16Gln27 (HMZG) | Gln27Gln, Arg16/Arg |
| 3 | 93 (9) | Ser49Gly389 Ser49Gly389 (HMZG) | Gly389Gly, Ser49Ser | 143 (14) | Gly16Glu27 Gly16Glu27 (HMZG) | Glu27Glu, Gly16Gly |
| 4 | 0 | Gly49Gly389 Gly49Gly389 (HMZG) | Gly389Gly, Gly49Gly | 0 | Arg16Glu27 Arg16Glu27 (HMZG) | Glu27Glu, Arg16Arg |
| 5 | 149 (14) | Ser49Arg389 Gly49Arg389 (1HTZG) | Arg389Arg, Ser49Gly | 210 (20) | Arg16Gln27 Gly16Gln27 (1HTZG) | Gln27Gln, Gly16Arg, |
| 6 | 0 | Ser49Gly389 Gly49Gly389 (1HTZG) | Gly389Gly, Ser49Gly | 0 | Gly16Glu27 Arg16 Glu27 (1HTZG) | Glu27Glu' Gly16Arg |
| 7 | 101 (10) | Gly49Arg389 Ser49Gly389 (2HTZG) | Arg389Gly, Ser49Gly | 279 (27) | Arg16Gln27 Gly16Glu27 (2HTZG) | Gln27Glu, Gly16/Arg, |
| 8 | 0 | Ser49Arg389 Gly49Gly389 (2HTZG) | Arg389Gly, Ser49Gly | 0 | Gly16Gln27 Arg16Glu27 (2HTZG) | Gln27Glu, Gly16Arg |
| 9 | 346 (34) | Ser49Arg389 Ser49Gly389 (1HTZG) | Arg389Gly, Ser49Ser | 160 (15) | Gly16Gln27 Gly16Glu27 (1HTZG) | Gln27Glu, Gly16Gly |
| 10 | 0 | Gly49Arg349 Gly49Gly389 (1HTZG) | Arg389Gly, Gly49Gly | 0 | Arg16Gln27 Arg16Glu27 (1HTZG) | Gln27Glu, Arg16Arg |
| | | | | 667 (64) | 1 or 2 copies Arg16Gln27 | |
| | | | | 373 (36) | No Arg16Gln27 | |

HMZG = homozygote; 1HTZG = single heterozygote; 2HTZG = double heterozygote.

**Table S5.** Linkage Disequilibrium (LD) Coefficients for *ADRB1*, *ADRB2* polymorphisms in the BEST Substudy.

| Gene, locus 1 | Gene, locus 2 | LD Coefficient | C.I. | $X^2$ P value |
|---|---|---|---|---|
| *ADRB1* 389 | *ADRB1* 49 | 0.98 | (0.93, 1.00) | <0.0001 |
| *ADRB1* 389 | *ADRB2* 16 | 0.02 | (0.001, 0.13) | 0.64 |
| *ADRB1* 389 | *ADRB2* 27 | 0.01 | (0.001, 0.13) | 0.86 |
| *ADRB1* 49 | *ADRB2* 16 | 0.02 | (0.002, 0.18) | 0.77 |
| *ADRB1* 49 | *ADRB2* 27 | 0.06 | (0.02, 0.24) | 0.40 |
| *ADRB2* 27 | *ADRB2* 16 | 1.00 | (1.00, 1.00) | <0.0001 |
| *ADRB2* 164 | *ADRB2* 27 | 0.99 | (0.48, 0.99) | 0.0004 |
| *ADRB2* 164 | *ADRB2* 16 | 0.85 | (0.51, 0.99) | 0.0005 |
| *ADRB2* 164 | *ADRB1* 389 | 0.10 | (0.01, 0.99) | 0.45 |
| *ADRB2* 164 | *ADRB1* 49 | 0.07 | (0.01, 0.70) | 0.44 |

f = haplotype frequency (haplotype N/Total N Haplotypes).

**Table S6.** Haplotype frequencies by Race and Sex, BEST Substudy

| By Race | | | | | | |
|---|---|---|---|---|---|---|
| **Gene** | **Haplotype** | **Black (B)** **N = 410** ***ADRB1*, 414** ***ADRB2*** **(f)** | **non-Black** **N = 1648** ***ADRB1*,** **1666** ***ADRB2*** **(f)** | **All Races** **N = 2058** ***ADRB1*,** **2080** ***ADRB2*** **(f)** | **Black/non-Black Fold Difference** | **P value, Black vs. non-Black** |
| ***ADRB1*** | Ser49Arg389 | 137 (0.33) | 962 (0.58) | 1099 (0.53) | 0.57 | <0.0001 |
| *Internalizing* | Gly49Arg389 | 98 (0.24) | 228 (0.14) | 326 (0.16) | 1.71 | <0.0001 |
| | Ser49Gly389 | 175 (0.43) | 458 (0.28) | 633 (0.31) | 1.54 | <0.0001 |
| | Gly49Gly389 | 0 | 0 | 0 | – | – |
| *Internalization-resistant* | All Ser49 containing | 312 (0.76) | 1420 (0.86) | 1732 (0.84) | 0.88 | <0.0001 |
| ***ADRB2*** | Gly16Gln27 | 143 (0.35) | 367 (0.22) | 510 (0.25) | 1.59 | <0.0001 |
| *Internalizing* | Arg16Gln27 | 194 (0.47) | 651 (0.39) | 845 (0.41) | 1.21 | 0.004 |
| | Gly16Glu27 | 77 (0.19) | 648 (0.39) | 725 (0.35) | 0.54 | <0.0001 |
| | Arg16Glu27 | 0 | 0 | 0 | – | – |
| *Internalization-resistant* | Arg16Not Gln27 | 220 (0.53) | 1015 (0.61) | 1235 (0.59 | 0.87 | 0.004 |
| **By Sex and Race** | | | | | | |
| **Gene** | **Haplotype** | **Female/Black N=53 *ADRB1*, 53 *ADRB2* (f)** | **Male/Black N=152 *ADRB1*, 154 *ADRB2* (f)** | **Female/non-Black N = 159 *ADRB1*, 162 *ADRB2* (f)** | **Male/non-Black N = 665 *ADRB1*, 671 *ADRB2* (f)** | **P value by sex, Black/non-Black** |
| ***ADRB1*** | Ser49Arg389 | 35 0.33 | 102 0.34 | 179 0.56 | 783 0.59 | 1.00/0.41 |
| *Internalizing* | Gly49Arg389 | 26 0.25 | 72 0.24 | 55 0.17 | 173 0.13 | 0.89/0.057 |
| | Ser49Gly389 | 45 0.42 | 130 0.43 | 84 0.26 | 374 0.28 | 1.00/0.58 |
| *Internalization-resistant* | All Ser49 containing | 80 0.75 | 232 0.76 | 263 0.83 | 1157 0.87 | 0.89/0.057 |
| ***ADRB2*** | Gly16Gln27 | 42 0.40 | 101 0.33 | 79 0.24 | 288 0.21 | 0.24/0.26 |
| *Internalizing* | Arg16Gln27 | 50 0.47 | 144 0.47 | 117 0.36 | 534 0.40 | 1.00/0.23 |
| | Gly16Glu27 | 14 0.13 | 63 0.20 | 128 0.40 | 520 0.39 | 0.11/0.80 |
| *Internalization-resistant* | Not Arg16Gln27 | 56 0.53 | 164 0.53 | 207 0.64 | 808 0.60 | 1.00/0.23 |

f = haplotype frequency (haplotype N/Total N Haplotypes).

**Table S7.** By-subject, SEM analysis of {*ADRB1*/*ADRB2*} haplotype internalization tier data in Figures 1-3, S1-S3, with event probability in tiers ordered from most (Tier 1, ≥3DI, 3-4 internalizing haplotypes) to least internalizing (Tier 3, ≥3DIR, 0-1 internalizing haplotypes).

| **Endpoint** | **Placebo** | | **Bucindolol** | | **Bucindolol/Placebo Treatment Effect** | |
|---|---|---|---|---|---|---|
| | **Slope*** | **Slope P Value** | **Slope*** | **Slope P Value** | **Slope†** | **Slope P Value** |
| | **All LVEF** | | | | | |
| ACM | 0.17±0.09 | 0.049 | -0.09 ± 0.10 | 0.37 | -0.26 ± 0.13 | 0.047 |
| ACM/Tx | 0.19 ± 0.08 | 0.026 | -0.09 ± 0.09 | 0.36 | -0.27 ± 0.13 | 0.03 |
| CVM | 0.13 ± 0.09 | 0.140 | -0.04 ± 0.10 | 0.68 | -0.17 ± 0.14 | 0.21 |
| CVH | 0.23 ± 0.07 | 0.002 | 0.13 ± 0.07 | 0.07 | -0.10 ± 0.10 | 0.33 |
| HFH | 0.19 ± 0.08 | 0.013 | 0.08 ± 0.07 | 0.25 | -0.11 ± 0.11 | 0.32 |
| ACM/AF/AFL | 0.10 ± 0.07 | 0.16 | -0.03 ± 0.08 | 0.74 | -0.13 ± 0.11 | 0.22 |
| CVM/HFH | 0.21 ± 0.07 | 0.004 | 0.06 ± 0.07 | 0.43 | -0.16 ± 0.10 | 0.12 |
| All 7 Endpoints | 0.18 ± 0.06 | 0.002 | 0.02 ± 0.06 | 0.78 | -0.16 ± 0.08 | 0.048 |
| | **LVEF ≥0.20** | | | | | |
| ACM | 0.15 ± 0.11 | 0.20 | -0.19 ± 0.13 | 0.150 | -0.34 ± 0.17 | 0.050 |
| ACM/Tx | 0.19 ± 0.11 | 0.098 | -0.19 ± 0.13 | 0.154 | -0.38 ± 0.17 | 0.028 |
| CVM | 0.10 ± 0.12 | 0.38 | -0.17 ± 0.16 | 0.74 | -0.27 ± 0.19 | 0.158 |
| CVH | 0.22 ± 0.09 | 0.015 | -0.06 ± 0.09 | 0.51 | -0.28 ± 0.13 | 0.027 |
| HFH | 0.21 ± 0.10 | 0.034 | -0.07 ± 0.09 | 0.46 | -0.28 ± 0.13 | 0.037 |
| ACM/AF/AFL | 0.07 ± 0.09 | 0.45 | -0.15 ± 0.11 | 0.152 | -0.22 ± 0.14 | 0.11 |
| CVM/HFH | 0.24 ± 0.09 | 0.010 | -0.11 ± 0.09 | 0.19 | -0.36 ± 0.13 | 0.005 |
| All 7 Endpoints | 0.17 ± 0.08 | 0.022 | -0.13 ± 0.07 | 0.074 | -0.30 ± 0.10 | 0.003 |

*From Tier 1 to Tier 3; †Bucindolol slope minus placebo slope; Interaction between bucindolol and placebo group data.

**Table S8.** GENETIC-AF DNA Substudy subject characteristics by diplotype, (Mean±SD), GENETIC-AF Substudy (N = 138.

| Diplotype, Treatment | N | Age (Yrs) | M/F (%) | †Race/ Ethnicity | HF Duration (Mos) | AF Duration (Mos | BMI ($Kg/Ht^2$) | Hx DM (%) | Hx Htn (%) | NYHA III/IV (%) | LVEF (%) | HR (bpm) | SBP (mmHg) | AF (%) | ‡AF Type |
|---|---|---|---|---|---|---|---|---|---|---|---|---|---|---|---|
| ***ADRB1*** | | | | | | | | | | | | | | | |
| Ser49Arg389<br>Ser49Arg389 | 96 | 66<br>±11 | 84/<br>16 | 1/0/1<br>/0/0/98* | 36<br>±60 | 36<br>±48 | 33<br>±7 | 30 | 77 | 28/53/19 | 36<br>±9 | 75<br>±16 | 125<br>±15 | 51/<br>49 | 46/<br>54 |
| Gly49Arg389<br>Gly49Arg389 | 1 | 64<br>±NA | 100/<br>0 | 0/0/0<br>/0/0/100 | 60 | 0 | 24<br>±NA | 0 | 100 | 0/0/100 | 18<br>±NA | 76<br>±NA | 105<br>±NA | 0/<br>100 | 100/<br>0 |
| Ser49Arg389<br>Gly49Arg389 | 41 | 65<br>±11 | 85/<br>15 | 0/7/0<br>/0/0/93* | 48<br>±84 | 36<br>±48 | 32<br>±7 | 24 | 78 | 24/59/17 | 36<br>±9 | 77<br>±19 | 121<br>±16 | 56/<br>44 | 49/<br>51 |
| ***ADRB2*** | | | | | | | | | | | | | | | |
| Gly16Gln27<br>Gly16Gln27 | 5 | 69<br>±7 | 60/<br>40 | 0/0/0<br>/0/0/100 | 60<br>±84 | 12<br>±12 | 30<br>±3 | 40 | 100 | 20/60/20 | 34<br>±14 | 65<br>±4 | 125<br>±17 | 20/<br>80 | 60/<br>40 |
| Arg16Gln27<br>Arg16Gln27 | 23 | 65<br>±16 | 87/<br>13 | 4/4/0<br>/0/0/91 | 36<br>±72 | 24<br>±36 | 31<br>±9 | 26 | 61 | 30/57/13 | 35<br>±9 | 75<br>±16 | 119<br>±16 | 61/<br>39 | 52/<br>48 |
| Gly16Glu27<br>Gly16Glu27 | 23 | 65<br>±16 | 87/<br>13 | 4/4/0<br>/0/0/91 | 36<br>±72 | 24<br>±36 | 31<br>±9 | 26 | 61 | 30/57/13 | 35<br>±9 | 75<br>±16 | 119<br>±16 | 61/<br>39 | 52/<br>48 |
| Arg16Gln27<br>Gly16Gln27 | 17 | 65<br>±8 | 82/<br>18 | 0/6/0<br>/0/0/94 | 24<br>±36 | 36<br>±48 | 32<br>±6 | 6 | 82 | 47/29/24 | 39<br>±10 | 72<br>±17 | 127<br>±15 | 41/<br>59 | 41/<br>59 |
| Arg16Gln27<br>Gly16Glu27 | 47 | 64<br>±9 | 89/<br>11 | 0/0/0<br>/0/0/100 | 24<br>±36 | 36<br>±60 | 33<br>±8 | 28 | 85 | 21/62/17 | 37<br>±9 | 79<br>±15* | 125<br>±15 | 55/<br>45 | 36/<br>64 |
| Gly16Gln27<br>Gly16Glu27 | 23 | 68<br>±11 | 78/<br>22 | 0/4/4<br>/0/0/91 | 48<br>±96 | 36<br>±60 | 32<br>±6 | 26 | 74 | 17/61/22 | 36<br>±8 | 79<br>±21 | 121<br>±16 | 57/<br>43 | 61/<br>39 |
| Metoprolol | 68 | 65<br>±11 | 82/<br>18 | 0/3/1<br>/0/0/96 | 36<br>±60 | 24<br>±48 | 33<br>±8 | 25 | 87 | 22/51/26 | 36<br>±8 | 75<br>±19 | 124<br>±16 | 49/<br>51 | 44/<br>56 |
| Bucindolol | 70 | 66<br>±11 | 87/<br>13 | 1/1/0<br>/0/0/97 | 36<br>±72 | 36<br>±48 | 32<br>±6 | 31 | 69 | 31/57/11 | 36<br>±9 | 77<br>±14 | 123<br>±16 | 56/<br>44 | 50/<br>50 |

*P<0.05, **P<0.01, ***P<0.001 vs. all other members of the same genotype group. †In sequence: White/Black/Hispanic/Asian/Pacific Islander, American Indian, Alaskan/Other; ‡Paroxysmal/Persistent. HF=heart failure, HxDM = history of diabetes, Hx Htn=history of hypertension, HR=heart rate, SBP=systolic blood pressure, AF= atrial fibrillation.

**Table S9.** Genotypes, haplotypes, and diplotypes in the GENETIC-AF Trial

| **Genotypes** | | | | | | | | |
|---|---|---|---|---|---|---|---|---|
| **β-AR gene** | ***ADRB1* (N= 70 Bucindolol, 68 Metoprolol)** | | | | ***ADRB2* (N= 70 Bucindolol, 68 Metoprolol)** | | | |
| **Position, (N) {%}** | **389 (N) {%}** | | **49 (N) {%}** | | **27 (N) {%}** | | **16 (N) {%}** | |
| **Treatment Arm** | **Bucindolol** | **Metoprolol** | **Bucindolol** | **Metoprolol** | **Bucindolol** | **Metoprolol** | **Bucindolol** | **Metoprolol** |
| **Genotypes** | Arg389Arg (70) {100} | Arg389Arg (68) {100} | Ser49Ser (48) {69} | Ser49Ser (48) {71} | Gln27/Gln (22) {31} | Gln27/Gln (23) {34} | Gly16Gly (26) {37} | Gly16Gly (25) {37} |
| | Arg389Gly (0) {0} | Arg389Gly (0) {0} | Ser49Gly (22) {31} | Ser49Gly (19) {28} | Gln27Glu (35) {50} | Gln27Glu (35) {52} | Gly16Arg (34) {49} | Gly16Arg (30) {44} |
| | Gly389Gly (0) {0} | Gly389Gly (0) {0} | Gly49Gly (0) {0} | Gly49Gly (1) {1} | Glu/27Glu (13) {19} | Glu/27Glu (10) {15} | Arg16Arg (10) {14} | Arg16Arg (13) {19} |

| **Haplotypes** | | | | |
|---|---|---|---|---|
| **β-AR genes** | ***ADRB1*** | | ***ADRB2*** | |
| **Treatment Arm** | **Bucindolol** | **Metoprolol** | **Bucindolol** | **Metoprolol** |
| **Haplotypes** | Ser49Arg389 (118) {84} | Ser49Arg389 (115) {85} | Gly16Gln27 (25) {18} | Gly16Gln27 (25) {18} |
| | Gly49Arg389 (22) {16} | Gly49Arg389 (21) {15} | Arg16Gln27 (54) {39} | Arg16 Gln27 (56) {41} |
| | | | Gly16Glu27 (61) {44} | Gly16 Glu27 (55) {40} |
| | | | Arg16Glu27 (0) {0} | Arg16Glu27 (0) {0} |

| **Diplotypes** | | | | |
|---|---|---|---|---|
| **β-AR gene** | ***ADRB1*** | | ***ADRB2*** | |
| **Treatment Arm** | **Bucindolol** | **Metoprolol** | **Bucindolol** | **Metoprolol** |
| **Diplotypes** | Ser49Arg389 Ser49Arg389 (48) {34} | Ser49Arg389 Ser49Arg389 (48) {35} | Gly16Gln27 Gly16Gln27 (4) {3} | Gly16Gln27 Gly16Gln27 (1) {1} |
| | Gly49Arg389 Gly49Arg389 (0) {0} | Gly49Arg389 Gly49Arg389 (1) {1} | Gly16Glu27 Gly16Glu27 (13) {10} | Gly16Glu27 Gly16Glu27 (10) {8} |
| | Ser49Arg389 Gly49Arg389 (22) {16} | Ser49Arg389 Gly49Arg389 (19) {14} | Arg16Gln27 Arg16Gln27 (10) {8} | Arg16Gln27 Arg16Gln27 (13) {10} |
| | | | Arg16Glu27 Arg16Glu27 (0) {0} | Arg16Glu27 Arg16Glu27 (0) {0} |
| | | | Gly16Gln27 Gly16Glu27 (9) {6} | Gly16Gln27 Gly16Glu27 (14) {10} |
| | | | Arg16Gln27 Arg16Glu27 (0) {0} | Arg16Gln27 Arg16Glu27 (0) {0} |

| | | | | |
|---|---|---|---|---|
| | | | Arg16Gln27<br>gly16Gln27<br>(8) {6} | Arg16Gln27<br>Gy16Gln27<br>(9) {7} |
| | | | Gly16Glu27<br>Arg16Glu27<br>(0) {0} | Gly16Glu27<br>Arg16Glu27<br>(0) {0} |
| | | | Gly16Gln27<br>Arg16Glu27<br>(0) {0} | Gly16Gln27<br>Arg16Glu27<br>(0) {0} |
| | | | Arg16Gln27<br>Gly16Glu27<br>(26) {19} | Arg16Gln27<br>Gly16Glu27<br>(21) {15} |

**Table S10.** NT-proBNP (pg/ml), Bucindolol Arm.

| **Maximum (4) Double Internalization-resistant diplotypes (4DIR, no copies of *ADRB1* Gly49Arg389 AND no copies of *ADRB2* Arg16Gln27).** | | | | | | |
|---|---|---|---|---|---|---|
| **Receptor Gene** | **Diplotype Combinations** | **N** | **Baseline** ±SD | **Δ, 4 Weeks** ±SEM | **Δ, 12 Weeks** ±SEM | **Δ, 24 Weeks** ±SEM |
| *ADRB1* | Ser49Arg389 | 7 | 992 ±977 | -905 ±546 | -902 ±562 | -948 ±470 |
| | Ser49Arg389 | | | | | |
| *ADRB2* | Gly16Glu27 | | | | | |
| | Gly16Glu27 | | | | | |
| *ADRB1* | Ser49Arg389 | 7 | 1953 ±1606 | -658 ±385 | -37.3 ±327 | -728 ±310 |
| | Ser49Arg389 | | | | | |
| *ADRB2* | Gly16Glu27 | | | | | |
| | Gly16Gln27 | | | | | |
| *ADRB1* | Ser49Arg389 | 2 | 123 ±74.6 | -34.7 ± NA | 2.0 ±23.4 | -0.4 ± NA |
| | Ser49Arg389 | | | | | |
| *ADRB2* | Gly16Gln27 | | | | | |
| | Gly16Gln27 | | | | | |
| *ADRB1* *ADRB2* | **4DIR Total** | 16 | 1304 ±1313 | -737 ±312 | -493 ±329 | -778 ±266 |
| Statistical Analysis, 4DIR: | | | | | | |
| - | P value (WSR*) | - | - | 0.009 | 0.15 | 0.005 |

*Wilcoxon signed rank test, values compared to baseline (pre-randomization values).

**Table S11.** NT-proBNP, Metoprolol Arm.

| **Maximum (4) Double Internalization-resistant Diplotypes (4DIR), no copies of *ADRB1* Gly49Arg389 AND no copies of *ADRB2* Arg16Gln27.** | | | | | | |
|---|---|---|---|---|---|---|
| **Receptor** | **Diplotype Combinations** | **N** | **Baseline ±SD** | **Δ, 4 Weeks ±SEM** | **Δ, 12 Weeks ±SEM** | **Δ, 24 Weeks ±SEM** |
| *ADRB1* | Ser49Arg389 | 5 | 897 ±1096 | -246 ±228 | 509 ±502 | -96.0 ±183 |
| | Ser49Arg389 | | | | | |
| *ADRB2* | Gly16Glu27 | | | | | |
| | Gly16Glu27 | | | | | |
| *ADRB1* | Ser49Arg389 | 8 | 1241 ±1172 | 72.7 ±377 | -79.9 ±466 | 559 ±756 |
| | Ser49Arg389 | | | | | |
| *ADRB2* | Gly16Glu27 | | | | | |
| | Gly16Gln27 | | | | | |
| *ADRB1* | Ser49Arg389 | 1 | 1983 ± NA | -784 ± NA | NaN ± NA | -681 ± NA |
| | Ser49Arg389 | | | | | |
| *ADRB2* | Gly16Gln27 | | | | | |
| | Gly16Gln27 | | | | | |
| *ADRB1* *ADRB2* | **4DIR Total** | 14 | 1171 ±1053 | -91.1 ±245 | 156 ±339 | 238 ±461 |
| Statistical Analysis, 4DIR: | | | | | | |
| | P value (WSR*) | - | - | 0.588 | 0.432 | 0.770 |

*Wilcoxon signed rank test, values compared to baseline (pre-randomization values).

**Table S12** Norepinephrine (NE, pg/ml), Bucindolol Arm.

| **Maximum (4) Double Internalization-resistant diplotypes (4DIR, no copies of *ADRB1* Gly49Arg389 AND no copies of *ADRB2* Arg16Gln27).** | | | | | | |
|---|---|---|---|---|---|---|
| **Receptor Gene** | **Diplotype Combinations** | **N** | **Baseline** ±Sem | **Δ, 4 Weeks** ±SEM | **Δ, 12 Weeks** ±SEM | **Δ, 24 Weeks** ±SEM |
| *ADRB1* | Ser49Arg389 | 9 | 581 ±232 | -121 ±62.2 | -139 ±41.5 | -47.7 ±57.7 |
| | Ser49Arg389 | | | | | |
| *ADRB2* | Gly16Glu27 | | | | | |
| | Gly16Glu27 | | | | | |
| *ADRB1* | Ser49Arg389 | 7 | 690 ±140 | -169 ±88.5 | -166 ±123 | -232 ±53.8 |
| | Ser49Arg389 | | | | | |
| *ADRB2* | Gly16Glu27 | | | | | |
| | Gly16Gln27 | | | | | |
| *ADRB1* | Ser49Arg389 | 3 | 644 ±325 | -184 ±285 | -198 ±247 | -232 ±110 |
| | Ser49Arg389 | | | | | |
| *ADRB2* | Gly16Gln27 | | | | | |
| | Gly16Gln27 | | | | | |
| ADRB1 ADRB2 | **4DIR Total** | 19 | 631 ±48.6 | -147 ±50.6 | -158 ±55.8 | -140 ±43.3 |
| Statistical Analysis, 4DIR: | | | | | | |
| - | P value (WSR*) | - | - | 0.014 | 0.01 | 0.016 |

*Wilcoxon signed rank test, values compared to baseline (pre-randomization values). Δ = change from Baseline.

**Table S13** Norepinephrine (NE, pg/ml), Metoprolol Arm.

| **Maximum (4) Double Internalization-resistant diplotypes (4DIR, no copies of *ADRB1* Gly49Arg389 AND no copies of *ADRB2* Arg16Gln27).** | | | | | | |
|---|---|---|---|---|---|---|
| **Receptor** | **Diplotype Combinations** | **N** | **Baseline ±SD** | **Δ, 4 Weeks ±SEM** | **Δ, 12 Weeks ±SEM** | **Δ, 24 Weeks ±SEM** |
| *ADRB1* | Ser49Arg389 | 5 | 635 ±208 | 41.2 ±91.5 | -9.0 51.8 | 58.0 ±150 |
| | Ser49Arg389 | | | | | |
| *ADRB2* | Gly16Glu27 | | | | | |
| | Gly16Glu27 | | | | | |
| *ADRB1* | Ser49Arg389 | 8 | 501 ±58.4 | -37.5 ±47.5 | 6.4 ±95.6 | 49.0 ±94.6 |
| | Ser49Arg389 | | | | | |
| *ADRB2* | Gly16Glu27 | | | | | |
| | Gly16Gln27 | | | | | |
| *ADRB1* | Ser49Arg389 | 1 | 879 | -265 | – | -474 |
| | Ser49Arg389 | | | | | |
| *ADRB2* | Gly16Gln27 | | | | | |
| | Gly16Gln27 | | | | | |
| *ADRB1* *ADRB2* | **4DIR Total** | 14 | 576 ±52.2 | -30.8 ±44.0 | 0.8 ±61.5 | 8.4 ±82.7 |
| Statistical Analysis, 4DIR: | | | | | | |
| | P value (WSR*) | - | - | 0.455 | 0.700 | 0.622 |

*Wilcoxon signed rank test, values compared to baseline (pre-randomization values). Δ = change from Baseline.

**Table S14.** GENETIC-AF subject characteristics by haplotype group and β-blocker treatment at randomization, (mean±sd), N = 138).

| Internalization Category; BB Previous | N | Age (Yrs) | M/F (%) | [†]Race/ Ethnicity | HF Duration (Yrs) | AF Duration (Yrs) | BMI (Kg/ $Ht^2$) | Hx DM (%) | Hx Htn (%) | NYHA I/II/ III (%) | LVEF (%) | HR (bpm) | SBP (mm Hg) | Rhythm @ Rand AF/SR | AF Type PAR/PER @ Rand | Pre-randomization BB Dose (Carvedilol Equivalents mg/d, (mg/kg/d)) |
|---|---|---|---|---|---|---|---|---|---|---|---|---|---|---|---|---|
| ≥ 2IR BB most recent: Carvedilol | 24 | 64 ±12 | 96/ 4 | 4/0/0 /0/0/96 | 6 ±8** | 3 ±4 | 34 ±8 | 33 | 71 | 17/58/25 | 31 ±7*** | 77 ±15 | 117 ±17* | 71/ 29 | 46/ 54 | 38±21 (0.35±0.16) |
| ≥ 2IR BB most recent: Not Carvedilol | 89 | 67 ±11 | 82/ 18 | 0/1/1 /0/0/98 | 2 ±4** | 3 ±4 | 32 ±7 | 29 | 79 | 28/55/17 | 38 ±8** | 75 ±17 | 127 ±15** | 46/ 54 | 48/ 52 | 38±27 (0.39±0.25) |
| ≥ 2DI BB most recent: Carvedilol | 9 | 65 ±5 | 78/ 22 | 0/11/0 /0/0/89 | 3 ±3 | 3 ±3 | 29 ±4 | 22 | 100 | 11/56/33 | 32 ±9 | 79 ±13 | 114 ±13 | 56/ 44 | 33/ 67 | 28±17 (0.33±0.22) |
| ≥ 2DI BB most recent: Not Carvedilol | 16 | 63 ±13 | 88/ 12 | 0/6/0 /0/0/94 | 2 ±3 | 3 ±5 | 31 ±8 | 19 | 69 | 44/44/12 | 37 ±11 | 78 ±18 | 122 ±14 | 56/ 44 | 50/ 50 | 30±16 (0.31±0.16) |

[†]In sequence: A/B/H/I/O/W; BB = β-blocker, PAR = paroxysmal AF, PER = persistent AF, Rand = randomization; ** = P <0.01 vs. all other categories, *** = P < 0.001 vs. all other categories; [‡]Carvedilol 50 mg/d Equivalents: Metoprolol 150 mg/d, Bisoprolol 10mg/d, Nebivolol 10mg/d; See Table S6 for definition of haplotype groups. Number of Not Carvedilol subjects per β-blocker and actual dose (mg/d): metoprolol, 117±60 (N=59); bisoprolol, 6±4 (N=37); nebivolol, 40 (1).

**Table S15.** Target Expansion and Treatment Effect enhancement by haplotype (diplotypes) selection, Treatment Effects in % ((1-Hazard Ratio) x 100). The Cohort is the BEST Adrenergic Receptor Polymorphism Substudy with LVEFs ≥0.20 (mean±SD 0.273±0.046).

| **Treatment Effect (%), (P value), # events Bucindolol (B) Placebo (P) →**<br>**Endpoint** | **1. DNA SS All Genotypes, haplotypes; N=731** | **2. *ADRB1* Arg389Arg Genotype (Pharmaco-genetic Index Subgroup); N=331** | **3. *ADRB1* Ser49Arg389 homozygous Diplotype (Internalization-resistant *ADRB1*); N=200** | **4. *ADRB1* Ser49Gly389 homozygous Diplotype (Internalization-resistant *ADRB1*); N=67** | **5. *ADRB2* Not-Arg16Gln27; Containing Diplotypes (Internalization-resistant *ADRB2*); N=272** | **6. *ADRB1*[‖] AND *ADRB2* Diplotypes (4DIR, Internalization-resistant); N=184** | **7. *ADRB1* Arg389Arg Genotype OR 4DIR *ADRB1* AND *ADRB2* Diplotypes; N=441** | **8. *ADRB1* Ser49Arg389 Homozygous Diplotype AND Internalization-resistant *ADRB2* Diplo-types (subset of 4DIR); N=74** |
|---|---|---|---|---|---|---|---|---|
| ACM | 39<br>(P=0.011)<br>43B, 69P | 52<br>(P=0.016)<br>17B,30P | 54<br>(P=0.052)<br>10B, 18P | 70<br>(P=0.085)<br>3B, 8P | 62<br>(P=0.009)<br>10B, 26P | 81<br>(P=0.009)<br>3B, 17P | 54<br>(P=0.005)<br>20B, 39P | 100<br>(P=NA)<br>0B, 8P |
| ACM/Tx | 38<br>(P=0.012)<br>46B, 72P | 52<br>(P=0.016)<br>17B, 30P | 54<br>(P=0.052)<br>10B, 18P | 75<br>(P=0.039)<br>3B, 10P | 66<br>(P=0.003)<br>10B, 29P | 83<br>P=0.004)<br>3B, 20P | 57<br>(P=0.002)<br>20B, 42P | 100<br>(P=NA)<br>0B, 8P |
| CVM | 49<br>(P=0.003)<br>31B, 59P | 58<br>(P=0.015)<br>12B, 24P | 49<br>(P=0.13)<br>8B, 13P | 78<br>(P=0.062<br>2B, 7P | 69<br>(P=0.007)<br>7B, 22P | 84<br>(P=0.014)<br>2B, 14P | 60<br>(P=0.004)<br>14B, 32P | 100<br>(P=NA)<br>0B, 6P |
| HF Hosp | 34<br>(P=0.002)<br>90B, 129P | 37<br>(P=0.021)<br>43B, 56P | 56<br>(P=0.002)<br>22B, 38P | 58<br>(P=0.103)<br>5B, 12P | 42<br>(P=0.016)<br>32B, 52P | 43<br>(P=0.058)<br>18B, 34P | 38<br>(P=0.009)<br>53B, 73P | 41<br>(P=0.22<br>8B, 17P |
| CVH Hosp | 32<br>(P=0.001)<br>125B, 175P | 43<br>(P=0.001)<br>54B, 76P | 51<br>(P=0.002)<br>32B, 48P | 36<br>(P=0.24)<br>11B, 19P | 45<br>(P=0.002)<br>45B, 74P | 47<br>(P=0.007)<br>28B, 53P | 44<br>(P=0.0001)<br>72B, 106P | 52<br>(P=0.053)<br>10B, 23P |
| ACM/HFH | 34<br>(P=0.001)<br>114B, 161P | 38<br>(P=0.011)<br>52B, 67P | 55<br>(P=0.001)<br>27B, 45P | 67<br>(P=0.015)<br>7B, 19P | 44<br>(P=0.005)<br>39B, 64P | 50<br>(P=0.012)<br>20B, 42P | 40<br>(P=0.002)<br>64B, 90P | 48<br>(P=0.12)<br>8B, 19P |
| CVM/HFH | 37<br>(P=0.0001)<br>106B, 156P | 42<br>(P=0.005)<br>47B, 65P | 56<br>(P=0.001)<br>25B, 43P | 65<br>(P=0.021)<br>7B, 18P | 46<br>(P=0.004)<br>36B, 62P | 51<br>(P=0.011)<br>19B, 41P | 43<br>(P=0.001)<br>58B, 87P | 48<br>(P=0.12)<br>B8, P19 |
| AF/AFL/ACM | 32<br>(P=0.046)<br>63B, 98P | 50<br>(P=0.020)<br>(25B, 43P | 46<br>(P=0.103)<br>15B, 25P | 48<br>(P=0.29)<br>5B, 9P | 60<br>(P=0.008)<br>19B, 38P | 59<br>(P=0.042)<br>11B, 25P | 48<br>(P=0.013)<br>33B, 57P | 76<br>(P=0.07)<br>3B, 11P |

| | | | | | | | | |
|---|---|---|---|---|---|---|---|---|
| All Endpoints, Mean ± SD | 36.9 ±5.6 | 46.5‡ ±7.6 | 52.6* ±3.6 | 62.1*,§ ±14.2 | 54.3*,† ±11.1 | 62.3*,† ±17.5 | 48.0* ±8.2 | 70.6*,† ±26.4 |
| Fold difference (FD) vs. Columns 1/2 | 1.00/ 0.80±0.10 | 1.27±0.16/ 1.00 | 1.45±0.21/ 1.13 | 1.68±0.28/ 1.33 | 1.47±0.24/ 1.17 | 1.68±0.34/ 1.34 | 1.30±0.15 1.03 | 1.92±0.58/ 1.52 |

4DIR=Maximum Double Internalization-resistant diplotypes (2 copies of *ADRB1* AND 2 copies of *ADRB2* internalization-resistant haplotypes). *P value (paired t) <0.01 vs. Column1; †P <0.01 vs. Column 2; ‡P<0.05 vs. Column 1; §P value <0.05 vs. Col 2 ‖Either Gly or Arg389 AND Ser49 diplotypes.

**Table S16.** Target Expansion and Treatment Effect enhancement by haplotype (diplotypes) selection, Treatment Effects in % ((1-Hazard Ratio) x 100). The Cohort is the BEST Adrenergic Receptor Polymorphism Substudy with All LVEFs mean±SD 0.236±0.071).

| **Treatment Effect (%), (P value), # events Bucindolol (B) Placebo (P) →** / **Endpoint** | **1. DNA SS All Genotypes, Haplotypes N=1040** | **2. *ADRB1* Arg389Arg Genotype (Pharmaco-genetic Index Subgroup); N=493** | **3. *ADRB1* Ser49Arg389 homozygous Diplotype (Internalization-resistant *ADRB1*); N=302** | **4. *ADRB1* Ser49Gly389 homozygous Diplotype (Internalization-resistant *ADRB1*); N=93** | **5. A*DRB2* Not-Arg16Gln27; Containing Diplotypes (Internalization-resistant *ADRB2*); N=373** | **6. *ADRB1*‖ AND *ADRB2* (4DIR) Internalization-resistant Diplotypes; N=267** | **7. ADRB1 Arg389Arg Genotype OR 4DIR *ADRB1* AND *ADRB2* Diplotypes; N=649** | **8. A*DRB1* Arg389Ser49 homozygous Diplotype AND Internalization-resistant *ADRB2* Diplo-types (subset of 4DIR); N=111** |
|---|---|---|---|---|---|---|---|---|
| ACM | 23 (P=0.073) 84B, 105P | 36 (P=0.050) 35B, 45P | 30 (P=0.197) 23B, 28P | 61 (P=0.058) 6B, 14P | 45 (P=0.020) 24B, 40P | 52 (P=0.023) 14B, 29P | 37 (P=0.021) 45B, 61P | 69 (P=0.038) 4B, 13P |
| ACM/Tx | 26 (P=0.032) 90B, 117P | 40 (P=0.019) 37B, 51P | 36 (P=0.096) 24B, 32P | 59 (P=0.052) 7B, 16P | 50 (P=0.005) 25B, 46P | 58 P=0.006) 15B, 35P | 42 (P=0.004) 48B, 71P | 73 (0.020) 4B, 15P |
| CVM | 33 (P=0.012) 65B, 94P | 43 (P=0.025) 26B, 38P | 35 (P=0.19) 17B, 22P | 66 (P=0.045 5B, 13P | 45 (P=0.030) 21B, 35P | 49 (P=0.052) 13B, 25P | 43 (P=0.010) 35B, 53P | 60 (P=0.12 4B, 10P |
| HF Hosp | 26 (P=0.007) 149B, 186P | 36 (P=0.006) 69B, 85P | 46 (P=0.002) 41B, 60P | 38 (P=0.22) 10B, 18P | 24 (P=0.012) 57B, 67P | 9 (P=0.67) 42B, 47P | 26 (P=0.037) 96B, 107P | 35 (0.19) 15B, 25P |
| CVH Hosp | 21 (P=0.013) 125B, 175P | 34 (P=0.003) 93B, 111P | 42 (P=0.002) 57B, 76P | 25 (P=0.37) 16B, 25P | 22 (P=0.10) 80B, 93P | 14 (P=0.39) 59B, 68P | 26 (P=0.013) 131B, 147P | 37 (P=0.096) 21B, 33P |
| ACM/HFH | 24 | 33 | 43 | 48 | 28 | 20 | 27 | 41 |

| | | | | | | | | |
|---|---|---|---|---|---|---|---|---|
| | (P=0.004)<br>192B, 233P | (P=0.006)<br>87B, 102P | (P=0.003)<br>51B, 70P | (P=0.051)<br>14B, 28P | (P=0.004)<br>70B, 86P | (P=0.024)<br>48B, 61P | (P=0.013)<br>119B, 134P | (P=0.095)<br>16B, 29P |
| CVM/HFH | 28<br>(P=0.001)<br>179B, 228P | 37<br>(P=0.002)<br>80B, 100P | 46<br>(P=0.001)<br>47B, 68P | 46<br>(P=0.066)<br>14B, 27P | 29<br>(P=0.034)<br>67B, 84P | 21<br>(P=0.24)<br>47B, 60P | 30<br>(P=0.006)<br>111B, 131P | 41<br>(P=0.095)<br>B16, P29 |
| AF/AFL/ACM | 22<br>(P=0.088)<br>113B, 151P | 36<br>(P=0.049)<br>(48B, 67P | 31<br>(P=0.19)<br>31B, 42P | 39<br>(P=0.28)<br>9B, 13P | 45<br>(P=0.018)<br>38B. 56P | 45<br>(P=0.049)<br>26B, 41P | 36<br>(P=0.024)<br>66B, 90P | 57<br>(P=0.08)<br>8B, 18P |
| All Endpoints, Mean ± SD | 25.4<br>±3.8 | 36.9*<br>±3.2 | 38.6*<br>±6.5 | 47.8[*,§]<br>±13.8 | 36.0[‡]<br>±11.3 | 33.5<br>±19.4 | 33.4<br>±7.0 | 51.6[*,§]<br>±15.0 |
| Fold difference (FD) vs. Columns 1/2 | 1.00/0.69 | 1.47±0.14/<br>1.00 | 1.55±0.31/<br>1.05 | 1.87±0.46/<br>1.30 | 1.43±0.47/<br>0.98 | 1.32±0.26/<br>0.91 | 1.32±0.91/<br>0.91 | 2.06±0.64/<br>1.40 |

4DIR=Maximum Double Internalization-resistant diplotypes (2 copies of *ADRB1* AND 2 copies of *ADRB2* internalization-resistant haplotypes). *P value (paired t) <0.01 vs. Column1; [†]P <0.01 vs. Column 2; [‡]P<0.05 vs. Column 1; [§]P value <0.05 vs. Col 2 [ǀ]Either Gly or Arg389 AND Ser49 diplotypes.

**Table S17.** Explanted human heart RV trabeculae, ratio of pERK1,2/total ERK1,2 densitometry normalized to vehicle control after a 5 or 10 minute incubation.

| **Explanted Hearts** | **1** | **2** | **3** | **4** | **5** | **6** | **Mean or Total** |
|---|---|---|---|---|---|---|---|
| Type | NDC‡ | AR, NDC | RCM§ | ICM | NDC | NDC | 4 different Dxs |
| Age | 32 | 29 | 54 | 56 | 49 | 72 | 48.7±16.1 |
| Sex | M | M | M | M | M | M | 6 M |
| Race/ethnicity | Black | Non-Black (White/Hispanic) | Non-Black (White) | Black | Non-Black (White) | Non-Black (White) | 4 Non-black 2 Black |
| LVEF | 0.16 | 0.24 | 0.28 | 0.10 | 0.19 | 0.14 | 0.18±0.07 |
| *ADRB1* Diplotype | Ser49Arg389 Ser49Arg389 | Ser49Arg389 Ser49Gly389 | Gly49Arg389 Ser49Gly389 | Ser49Gly389 Ser49Gly389 | Ser49Arg389 Ser49Arg389 | Ser49Arg389 Ser49Gly389 | Internalizing haplotypes: 5 with 0, 1 with 1 |
| *ADRB2* Diplotype | Arg16Gln27 Gly16Gln27 | Arg16Gln27 Gly16Gln27 | Arg16Gln27 Gly16Gln27 | Arg16Gln27 Gly16Gln27 | Arg16Gln27 Gly16Glu27 | Gly16Glu27 Gly16Glu27 | Internalizing haplotypes: 5 with 1, 1 with 0 |
| # $\beta_1/\beta_2$ Internalizing haplotypes | 0/1 | 0/1 | 1/1 | 0/1 | 0/1 | 0/0 | One 1/1, four 0/1, one 0/0 |
| Classification* | 3DIR (3 Internalization-resistant) | 3DIR (3 (Internalization-resistant) | 2DI (2 Internalizing, 1 each gene) | 3DIR (3 Internalization-resistant) | 3DIR (3 Internalization-resistant) | 4DIR (Maximum Internalization-resistant) | Four 3DIR, One 2DI One 4DIR |
| **pERK1,2 Densitometry (pERK/Total ERK) x 100 (%))** | | | | | | | |
| Incubation Time (Min.) | 5 | 10 | 10 | 10 | 10 | 10 | One 5, five 10† |
| Vehicle | 0.78 | 1.06 | 1.19 | 1.34 | 1.12 | 0.88 | 1.06 ±0.14 |
| Isoproterenol 1e-6M | ND | 1.04 | 1.33 | 1.75 | 1.70 | 0.98 | 1.36 ±0.36 |
| Bucindolol 1e-6M | 1.00 | 1.27 | 1.54 | 1.81 | 1.42 | 1.36 | 1.40‡ ±0.27 |
| Metoprolol 1e-5M | 0.89 | 1.25 | 1.50 | ND | 1.18 | 0.82 | 1.13 ±0.28 |

*As described in Figures 1 and S1; NDC=nonischemic dilated cardiomyopathy; AR = aortic regurgitation; RCM=restrictive cardiomyopathy (hemochromatosis); ICM= ischemic cardiomyopathy; Dxs =diagnoses; ND = not done; †All 6 included in statistical analysis (ANOVA/Mixed Model with repeated measures, Holm-Sidak's multiple comparison test vs. Vehicle); Mean±SD; ‡P =0.0027 vs. Vehicle.

### SUPPLEMENTAL FIGURES

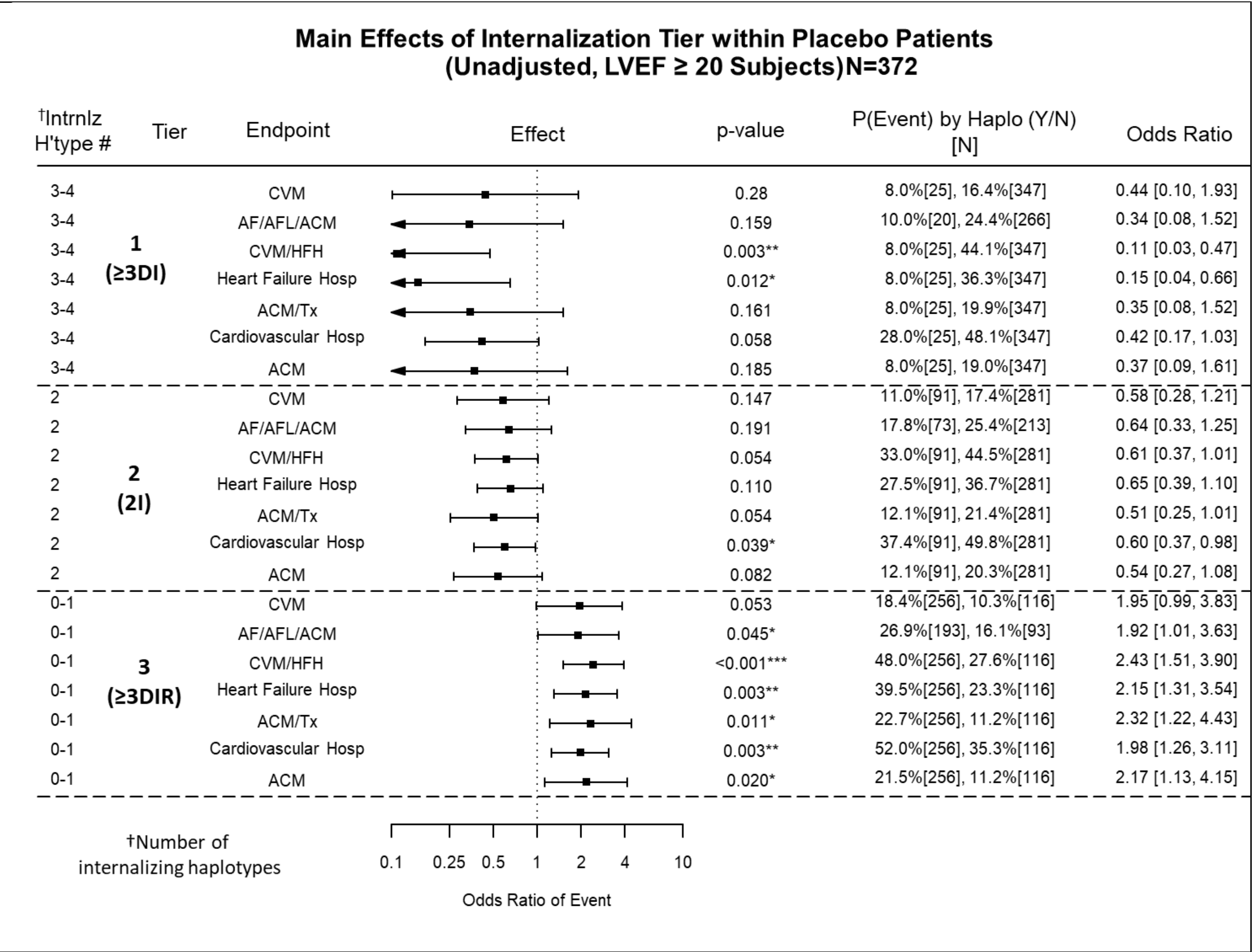


**Figure S1.** LVEF ≥0.20 cohort patients randomized to placebo in the BEST Adrenergic Receptor Polymorphism substudy, forest plot of *ADRB1* + *ADRB2* combination diplotypes arranged by the number of internalizing haplotypes (Column 1), displaying Odds Ratios (ORs, Columns 4 and 7) between event rates (Column 6) in the Column 1 diplotypes vs. all other combination diplotypes. The *ADRB1*, *ADRB2* diplotype combinations are displayed by "≥3 Double Internalizing" combinations in **Tier 1** (**≥3DI**, 1 or 2 copies of *ADRB1* Gly49Arg389 AND *ADRB2* Arg16Gln27, for a total of 3-4 copies of an internalizing haplotype); "Incompletely Internalizing" in **Tier 2** (**2I**), 2 total copies of internalizing haplotypes, either 1 copy of *ADRB1* AND *ADRB2*, or 2 copies on one of them); and "≥3 Double Internalization-resistant" haplotype combinations in **Tier 3** (**≥3IR,** 0-1 total copies of *ADRB1* Gly49Arg389 AND *ADRB2* Arg16Gln27, 3-4 copies of internalization-resistant haplotypes). The counterpart, comparison haplotypes used to generate ORs follow OR logic, i.e. if either the *ADRB1* or the *ADRB2* haplotype doesn't match the cognate Column 1 haplotype then the pair becomes a member of the counterpart group. Column 6 lists the event rate and [N] in the paired haplotypes in column 1, followed by the event rate and [N] in the counterpart, comparator groups. CVM=Cardiovascular Mortality; AF/AFL/ACM=Atrial fibrillation, Atrial Flutter or All-Cause Mortality; CVM/HFH=CVM or Heart Failure hospitalization; HFH=Heart Failure Hospitalization; ACM/Tx=ACM or Cardiac Transplantation; CVH=Cardiovascular Hospitalization; ACM=All-Cause Mortality. For the AF/AFL/ACM endpoint, only non-AF patients at the time of randomization are analyzed, resulting in a smaller N than for the 6 HF endpoints. For all endpoints only first events are considered. See **Figure 1** for OR calculation formula.

#### Main Effects of Internalization Tier within Bucindolol Patients (Unadjusted, LVEF ≥ 20 Subjects)N=351

| †Intrnlz H'type # | Tier | Endpoint | p-value | P(Event) by Haplo (Y/N) [N] | Odds Ratio |
|---|---|---|---|---|---|
| 3-4 | 1 (≥3DI) | CVM | 0.88 | 8.0%[25], 8.9%[326] | 0.89 [0.20, 3.97] |
| 3-4 | | AF/AFL/ACM | 0.35 | 25.0%[20], 16.7%[252] | 1.67 [0.57, 4.83] |
| 3-4 | | CVM/HFH | 0.85 | 28.0%[25], 29.8%[326] | 0.92 [0.37, 2.27] |
| 3-4 | | Heart Failure Hosp | 0.90 | 24.0%[25], 25.2%[326] | 0.94 [0.36, 2.43] |
| 3-4 | | ACM/Tx | 0.62 | 16.0%[25], 12.6%[326] | 1.32 [0.43, 4.05] |
| 3-4 | | Cardiovascular Hosp | 0.59 | 40.0%[25], 34.7%[326] | 1.26 [0.55, 2.89] |
| 3-4 | | ACM | 0.55 | 16.0%[25], 12.0%[326] | 1.40 [0.46, 4.30] |
| 2 | 2 (2I) | CVM | 0.96 | 9.0%[78], 8.8%[273] | 1.02 [0.42, 2.47] |
| 2 | | AF/AFL/ACM | 0.51 | 14.5%[62], 18.1%[210] | 0.77 [0.35, 1.69] |
| 2 | | CVM/HFH | 0.60 | 32.1%[78], 28.9%[273] | 1.16 [0.67, 1.99] |
| 2 | | Heart Failure Hosp | 0.47 | 28.2%[78], 24.2%[273] | 1.23 [0.70, 2.17] |
| 2 | | ACM/Tx | 0.70 | 11.5%[78], 13.2%[273] | 0.86 [0.39, 1.87] |
| 2 | | Cardiovascular Hosp | 0.93 | 34.6%[78], 35.2%[273] | 0.98 [0.58, 1.66] |
| 2 | | ACM | 0.83 | 11.5%[78], 12.5%[273] | 0.92 [0.42, 2.00] |
| 0-1 | 3 (≥3DIR) | CVM | 0.97 | 8.9%[248], 8.7%[103] | 1.02 [0.45, 2.29] |
| 0-1 | | AF/AFL/ACM | 0.95 | 17.4%[190], 17.1%[82] | 1.02 [0.51, 2.03] |
| 0-1 | | CVM/HFH | 0.70 | 29.0%[248], 31.1%[103] | 0.91 [0.55, 1.50] |
| 0-1 | | Heart Failure Hosp | 0.56 | 24.2%[248], 27.2%[103] | 0.85 [0.51, 1.44] |
| 0-1 | | ACM/Tx | 0.94 | 12.9%[248], 12.6%[103] | 1.03 [0.51, 2.04] |
| 0-1 | | Cardiovascular Hosp | 0.82 | 34.7%[248], 35.9%[103] | 0.95 [0.59, 1.53] |
| 0-1 | | ACM | 0.89 | 12.1%[248], 12.6%[103] | 0.95 [0.48, 1.91] |

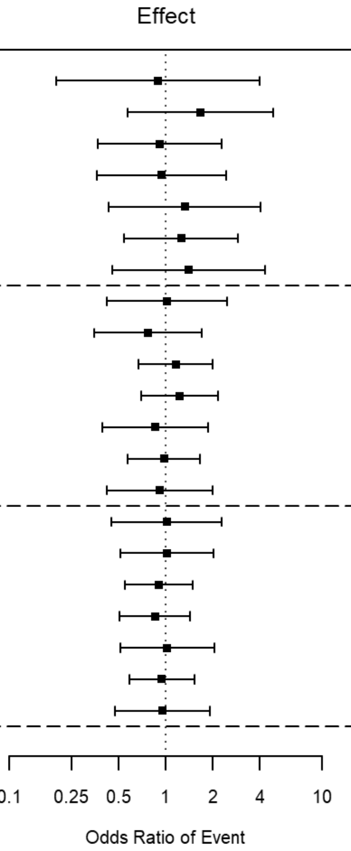


†Number of internalizing haplotypes

**Figure S2.** BEST substudy Patients with LVEFs ≥0.20 randomized to bucindolol in the BEST Adrenergic Receptor Polymorphisms substudy. The forest plot is constructed as for **Figure S1**.

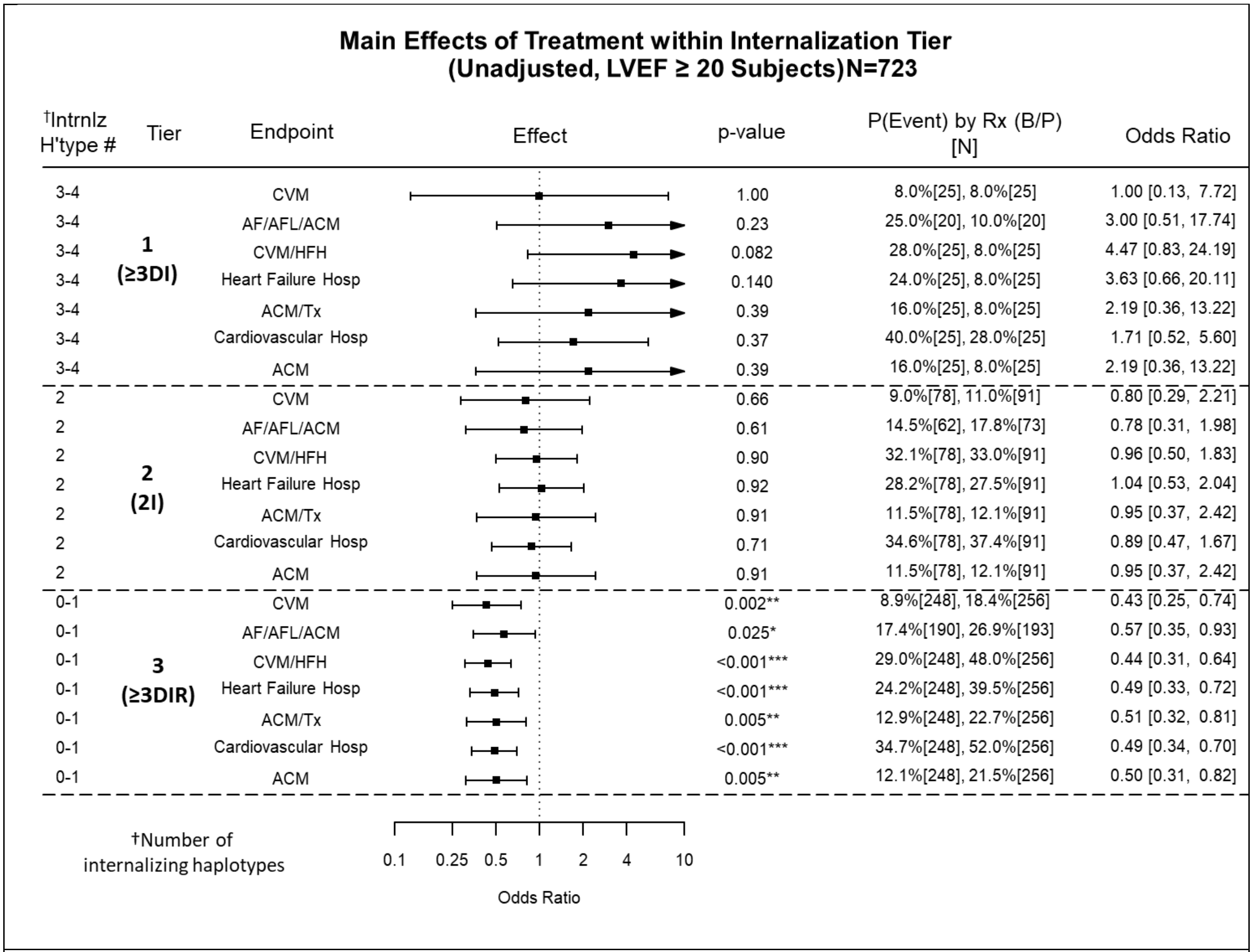


**Figure S3.** BEST substudy patients with LVEFs ≥0.20, forest plot of bucindolol vs. placebo treatment effect Odds Ratios (ORs) by haplotype, set-up otherwise as in Figure 3.

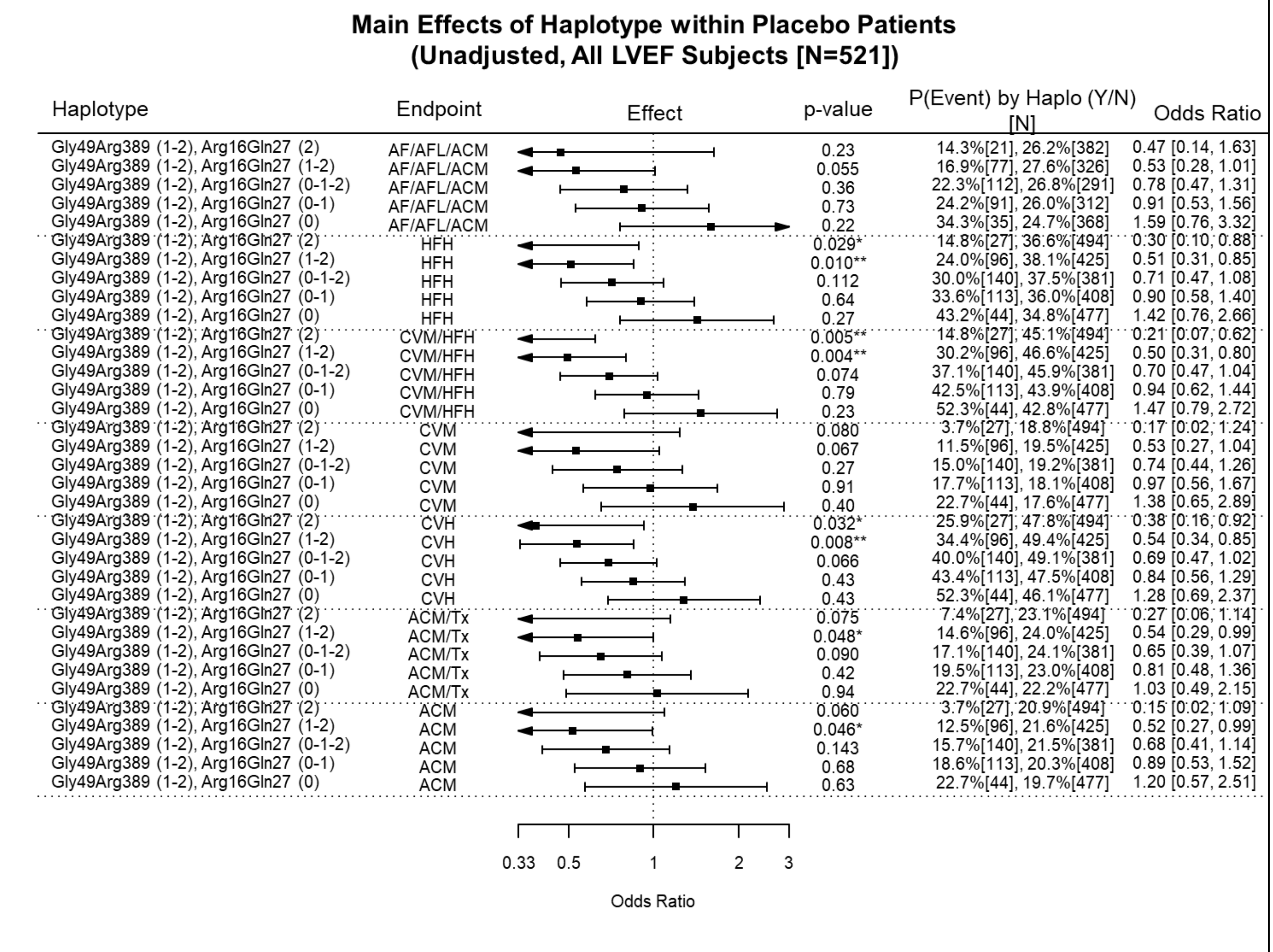


**Figure S4.** Odds Ratios in placebo treated subjects from the BEST Adrenergic Receptor Polymorphisms substudy by designated *ADRB1* and *ADRB2* haplotype combinations, with *ADRB1* Gly49Arg389 held constant and decreasing copies of Arg16Gln27 vs. all other Arg16Gln27 combinations, for 7 clinical endpoints (All LVEF cohort, unadjusted analysis, all races).

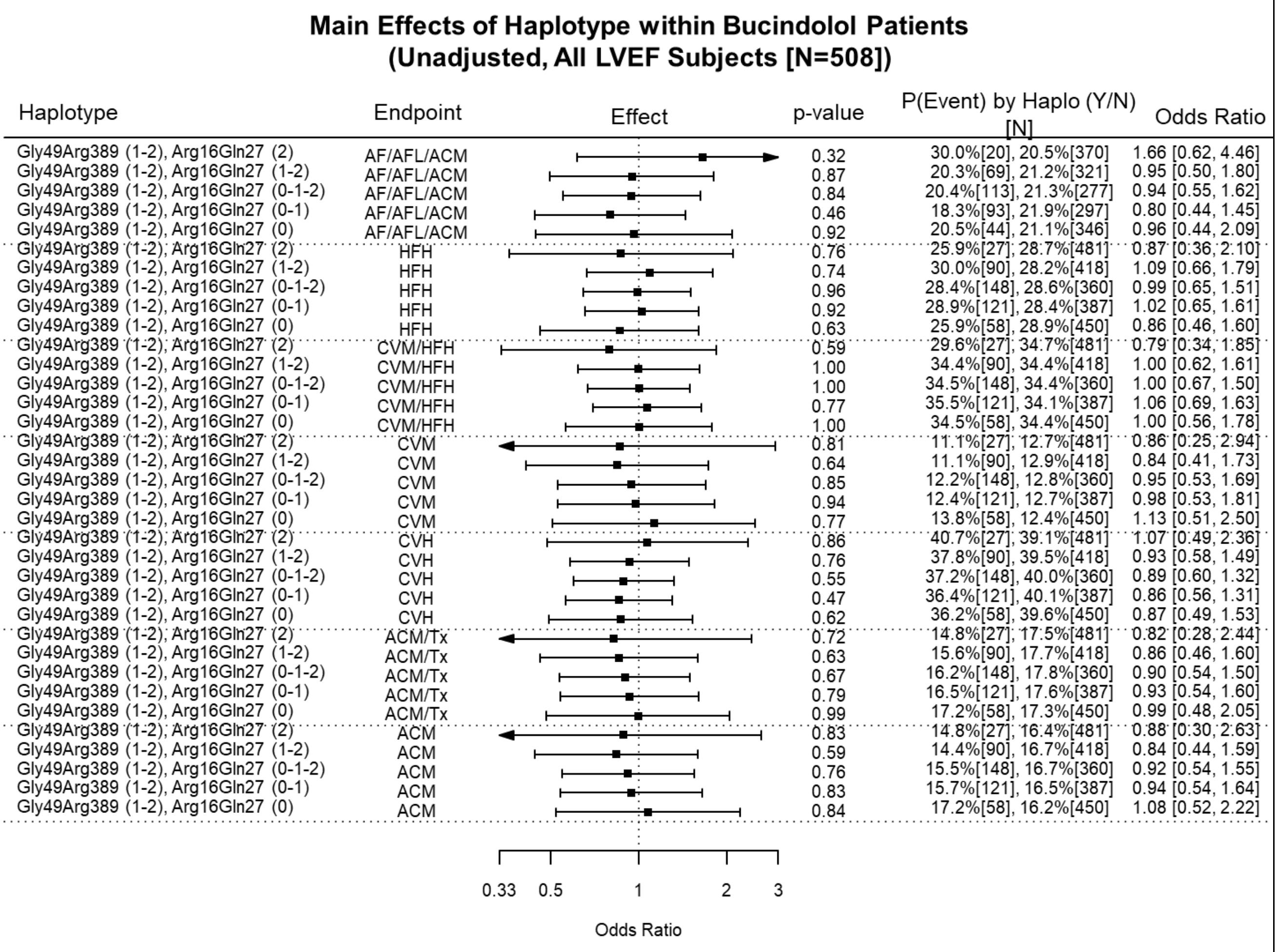


**Figure S5.** Odds Ratios in bucindolol treated subjects from the BEST Adrenergic Receptor Polymorphism substudy by designated *ADRB1* and *ADRB2* haplotype combinations with *ADRB1* Gly49Arg389 held constant and decreasing copies of Arg16Gln27 vs. all other Arg16Gln27 combinations, for 7 clinical endpoints (All LVEF cohort, unadjusted analysis, all races).

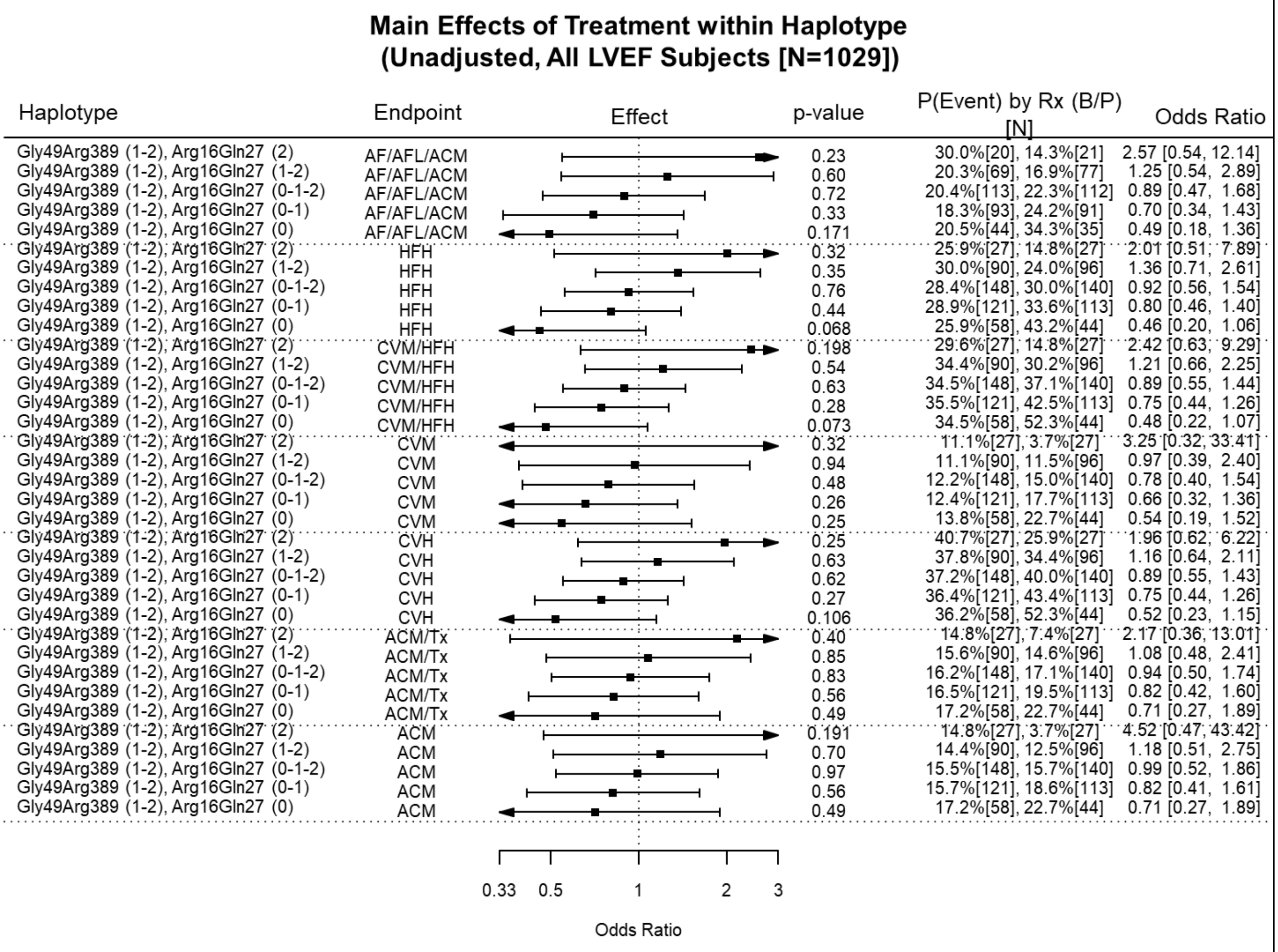


**Figure S6.** Odds Ratios for bucindolol vs. placebo treatment effects from the BEST Adrenergic Receptor Polymorphisms substudy by designated *ADRB1* and *ADRB2* haplotype combinations with *ADRB1* Gly49Arg389 held constant and decreasing copies of Gln27Arg16 vs. all other Gln27Arg16 combinations, for 7 clinical endpoints (All LVEF cohort, unadjusted analysis, all races).

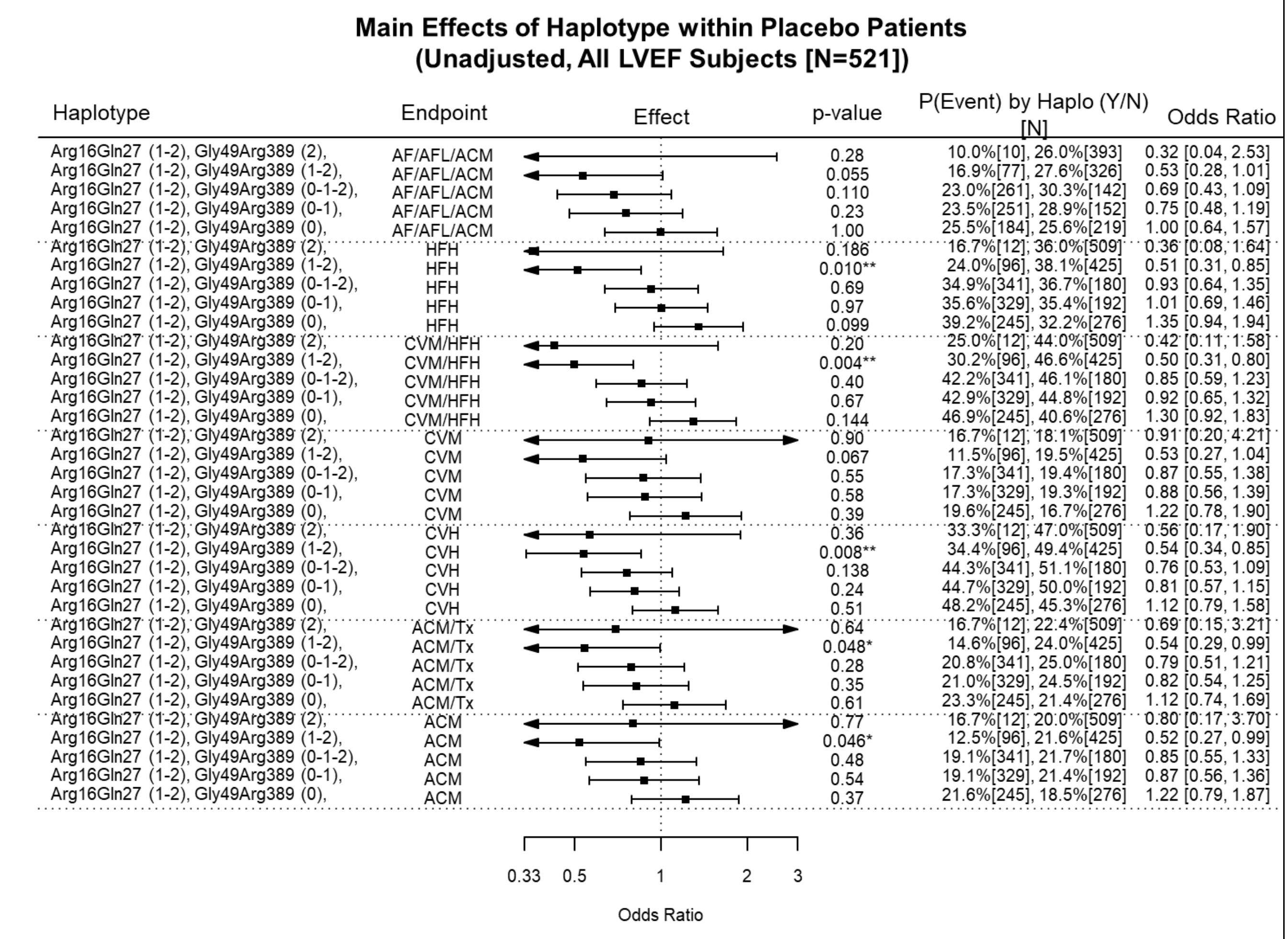


**Figure S7.** Odds Ratios in placebo treated subjects effects from the BEST Adrenergic Receptor Polymorphisms substudy by designated *ADRB1* and *ADRB2* haplotype combinations, with *ADRB2* Arg16Gln27 held constant and decreasing copies of Gly49Arg389 vs. all other Gly49Arg389 combinations, for 7 clinical endpoints (All LVEF cohort, unadjusted analysis, all races).

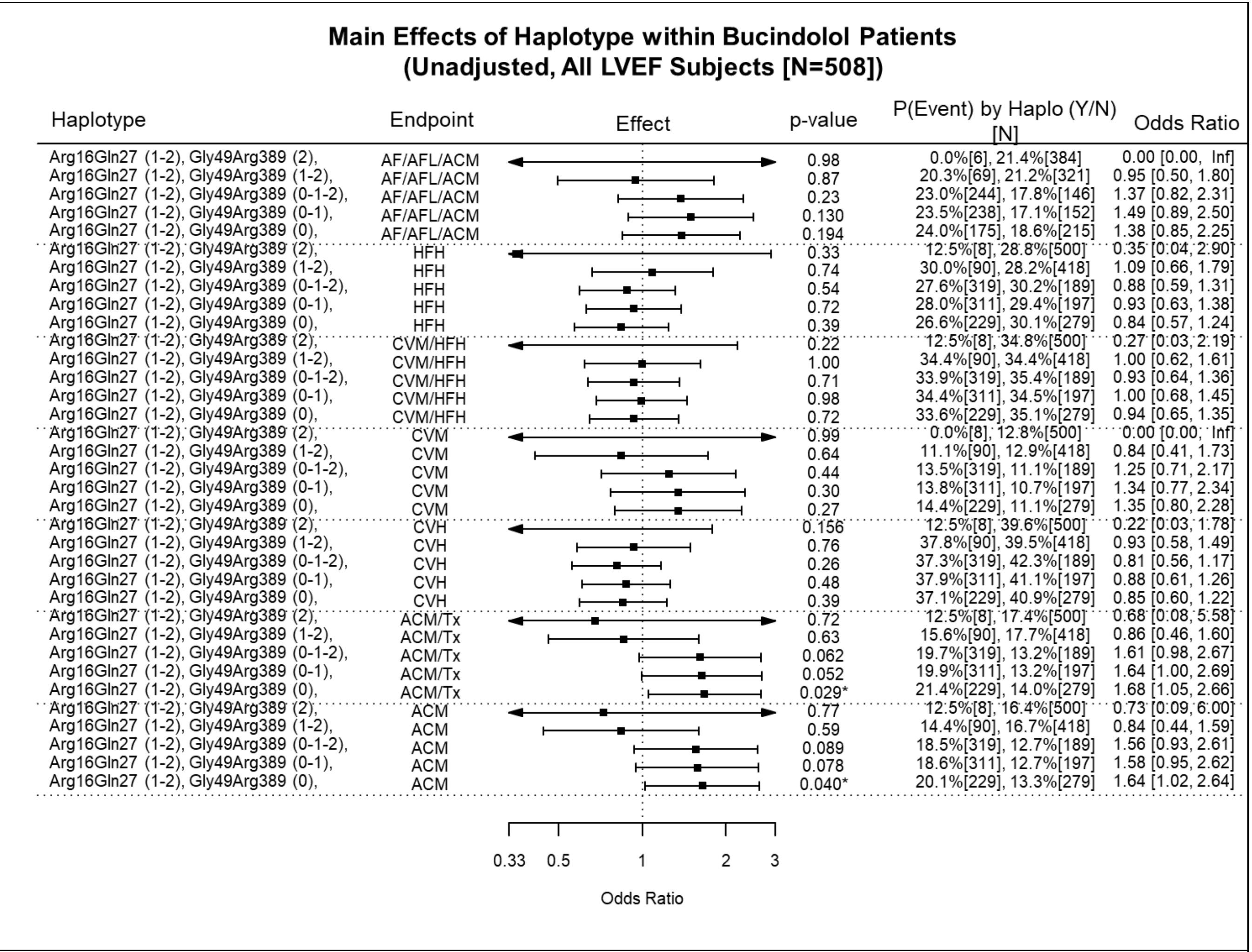


**Figure S8.** Odds Ratios in bucindolol treated subjects effects from the BEST Adrenergic Receptor Polymorphisms substudy by designated *ADRB1* and *ADRB2* haplotype combinations, with *ADRB2* Arg16Gln27 held constant and decreasing copies of Gly49Arg389 vs. all other Gly49Arg389 combinations, for 7 clinical endpoints (All LVEF cohort, unadjusted analysis, all races).

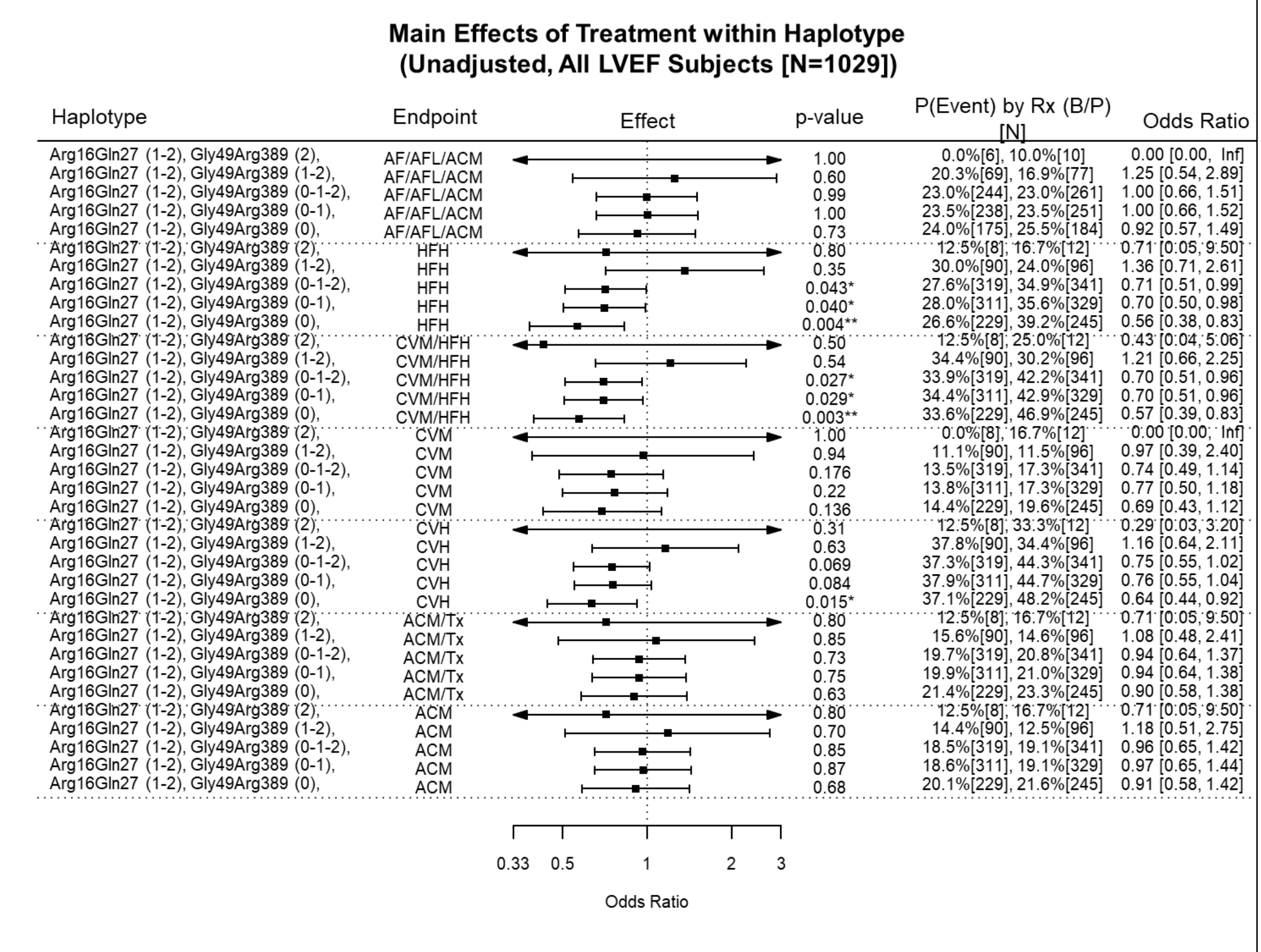


**Figure S9.** Odds Ratios for bucindolol vs. placebo treatment effects from the BEST Adrenergic Receptor Polymorphisms substudy by designated *ADRB1* and *ADRB2* haplotype combinations, with *ADRB2* Arg16Gln27 held constant and decreasing copies of Gly49Arg389 vs. all other Gly49Arg389G combinations, for 7 clinical endpoints (All LVEF cohort, unadjusted analysis, all races).

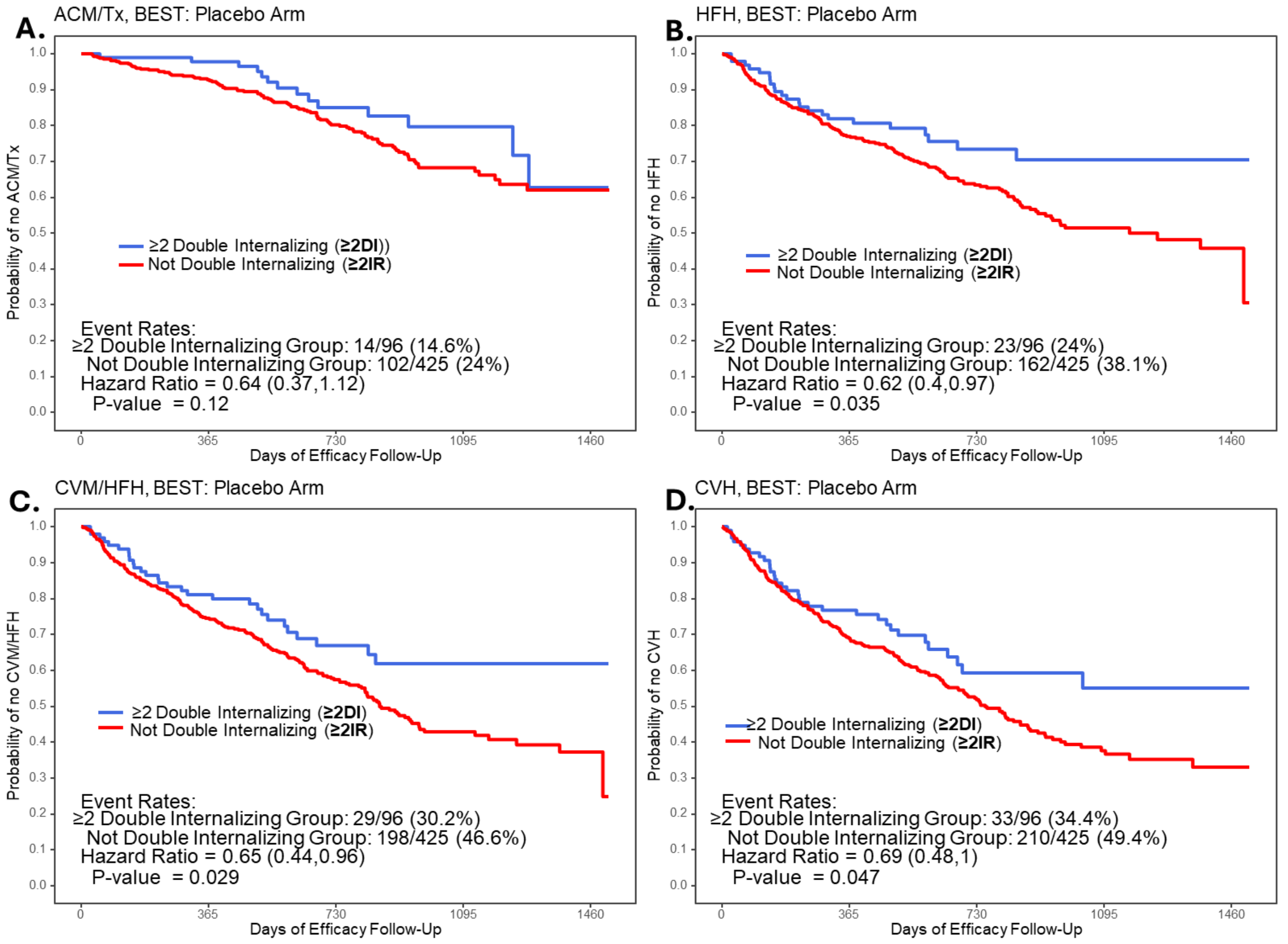


**Fig S10.** Time to first event curves in placebo treated BEST substudy All LVEF cohort patients with ≥1 copy (2-4 total copies) of internalizing haplotypes for both *ADRB1* AND *ADRB2* receptors "≥2 Double Internalizing", ≥2DI), vs. all other β-AR receptor diplotypes ("≥2 Internalization-resistant" (Not double Internalizing), ≥2IR). **A.** All-cause Mortality or Cardiac Transplantation (ACM/Tx); **B.** Heart Failure Hospitalization (HFH); **C.** Cardiovascular Mortality or Heart Failure Hospitalization (CVM/HFH); **D.** Cardiovascular Hospitalization (CVH). Double internalizing receptor haplotypes are associated with improved outcomes in all endpoints.

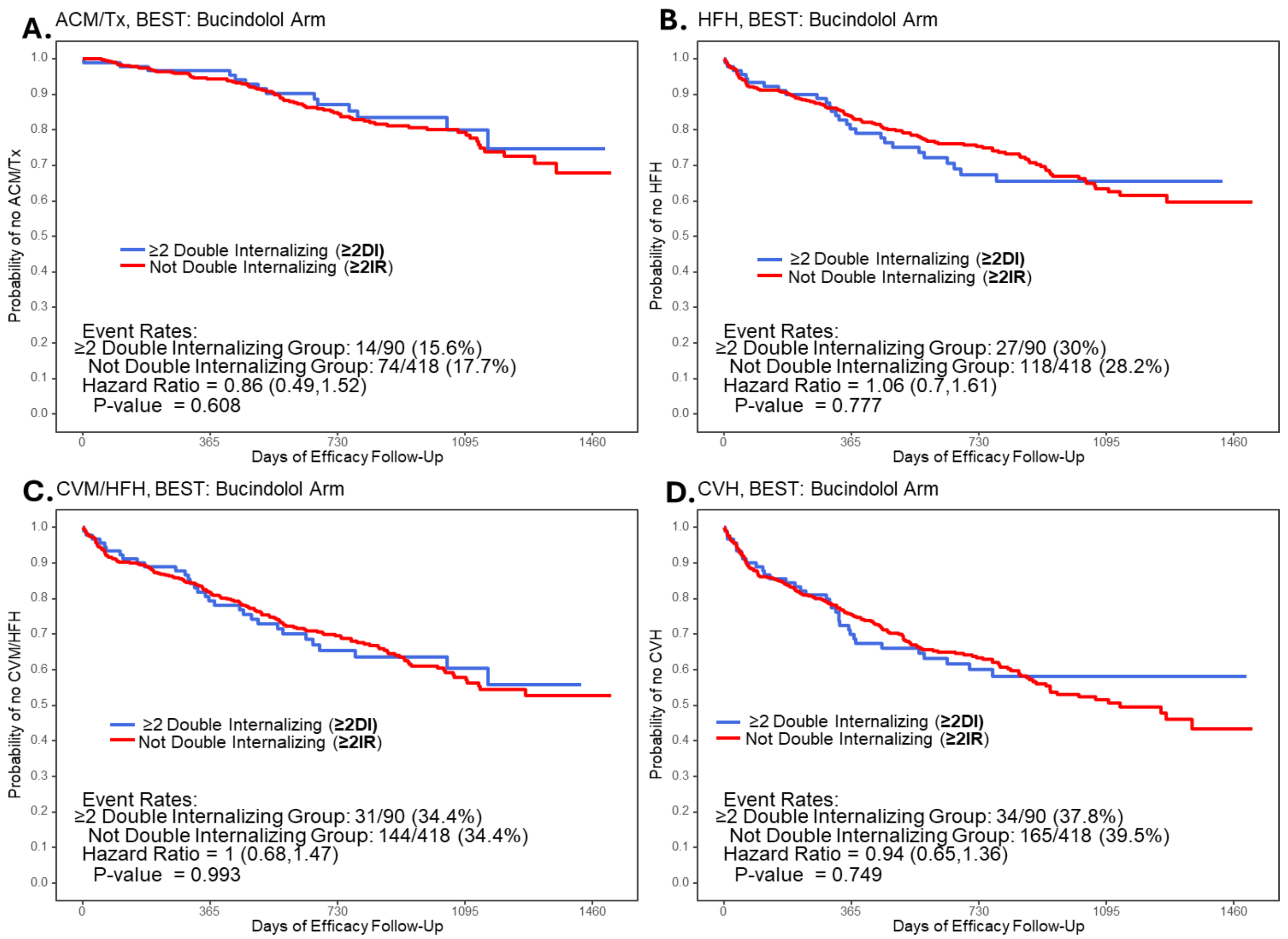


**Fig S11.** Time to first event curves in bucindolol treated BEST substudy All LVEF cohort patients with ≥1 copy (2-4 total copies) of internalizing haplotypes for both *ADRB1* AND *ADRB2* receptors ("≥2 Double Internalizing", ≥2DI), vs. all other β-AR receptor diplotypes ("≥2 Internalization-resistant", ≥2IR).). **A.** All-cause Mortality or Cardiac Transplantation (ACM/Tx); **B.** Heart Failure Hospitalization (HFH); **C.** Cardiovascular Mortality or Heart Failure Hospitalization (CVM/HFH); **D.** Cardiovascular Hospitalization (CVH). Bucindolol is equally effective in both types of receptor internalization, presumably by internalizing relatively internalization-resistant haplotypes.

#### Main Effects of Internalization Tier within Placebo Patients (Unadjusted, All LVEF Subjects; N= 97 Blacks, 424 non-Blacks)

| Internalization Tier | Endpoint | p-value | P(Event) by Haplo (Y/N) [N] | Odds Ratio |
|---|---|---|---|---|
| 3-4 (Black) | ACM | 0.54 | 9.1%[11], 16.3%[86] | 0.51 [0.06, 4.34] |
| 2 (Black) | ACM | 0.99 | 15.4%[26], 15.5%[71] | 0.99 [0.29, 3.44] |
| 0-1 (Black) | ACM | 0.68 | 16.7%[60], 13.5%[37] | 1.28 [0.40, 4.09] |
| 3-4 (Non-Black) | ACM | 0.119 | 8.0%[25], 21.8%[399] | 0.31 [0.07, 1.35] |
| 2 (Non-Black) | ACM | 0.27 | 17.3%[110], 22.3%[314] | 0.73 [0.42, 1.28] |
| 0-1 (Non-Black) | ACM | 0.062 | 23.5%[289], 15.6%[135] | 1.67 [0.97, 2.86] |
| 3-4 (Black) | ACM/Tx | 0.54 | 9.1%[11], 16.3%[86] | 0.51 [0.06, 4.34] |
| 2 (Black) | ACM/Tx | 0.99 | 15.4%[26], 15.5%[71] | 0.99 [0.29, 3.44] |
| 0-1 (Black) | ACM/Tx | 0.68 | 16.7%[60], 13.5%[37] | 1.28 [0.40, 4.09] |
| 3-4 (Non-Black) | ACM/Tx | 0.165 | 12.0%[25], 24.6%[399] | 0.42 [0.12, 1.43] |
| 2 (Non-Black) | ACM/Tx | 0.28 | 20.0%[110], 25.2%[314] | 0.74 [0.44, 1.27] |
| 0-1 (Non-Black) | ACM/Tx | 0.081 | 26.3%[289], 18.5%[135] | 1.57 [0.95, 2.61] |
| 3-4 (Black) | Cardiovascular Hosp | 0.072 | 18.2%[11], 48.8%[86] | 0.23 [0.05, 1.14] |
| 2 (Black) | Cardiovascular Hosp | 0.71 | 42.3%[26], 46.5%[71] | 0.84 [0.34, 2.09] |
| 0-1 (Black) | Cardiovascular Hosp | 0.114 | 51.7%[60], 35.1%[37] | 1.97 [0.85, 4.59] |
| 3-4 (Non-Black) | Cardiovascular Hosp | 0.26 | 36.0%[25], 47.6%[399] | 0.62 [0.27, 1.43] |
| 2 (Non-Black) | Cardiovascular Hosp | 0.033* | 38.2%[110], 50.0%[314] | 0.62 [0.40, 0.96] |
| 0-1 (Non-Black) | Cardiovascular Hosp | 0.010* | 51.2%[289], 37.8%[135] | 1.73 [1.14, 2.62] |
| 3-4 (Black) | CVM | 0.60 | 9.1%[11], 15.1%[86] | 0.56 [0.07, 4.76] |
| 2 (Black) | CVM | 0.62 | 11.5%[26], 15.5%[71] | 0.71 [0.18, 2.78] |
| 0-1 (Black) | CVM | 0.43 | 16.7%[60], 10.8%[37] | 1.65 [0.48, 5.70] |
| 3-4 (Non-Black) | CVM | 0.169 | 8.0%[25], 19.5%[399] | 0.36 [0.08, 1.55] |
| 2 (Non-Black) | CVM | 0.62 | 17.3%[110], 19.4%[314] | 0.87 [0.49, 1.53] |
| 0-1 (Non-Black) | CVM | 0.23 | 20.4%[289], 15.6%[135] | 1.39 [0.81, 2.40] |
| 3-4 (Black) | CVM/HFH | 0.043* | 9.1%[11], 46.5%[86] | 0.12 [0.01, 0.94] |
| 2 (Black) | CVM/HFH | 1.00 | 42.3%[26], 42.3%[71] | 1.00 [0.40, 2.49] |
| 0-1 (Black) | CVM/HFH | 0.126 | 48.3%[60], 32.4%[37] | 1.95 [0.83, 4.58] |
| 3-4 (Non-Black) | CVM/HFH | 0.046* | 24.0%[25], 45.1%[399] | 0.38 [0.15, 0.98] |
| 2 (Non-Black) | CVM/HFH | 0.163 | 38.2%[110], 45.9%[314] | 0.73 [0.47, 1.14] |
| 0-1 (Non-Black) | CVM/HFH | 0.019* | 47.8%[289], 35.6%[135] | 1.66 [1.09, 2.52] |
| 3-4 (Black) | Heart Failure Hosp | 0.072 | 9.1%[11], 40.7%[86] | 0.15 [0.02, 1.19] |
| 2 (Black) | Heart Failure Hosp | 0.87 | 38.5%[26], 36.6%[71] | 1.08 [0.43, 2.73] |
| 0-1 (Black) | Heart Failure Hosp | 0.24 | 41.7%[60], 29.7%[37] | 1.69 [0.71, 4.04] |
| 3-4 (Non-Black) | Heart Failure Hosp | 0.111 | 20.0%[25], 36.1%[399] | 0.44 [0.16, 1.20] |
| 2 (Non-Black) | Heart Failure Hosp | 0.124 | 29.1%[110], 37.3%[314] | 0.69 [0.43, 1.11] |
| 0-1 (Non-Black) | Heart Failure Hosp | 0.023* | 38.8%[289], 27.4%[135] | 1.68 [1.07, 2.62] |
| 3-4 (Black) | AF/AFL/ACM | 0.99 | 0.0%[10], 24.3%[74] | 0.00 [0.00, Inf] |
| 2 (Black) | AF/AFL/ACM | 0.76 | 23.8%[21], 20.6%[63] | 1.20 [0.37, 3.89] |
| 0-1 (Black) | AF/AFL/ACM | 0.37 | 24.5%[53], 16.1%[31] | 1.69 [0.54, 5.30] |
| 3-4 (Non-Black) | AF/AFL/ACM | 0.66 | 22.2%[18], 26.9%[301] | 0.78 [0.25, 2.43] |
| 2 (Non-Black) | AF/AFL/ACM | 0.162 | 20.9%[86], 28.8%[233] | 0.66 [0.36, 1.19] |
| 0-1 (Non-Black) | AF/AFL/ACM | 0.124 | 29.3%[215], 21.2%[104] | 1.54 [0.89, 2.69] |

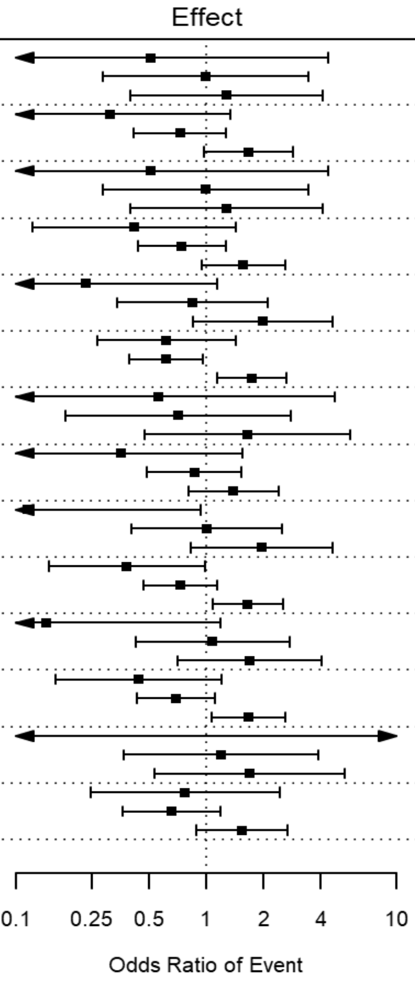


**Figure S12.** Placebo treatment forest plot, Black/non-Black subjects by internalization tier, All LVEF BEST substudy cohort: **Tier 1, ≥3DI**, 3-4 total copies of *ADRB1* Arg or Gly389 AND *ADRB2* Gln27Arg16 (internalizing) haplotypes; **Tier 2**, **2I,** 2 total copies of internalizing haplotypes, either 1 copy of *ADRB1* AND *ADRB2*, or 2 copies on one of them); **Tier 3**, **≥3IR** ,0-1 total copies *ADRB1* OR *ADRB2* internalizing haplotypes.

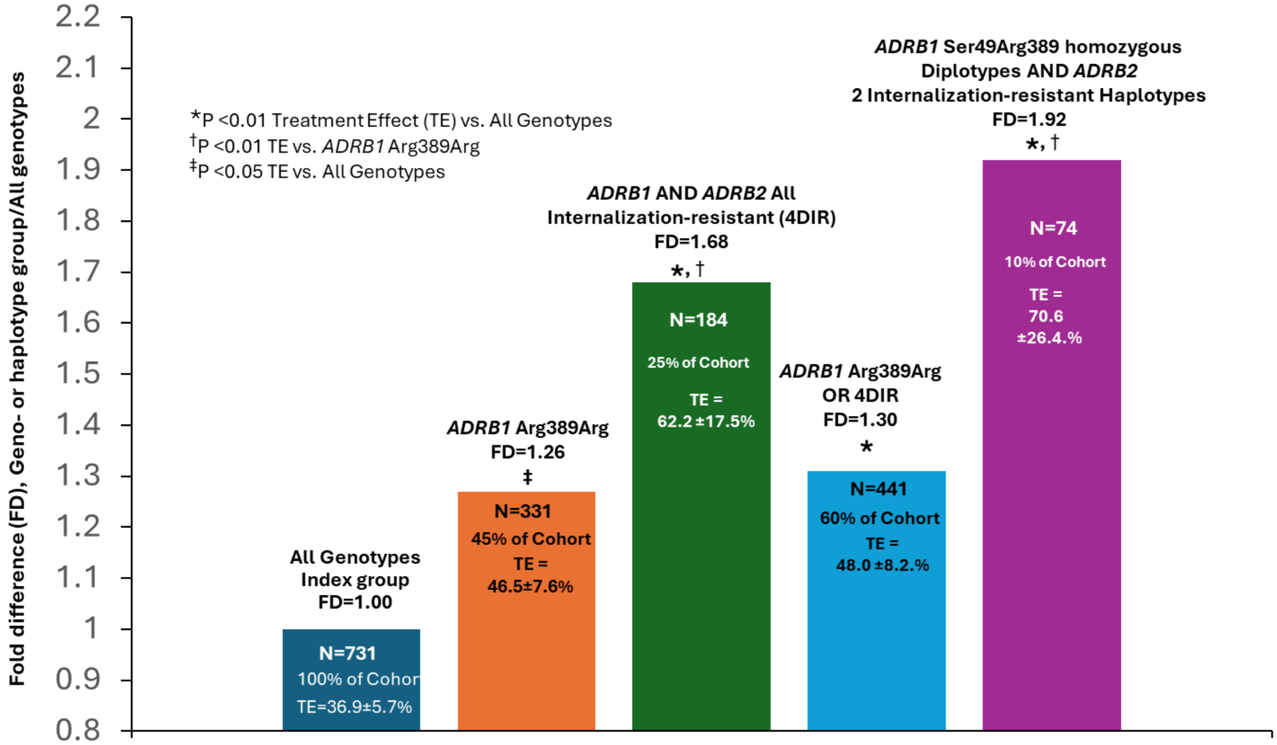


**Figure S13.** BEST Adrenergic Receptor Polymorphisms substudy, LVEF ≥0.20 cohort, TE = Treatment Effect % ((1-hazard ratio) x 100) expressed as fold difference (FD) vs. All Genotypes/haplotypes subjects (Column/bar 1): Column/bar 2, subjects with an *ADRB1* Arg389Arg genotype; Column/bar 3, maximum *ADRB1* AND *ADRB2* internalization-resistant diplotypes (N=4, "4DIR"); Column/bar 4, *ADRB1* Arg389Arg OR *ADRB1* AND *ADRB2* 4DIR Internalization-resistant diplotypes; Column/bar 5, *ADRB1* Arg389Ser49 homozygous diplotype AND 2 *ADRB2* Internalization-resistant diplotypes (4DIR with 2 ADRB1 Ser 49Arg389 haplotypes, and the 2 ADRB2 haplotypes being "not Arg16Gln27")
